# Social Compliance with NPIs, Mobility Patterns, and Reproduction Number: Lessons from COVID-19 in Europe

**DOI:** 10.1101/2025.01.10.25320334

**Authors:** Daniele Baccega, Jose Aguilar, Carlos Baquero, Antonio Fernández Anta, Juan Marcos Ramirez

## Abstract

Non-pharmaceutical interventions (NPIs), including measures such as lockdowns, travel limitations, and social distancing mandates, play a critical role in shaping human mobility, which subsequently influences the spread of infectious diseases. Using COVID-19 as a case study, this research examines the relationship between restrictions, mobility patterns, and the disease’s effective reproduction number (R_t_) across 13 European countries. Employing clustering techniques, we uncover distinct national patterns, highlighting differences in social compliance between Northern and Southern Europe. While restrictions strongly correlate with mobility reductions, the relationship between mobility and R_t_ is more nuanced, driven primarily by the nature of social interactions rather than mere compliance. Additionally, employing XGBoost regression models, we demonstrate that missing mobility data can be accurately inferred from restrictions, and missing infection rates can be predicted from mobility data. These findings provide valuable insights for tailoring public health strategies in future crisis and refining analytical approaches.

## 1 Introduction

Government measures can be categorized into two groups: pharmaceutical and non-pharmaceutical. Pharmaceutical interventions (PIs) include medical treatments aimed at preventing, managing, or curing diseases, such as vaccines, antiviral medications, monoclonal antibodies, immunomodulators, and antibiotics. On the other hand, non-pharmaceutical interventions (NPIs) focus on strategies such as social distancing, quarantine and isolation, screening campaigns, travel restrictions, mask-wearing, hygiene measures, remote work, closure of educational institutions and non-essential businesses, limitations on public gatherings, and contact tracing. These interventions are implemented to prevent or control the outbreak of an emergency, like the spread of a disease in a population. In this context, NPIs help to reduce the transmission through changes in behaviours, environments, and community interactions [36].

One of the most immediate and observable effects of NPIs has been their impact on human mobility. Mobility, which refers to the movement of individuals within and between geographic areas, plays a crucial role in the transmission dynamics of infectious diseases. The relationship between NPIs and mobility is complex and multifaceted. On the one hand, stringent measures such as nationwide lock-downs and curfews have led to dramatic declines in mobility, as evidenced by data from mobility tracking tools and transportation statistics [28, 2]. On the other hand, the relaxation of these measures has often been followed by a resurgence in mobility, sometimes correlating with subsequent waves of infections. Moreover, variations in compliance and enforcement, socio-economic factors, and public trust in government policies have influenced the effectiveness of NPIs and the extent of mobility reductions [1, 17].

The COVID-19 pandemic—caused by SARS-CoV-2—, which began in late 2019 and ended in May 2023, has led to widespread illness, economic disruption, and millions of deaths globally, demonstrating the far-reaching impact of emerging infectious diseases in a highly interconnected world [18]. Initially identified in Wuhan, China, the virus quickly spread worldwide, leading to an unprecedented global health crisis. In response to the rapid transmission and severe consequences of COVID-19, governments and public health authorities implemented a wide range of measures to mitigate the transmission of the virus and manage healthcare resources [37]. As of end-2024, the pandemic has resulted in over 800 million confirmed cases and more than 7 million deaths worldwide—data from the World Health Organization (WHO) [49].

During the COVID-19 pandemic, changes in mobility patterns have been both a direct consequence of policy measures and a reflection of individual behavioral responses to the perceived risk of infection. The reduction in mobility, especially through lock-downs and travel bans, aimed to decrease the opportunities for person-to-person transmission, thereby slowing the spread of the virus and reducing the burden on healthcare systems [4, 42, 9]. The implementation of NPIs has varied significantly across different regions and periods, reflecting diverse epidemiological, political, and socio-economical contexts [36].

The differences among countries in their approach to NPIs and their impact on mobility are striking. For instance, countries with robust healthcare systems and high levels of public trust in government, such as Germany, saw more consistent compliance with mobility restrictions, resulting in more effective control of the effective reproduction number R_t_—the average number of secondary infections generated by an infected individual in a population at a given time. In contrast, countries with fragmented healthcare systems and lower public trust, such as the USA and Brazil, experienced greater challenges in enforcing NPIs and saw less dramatic declines in mobility [8, 15]. Social compliance, shaped by trust in institutions and adherence to societal norms, rules, and regulations, played a crucial role in the effectiveness of non-pharmaceutical interventions [47, 34]. In countries where individuals more readily adhered to collective public health goals, NPIs proved to be more effective in reducing disease spread. Cultural factors, such as societal norms, values, and attitudes toward government authority, further influenced these outcomes. For example, in East Asian countries, where there is a strong tradition of community responsibility and compliance with public health measures, NPIs were more effective in reducing mobility compared to Western nations, where individual freedoms are more heavily emphasized [31]. Additionally, social interactions, representing the dynamic exchanges between individuals or groups that collectively shape relationships, cultural norms, and societal structures, contributed significantly to the effectiveness of these measures—where social networks facilitated compliance or, conversely, hindered adherence in regions with less social cohesion. Economic disparities also played a role, as lower-income countries and communities often faced greater difficulties in adhering to lock-down measures due to the necessity of continuing economic activities. These variations underscore the importance of context-specific strategies that consider both social compliance and the socio-economic landscape in managing mobility and R_t_ during a pandemic [33].

Thus, mobility data plays a critical role in understanding the dynamics of infectious disease transmission. Still, it often contains significant gaps due to limited geographic coverage, inconsistent data collection practices, and varying levels of technology adoption. Similarly, R_t_ data can be impacted by irregular reporting, testing backlogs, and inconsistent public health monitoring across different regions. These data limitations hinder the evaluation of policy impacts and the development of reliable predictive models. To overcome these challenges, it is essential to employ advanced methods that address these gaps, allowing for more accurate insights into how NPIs influence mobility patterns and, ultimately, the spread of infectious diseases. This leads to better-informed policy decisions and more effective public health interventions.

### 1.1 Contribution

This study investigates the relationship between government-imposed restrictions, mobility patterns, and the effective reproduction number R_t_ of COVID-19. We analyze mobility and restrictions data from major platforms to understand how movement trends respond to policy changes, providing insights into disease transmission dynamics across diverse countries and demographics. Our focus is on behavioral patterns and intervention responses. Specifically, we analyze data from several European countries, including Finland, Norway, Sweden, Belgium, Ireland, the United Kingdom, France, Germany, Austria, Italy, Greece, Spain, and Portugal. Our main contributions are as follows:

1. We identify a strong correlation between the considered restrictions and mobility variables, contrasted by a weaker correlation between mobility and infection rates.
2. We identify cultural patterns by analyzing how mobility and the R_t_ are influenced by both official restrictions and self-imposed limitations (in the absence of formal measures), and variations in social compliance and interaction across different countries.
3. Using K-means clustering [27], we examine the distinctions between Northern and Southern European countries in terms of their social compliance with restrictions, while also highlighting a lesser degree of distinction in social interactions. Additionally, in the ‘Supplementary Material’, we apply another clustering technique, CONNECTOR [35], for the same analysis.
4. We apply XGBoost regression models [16] to:

- Estimate missing mobility data using restriction indicators.
- Infer missing R_t_ values from mobility patterns.

Our approach consists of the following steps: we start pre-processing data to obtain mobility variables, restriction indices, and the R_t_. We then calculate three ratios from the average mobility, the stringency index, and the R_t_ to identify cultural patterns. Using clustering methods, we analyze social compliance with restrictions across European nations, noting differences between Northern and Southern countries. Finally, XGBoost models estimate missing mobility data from restrictions and inferred R_t_ values from mobility patterns.

The article is organized as follows. It begins with a review of related research on the impact of NPIs on mobility and COVID-19 spread (Section 2). It then details the data sources and pre-processing steps, including how data was prepared for analysis, and provide preliminary analyses that highlight a strong correlation among the considered variables (Section 3). The methods section describes the analytical techniques used, particularly, clustering and regression models (Section 4). The results section presents findings on mobility patterns, the clustering of countries, and the performance of regression models (Section 5). In the discussion, we interpret the findings in terms of policy implications and behavioral trends (Section 6). The article concludes with a summary of the key results and their contributions to the field (Section 7).

## 2 State of the art

The effect of government-imposed restrictions or NPIs on people’s mobility and the consequent repercussions on the spread of the disease has been explored in several studies [11, 50].

Snoeijer et al. [42] investigate the relative impact of NPIs on mobility changes across multiple countries using Apple and Google mobility data. They found that lock-downs, states of emergency, business and public service closures, and school closures had the largest effect on reducing mobility. Chi-square and cluster analyses revealed that NPIs like school closures and business shutdowns were highly correlated in timing and implementation, suggesting that the observed effects of these interventions may be amplified by their simultaneous enforcement.

Askitas et al. [4] estimate the average dynamic impact of various interventions on COVID-19 incidence and individuals’ movement patterns by developing a statistical model that accounts for the simultaneous implementation of multiple interventions using daily data from 175 countries. Their findings indicate that restrictions on private gatherings and the closure of schools and workplaces significantly reduced COVID-19 infections. In contrast, restrictions on internal movement and public transport had no effects, while international travel restrictions yielded only short-lived effects.

Banholzer et al. [9] evaluate the effectiveness of seven NPIs aimed at controlling the spread of SARS-CoV-2, including school closures and bans on gatherings. Using a semi-mechanistic Bayesian hierarchical model and data from the first wave of COVID-19 across 20 countries, the findings reveal that bans on large gatherings were the most effective in reducing new infections, followed by venue and school closures. In contrast, stay-at-home and work-from-home orders were the least effective.

Cartenì et al. [14] quantify the impact of mobility habits on the spread of COVID-19 in Italy using a multiple linear regression model. The results showed that mobility patterns significantly contributed to explaining the number of infections, alongside other factors such as daily testing rates and environmental variables like air pollution and temperature. These findings highlight the role of mobility, in combination with other key variables, in influencing the transmission of the virus.

Shao et al. [41] examine the relationship between ambient temperature and COVID-19 transmission by examining human mobility data from 47 countries. The findings show that higher temperatures are negatively correlated with transmission rates but positively correlated with human mobility, which increases transmission. This suppression effect suggests that, while warmer weather may reduce transmission rates, it can also lead to increased outdoor activities, potentially worsening COVID-19 spread. Effective control measures are crucial, particularly as temperatures rise, to address the ongoing transmission despite warmer weather in some regions.

Bryant et al. [12] introduce a Bayesian model that estimates daily COVID-19 deaths based on mobility patterns in response to NPIs across Europe. Using Google mobility data from five categories, the model finds strong correlations between mobility shifts and death rates, with reductions in grocery and pharmacy mobility significantly lowering the basic reproduction number R_0_—the average number of secondary infections caused by a single infected individual in a fully susceptible population.

Ilin et al. [29] use simple statistical models—using the same data as Bryant et al. [12] and Facebook data—to show that NPIs significantly reduce mobility, which can effectively predict infections based on data from countries like China, France, Italy, South Korea, and the United States.

Garcìa-Cremades et al. [22] explore the wide-ranging impacts of the COVID-19 pandemic, highlighting that, until herd immunity is achieved, measures like quarantines and social distancing are necessary to reduce mobility, despite their economic repercussions. The study evaluates various prediction models to create a decision support system for policymakers. It proposes consensus strategies to enhance model accuracy and introduces a multivariate model incorporating Google mobility data for better forecasting of 14-day case incidence (CI).

Guan et al. [23] analyze anonymized health and mobility data from Israel to develop predictive models for daily new COVID-19 cases and test positivity rates. Models that incorporate mobility data significantly outperformed those that do not, reducing RMSE by 17.3% for new cases and 10.2% for positivity rates. The findings indicate high accuracy in predicting outbreak severity, offering a valuable tool for policymakers to implement targeted interventions in the post-vaccination era.

Barros et al. [10] investigate the effectiveness of NPIs on COVID-19 transmission by analyzing their impact on the effective reproduction number R_t_ and human mobility, used as a measure of policy adherence. Using a causal inference approach, the study examines five NPIs—confinement, school closures, mask mandates, events, and work restrictions—across 113 countries up to June 2020. The findings reveal that all NPIs, except mask-wearing, significantly influenced mobility trends, with school and events having the greatest effect on social distancing. Moreover, school closures, mask mandates, and work-from-home policies resulted in a sustained reduction in Rt.

Badr et al. [7] examine the impact of social distancing on COVID-19 transmission across the 25 most affected U.S. counties from January to April 2020. Using anonymized cell phone mobility data, the researchers found a strong correlation between reduced mobility and slower COVID-19 case growth rates, with Pearson correlation coefficients above 0.7 in 20 counties. Mobility dropped by 35–63%, with the effect on transmission typically observed 9–12 days later, consistent with the virus’s incubation period. The study also found that behavioral changes in mobility began before official stay-at-home orders were issued, indicating a proactive public response.

Tokey [46] analyzes the relationship between human mobility and COVID-19 transmission across U.S. counties from March to August 2020. It explores spatiotemporal mobility patterns and their correlation with infection rates, controlling for socio-demographic factors and policy changes. The findings indicate that mobility decreased until the first COVID-19 wave peaked, then began to rise. Spatial models reveal a negative correlation between infection rates and mobility indicators, such as miles traveled and out-of-county trips. Higher infection areas had more online workers, fewer older and educated individuals, and higher poverty rates.

Adeniyi et al. [1] analyze a COVID-19 compartmental model that incorporates public compliance with health rules and sanitation, aiming to determine the effective reproduction number R_t_ and system stability. It shows that the system is stable under certain conditions and that combining adherence to rules and sanitation is the most cost-effective strategy for controlling the virus.

Chung et al. [17] investigate how trust in social NPIs influences travel intentions during a pandemic using data from South Korea, one of the few countries that did not impose mobility restrictions after the COVID-19 outbreak. The findings indicate that trust in social NPIs mediates the relationship between efforts to cope with travel restrictions and the intention to travel during the pandemic.

Painter et al. [34] find that residents in U.S. Republican counties are less likely to fully comply with stay-at-home orders compared to those in Democratic counties—using geolocation and debit card transaction data. On the other hand, Democratic counties are more likely to shift to remote spending after such orders are implemented. While factors like COVID-19 risk and geography are considered, political beliefs appear to be a key factor influencing compliance with government mandates, with political alignment potentially explaining these partisan differences.

Van Rooij et al. [47] examine factors influencing Americans’ compliance with stay-at-home measures using a survey of 570 participants from 35 states. It finds that fear of authorities reduces compliance, while personal ability, self-control, and intrinsic motivations like moral support and social norms promote compliance. The findings highlight key factors for improving compliance to mitigate the virus.

In general, these works highlight the crucial role of NPIs and mobility patterns in controlling the spread of COVID-19 infections. Key interventions like school closures, lock-downs, and restrictions on gatherings were effective in reducing mobility and, consequently, transmission rates. However, the effectiveness of specific measures varied, with some studies finding that restrictions on internal movement or public transport had limited or short-lived effects. The correlation between reduced mobility and lower infection rates was a recurring theme, underscoring mobility as a key factor in the pandemic’s dynamics. Some studies also noted that mobility began declining before formal interventions, indicating proactive public behavior. Environmental factors, such as temperature, interacted with mobility and transmission patterns, with higher temperatures potentially increasing mobility, which could counteract reductions in transmission due to weather. Moreover, some studies highlight that compliance with COVID-19 measures is influenced by various factors. Trust in social measures and political beliefs can significantly impact adherence to restrictions, while personal ability, self-control, and intrinsic motivations like moral support play crucial roles. Combining rule adherence with sanitation is found to be particularly effective in controlling the virus.

While numerous studies have examined the impact of NPIs on mobility and the spread of COVID-19, most focus on specific regions, interventions, or mobility data sources, independently. There is a notable lack of comprehensive analyses exploring how mobility patterns across different countries respond to government-imposed and self-imposed restrictions, and how these patterns influence the effective reproduction number R_t_ over time, shaped by cultural patterns, variations in social compliance and interpersonal interactions. Moreover, no study has estimated missing mobility data from restrictions and missing R_t_ data from mobility.

## 3 Materials

This section details the data employed in the study (Section 3.1), providing a comprehensive account of the resources used to achieve the research objectives, including the pre-processing (Section 3.2) and some preliminary analysis (Section 3.3).

### 3.1 Data

We utilized several types of COVID-19 data, including official surveillance data from the COVID-19 Data Hub [25, 24], estimates of active cases computed from the UMD Global CTIS’s survey [5, 38, 3, 39], mobility data from UMD Global CTIS’s survey [5, 21, 30], information on restrictions from the Oxford Covid-19 Government Response Tracker (OxCGRT) [26], and Google mobility data [32]. All datasets used in this work are open-source and publicly available. Data from UMD Global CTIS’s survey are available in an aggregated form—see *Availability of materials and data* section for more details.

The COVID-19 Data Hub is the primary source for surveillance data [25, 24]. Specifically, for each country, we used the cumulative number of fatalities to calculate daily new fatalities and the total population. While the Data Hub offers a comprehensive range of information, including vaccinations, testing numbers, hospitalizations, and intensive care unit occupancy, these additional data are not relevant to our current study. Official COVID-19 mortality data should be generally more reliable than case data due to the consistent and rigorous reporting standards applied to fatalities, unlike the variability and potential under-reporting often seen in case counts—also due to asymptomatic infections. For these reasons, we decided to use estimates of active cases from prior studies [5, 38, 3, 39], which were based on direct symptom responses indicative of COVID-like illness (CLI) proposed in UMD Global CTIS—i.e., fever along with cough, shortness of breath, or difficulty breathing—, along with the cumulative number of fatalities, rather than relying on surveillance data for officially reported cases.

Since the spring of 2020, Facebook has been collaborating in the context of their program Data for Good [20] with the University of Maryland promoting surveys created by this institution [5, 21, 30]. The University of Maryland Global COVID-19 Trends and Impact Survey, in partnership with Facebook (UMD Global CTIS) [21], has been recording daily responses from invited Facebook users on topics related to the COVID-19 pandemic, including RT-PCR results, symptoms, and vaccinations, from over 114 countries. We have obtained access to the responses through an agreement with the University of Maryland (UMD) and Facebook—see Section *Ethical Declaration*. The survey questions are available on the UMD Global CTIS website [45]. In particular, from this platform, we used aggregated mobility information considering the following questions—the labels correspond to those used in the survey:

- In the last 24 hours, have you done any of the following?

**–** *C0 1* : Gone to work outside the place where you are currently staying.
**–** *C0 2* : Gone to a market, grocery store, or pharmacy.
**–** *C0 3* : Gone to a restaurant, cafe, or shopping center.
**–** *C0 4* : Spent time with someone who isn’t currently staying with you.
**–** *C0 5* : Attended a public event with more than 10 people.
**–** *C0 6* : Used public transit.
- *C5* : In the past 7 days, how often did you wear a mask when in public?

Starting from May 20^th^, 2021, all these questions—except *C5* —were modified, leading to a significant change in their meaning and values. For example, question *C0 1* changed from *In the last 24 hours, have you done any of the following? Gone to work outside the place where you are currently staying* to *In the past 24 hours, have you done any of the following? Gone to work or school **indoors**, outside the place where you are currently staying*. The change in values is likely attributable to the use of the word ***indoors*** in the questions. For this reason, we focused the attention on data from April 29^th^, 2020 to May, 19^th^, 2021. We will refer to this data as Facebook data.

To combat the COVID-19 pandemic, countries worldwide have implemented stringent policies (NPIs), such as stay-at-home lock-downs, school and workplace closures, event cancellations, and public transport restrictions, to slow virus spread by enforcing physical distancing. The Oxford Covid-19 Government Response Tracker (OxCGRT) [26] provides information on which pandemic response measures were enacted by governments and when. The OxCGRT furnishes accessible data on 24 policy indicators, accompanied by a categorization of government responses into four groups: *i)* containment and closure policies (C)—i.e., schools and workplaces closure, cancel public events, restrictions on gathering, close public transport, stay at home requirement, restrictions on internal movement, and on international travel—, *ii)* economic policies (E)—i.e., income support, debt/contract relief for households, fiscal measures, and giving international support—, *iii)* health system policies (H)—i.e., public information campaign, testing policy, contact tracing, emergency investment in healthcare, investment in COVID-19 vaccines, facial coverings, vaccination policy, and protection of elderly people—, and *iv)* vaccination policies (V)—i.e., vaccine prioritization, vaccine eligibility/availability, vaccine financial support, vaccine requirement/mandate, and many other indices. They also provide four indices that aggregate the information into a single number with each index reporting a number between 0 and 100, reflecting the level of government response across specific dimensions: *i)* overall government response index (includes all indicators, i.e. C, E, H, and V), *ii)* containment and health index (includes all C and H indicators), *iii)* stringency index (includes all C indicators, plus H1, which records public information campaigns), and *iv)* economic support index (includes all E indicators). In this study, we used the whole dataset on restrictions, focusing in some cases on the stringency index.

Using anonymized data from apps like Google Maps, Google’s COVID-19 Community Mobility Reports [32] provides a dataset showing the percentage of changes in people’s mobility throughout the pandemic. This dataset measures daily visits to specific locations (e.g., workplaces, homes, grocery stores, parks, transit stations) and compares them to baseline days preceding the outbreak, introducing a significant bias. This baseline represents typical values calculated as the median over the five weeks from January 3 to February 6, 2020, accounting for routine differences between weekdays and weekends. In particular, we used data on workplaces (time spent at work), retail and recreation (includes places like restaurants, cafes, shopping centers, theme parks, museums, libraries, and movie theaters), grocery and pharmacy stores (includes places like grocery markets, food warehouses, farmers markets, specialty food shops, drug stores, and pharmacies), transit stations (includes public transport hubs such as subway, bus, and train stations) and residential (time spent at home). We confirmed a strong correlation between Facebook and Google mobility data (see Section 3.3 for details).

### 3.2 Pre-processing Data

In this section, we describe the data pre-processing steps taken to clean, transform, and prepare the raw data for effective analysis and modeling. In particular:

1. We normalized in [0, 1] the mobility variables considered from Facebook data.
2. We normalized the stringency index and the mobility variables considered from Google data.
3. We extracted the infection rates of COVID-19—similar to R_t_—using Sybil [6], a machine learning and variant-aware compartmental model framework designed to enhance prediction accuracy and explainability.

The first step involves the pre-processing of the considered UMD Global CTIS’s survey questions—we took the number of people that answered *Yes* to the first six questions (see Section 3.1), divided by the daily number of responses, obtaining values in [0, 1]—to represent different mobility categories: *i) C0 1* as *Workplaces* mobility, *ii) C0 2* as *Grocery* mobility, *iii) C0 3 + C0 5* as *Recreation* mobility, *iv) 1 - C0 4* as the complement of *Residential* mobility—we took the complement to ensure uniformity in the direction of all mobility trajectories—, and *v) C0 6* as *Stations* mobility.

Moreover, we processed question *C5* in the following way: for each answer, there are five possible binary options after applying one-hot encoding^1^. Specifically, *i) ATT* indicates that the person used a mask all the time, *ii) MOTT* most of the time, *iii) SOTT* some of the time, *iv) ALOTT* a little of the time, and *v) NOTT* none of the time. We computed a weighted average—normalized in the range [0, 1]—using the following fraction:

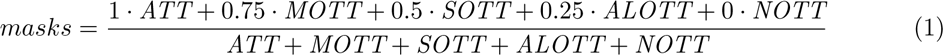

In particular, for each option, we applied a decreasing weight to the values, reflecting how frequently individuals used masks in public. Higher values indicate a greater proportion of people who consistently wore masks in public over the past week. We then divided the weighted sum by the total number of responses, resulting in values within the range [0, 1] that estimate the proportion of people who wore a mask in the past seven days. This variable was named *Masks*.

In the second step, we normalized the Google mobility variables—*Workplaces*, *Stations*, *Recreation*, *Residential*, and *Grocery* —and the *Stringency* index by dividing them by 100, as both are expressed as percentages. Google mobility data ranges from −1 to N, where −1 indicates no mobility compared to the baseline, and N represents the total population increase. The maximum possible increase occurs when only one person moves at time *t*, followed by the entire population moving at time *t + 1*. Conversely, the maximum possible decrease occurs when some individuals move at time *t*, followed by no one moving at time *t + 1*. However, these extreme scenarios do not appear in the data. As illustrated in Figure 1, the five Google mobility variables we analyzed range be-tween −0.75 and 1. Using Google mobility data, we were unable to compute the complement of *Residential* mobility because the values are relative to the baseline introduced above.

**Figure 1:**
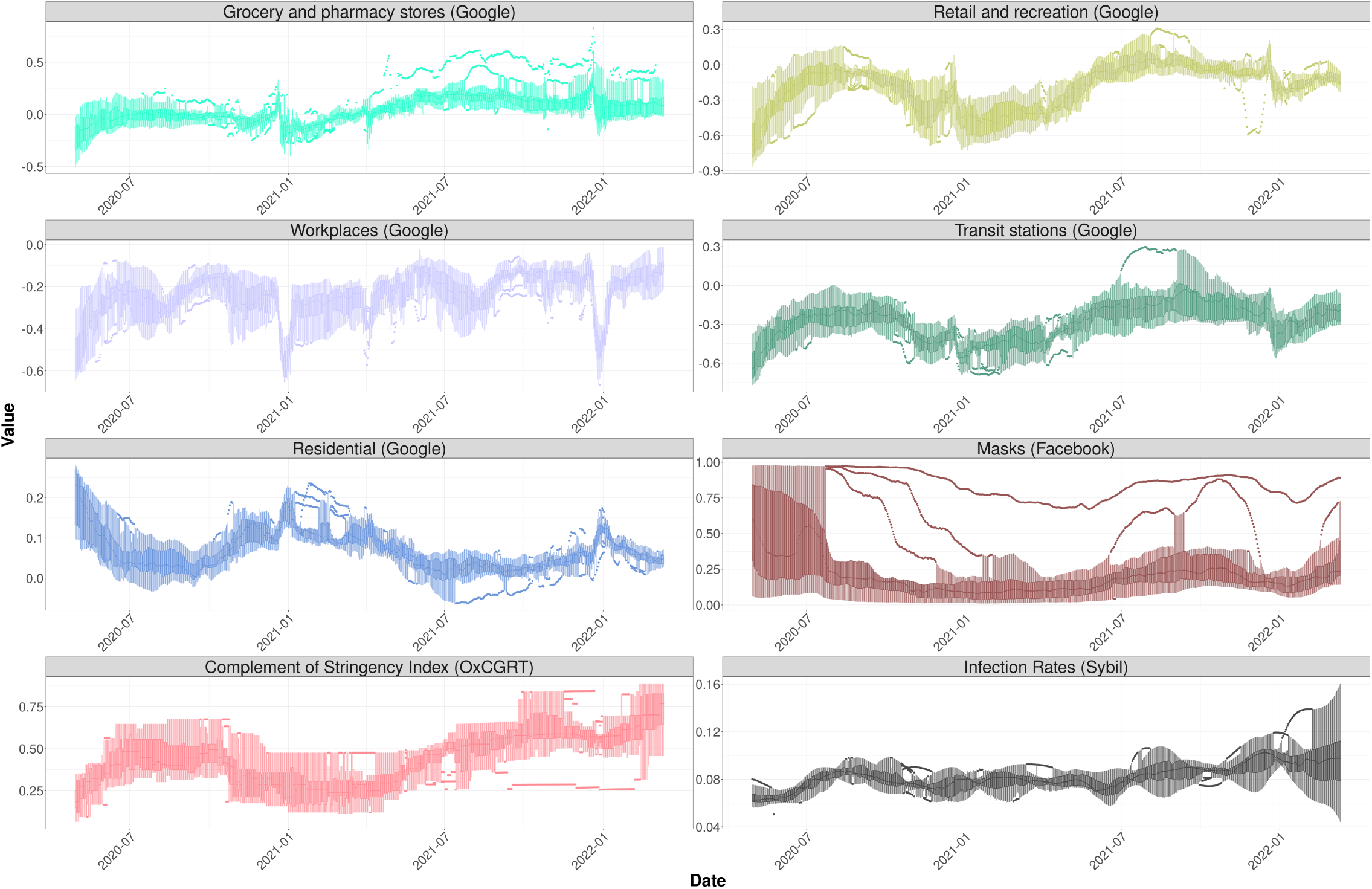
Temporal evolution of each variable—Google mobility variables (*Workplaces*, *Stations*, *Recreation*, *Residential*, and *Grocery*), the Facebook *Masks* variable, the complement of the OxCGRT *Stringency* index, and *Infection Rates* from Sybil—displayed as daily boxplots summarizing data across all considered countries from April 2020 to March 2022. For each variable, the median is displayed along with two bands: the darker band represents data from the first quartile (Q1) to the third quartile (Q3), while the lighter band includes data from *Q*1 *−* 1.5 *· IQR* to *Q*3 + 1.5 *· IQR*, where IQR stay for inter-quartile range. In addition, outliers can be easily identified.

In the third and last step, we extracted the *Infection Rates*—similar to R_t_—using Sybil [6], an integrated machine learning and variant-aware compartmental model framework designed to enhance prediction accuracy and explainability. Sybil leverages the relative stability of disease characteristic indices to make future projections and utilizes a straightforward, widely recognized analytical model to outline the infection dynamics. In particular, Sybil combines a simple compartmental model—in this case, a Susceptible - Infected - Recovered - Deceased - Susceptible (SIRDS) compartmental model with reinfections—with a machine learning-based predictive model—Prophet [44]—to forecast the future progression of infection, even in the presence of multiple virus strains. At its core, Sybil features an analytical model with dual functionality. In the first stage of Sybil’s operation, this analytical model derives critical parameter values from the data. These parameter values, particularly the reproduction number over time, R_t_, are then used as training data for Sybil’s ML component. Based on this training, the ML component predicts future values for the key parameters, which are subsequently fed back into the analytical model. Using these future parameter values, Sybil calculates the future evolution of daily infections using the analytical model.

We extended Sybil by *i)* including a pre-processing step using a technique—i.e., splines [19], piece-wise-defined mathematical functions that use multiple polynomial segments to create a smooth and continuous curve—to fill missing data and *ii)* adding the possibility to work with weekly data in case of unavailability of daily data—without significantly changing the information in the time series. In particular, we used an average number of days necessary to recover from the infection <u>^1^</u> equal to 14 days [13] and an average number of end-of-immunization days <u>^1^</u> equal to 180 days [48]. In this study, we focus solely on extracting infection rates from COVID-19 data, rather than modeling variants or using Sybil to make predictions. As mentioned in Section 3.1, for each country considered, we used official surveillance data [25, 24] on the cumulative number of deaths and the total population, as well as estimates of the fraction of active cases (COVID-like illness or CLI) [5, 38, 3, 39]—using a spline function to smooth these values.

Equation 2 shows the discretized version of the system of ordinary differential equations of the SIRDS compartmental model, with reinfections, used by Sybil. In this model, the population is divided into four categories: *i)* the Susceptible (S) population consists of healthy individuals who have not yet been infected by the disease, *ii)* the Infected (I) population comprises individuals who can transmit the disease to others, *iii)* the Recovered (R) population includes those who have recovered from the disease, and *iv)* the Deceased (D) population includes those who have died from it.

The model dynamics involve susceptible individuals becoming infected through contact with infected people. Infected individuals either recover and move to the recovered group or die of the disease, moving to the deceased group. Recovered people become again susceptible after 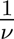 days on average. The model is governed by time-dependent rates—infection rates *β*(*t̃*), the recovery rates *γ*(*t̃*), and the fatality rates *λ*(*t̃*)—meaning that they may vary at each time step, with the time step corresponding to one day, with the only exception of the end-of-immunization rate *ν*.

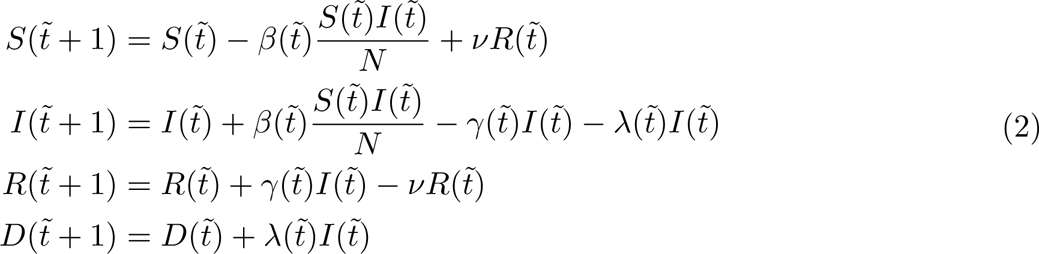

We primarily rely on Facebook data from April 29^th^, 2020 to May, 19^th^, 2021 for analyzing cultural patterns and clustering, as we focus on the first year of COVID-19, which saw a greater number of government-imposed restrictions. Additionally, we use Google data from April 2020 to March 2022 for XGBoost regression models, incorporating mask usage data from Facebook to enhance our analysis. We considered the following European countries: Finland, Norway, Sweden, Belgium, Ireland, the United Kingdom, France, Germany, Austria, Italy, Greece, Spain, and Portugal.

### 3.3 Preliminary Data Analysis

Figure 1 illustrates the temporal evolution of each variable from April 2020 to March 2022, including Google mobility variables—*Workplaces*, *Stations*, *Recreation*, *Residential*, and *Grocery* —, Facebook’s *Masks* usage, the complement of the OxCGRT *Stringency* index, and *Infection Rates* from Sybil. For each day, a boxplot aggregates values across the considered countries, showing the variability, trends, and seasonality over time. A clear and consistent correlation, whether positive or negative, can be observed across some of these variables. For example, a decrease in mobility at *Recreation* and *Stations* tends to correspond with a decrease in the complement of the *Stringency* index, reflecting the impact of stricter government-imposed restrictions aimed at reducing mobility. This suggests that as policies become more stringent, people are less likely to engage in activities that require them to travel or gather in public spaces. On the other hand, *Residential* mobility generally exhibits a negative correlation with these variables, indicating that as mobility restrictions increase, individuals tend to spend more time at home, possibly due to enforced lock-downs or heightened fear of exposure.

Similarly, the complement of the *Stringency* index appears to follow changes in *Infection Rates*, suggesting that reductions in NPIs may align with increases in *Infection Rates*, whereas stricter measures may coincide with a decline in *Infection Rates*. Lastly, *Grocery* and *Workplaces* mobilities appear to show weaker correlations with the other variables, indicating that changes in these particular types of mobility are less directly influenced by the complement of the *Stringency* index or *Infection Rates*. This may be due to the essential nature of grocery shopping and workplace attendance—strongly influenced by seasonality—, which might be less affected by the broader social distancing measures.

To address the first point outlined in Section 1.1 we utilized the Spearman [43] correlation index. Table 1 shows the correlation between the considered variables and averaged over all the selected countries. The complement of the *Stringency* index has moderately high correlations with most mobility variables, indicating that stricter policies tend to decrease people’s mobility. Generally, there is quite a high correlation among the mobility variables—including the Facebook *Masks* variable, except for *Grocery* and *Workplaces* where the correlation is lower. *Infection rates* have weaker correlations with most variables, showing a slightly higher correlation with the complement of the *Stringency* index (0.48). This suggests that fluctuations in *Infection Rates* correspond to changes in the complement of the *Stringency* index and are affected to a lesser extent by variations in mobility. The correlation is not particularly strong due to the delay between an increase in mobility and the subsequent rise in *Infection Rates*, as well as the lag between stricter restrictions and a decline in *Infection Rates*. In contrast, this delay is absent among mobility variables, and the lag between mobility and the complement of the *Stringency* index remains minimal.

**Table 1:** Spearman [43] correlation among selected variables—including Google mobility indicators (*Workplaces*, *Stations*, *Recreation*, *Residential*, and *Grocery*), the Facebook *Masks* variable, the complement of the OxCGRT *Stringency* index, and *Infection Rates* from Sybil—averaged across all considered countries.

|  | *Compl. of stringency index* | *Grocery* | *Recreation* | *Workplaces* | *Stations* | *Residential* | *Masks* | *Inf. rates* |
| --- | --- | --- | --- | --- | --- | --- | --- | --- |
| *Compl. of stringency index* | 1 | 0.52 | 0.69 | 0.52 | 0.68 | -0.64 | 0.52 | 0.48 |
| *Grocery* | 0.52 | 1 | 0.80 | 0.50 | 0.76 | -0.69 | 0.33 | 0.31 |
| *Recreation* | 0.69 | 0.80 | 1 | 0.48 | 0.91 | -0.89 | 0.54 | 0.39 |
| *Workplaces* | 0.52 | 0.50 | 0.48 | 1 | 0.61 | -0.60 | 0.26 | 0.28 |
| *Stations* | 0.68 | 0.76 | 0.91 | 0.61 | 1 | -0.92 | 0.59 | 0.37 |
| *Residential* | -0.64 | -0.69 | -0.89 | -0.60 | -0.92 | 1 | -0.54 | -0.32 |
| *Masks* | 0.52 | 0.33 | 0.54 | 0.26 | 0.59 | -0.54 | 1 | 0.12 |
| *Inf. rates* | 0.48 | 0.31 | 0.39 | 0.28 | 0.37 | -0.32 | 0.12 | 1 |

Finally, we compute the Spearman correlation index among Google and Facebook mobility variables, averaged across all chosen countries, in the period from April 29^th^, 2020 to May, 19^th^, 2021—we could not consider the whole period due to questions change (refer to Section 3.1 for more details). In particular, we obtained the following values, concluding that the correlation is high, especially for *Recreation*, *Residential*, and *Stations*: 0.70 for *Grocery*, 0.95 for *Recreation*, 0.75 for *Workplaces*, 0.83 for *Stations*, and −0.83 for *Residential* —data on mask usage is only available in the Facebook dataset. In the ‘Supplementary Material’ we included more figures illustrating the time evolution of each considered variable in detail, for each country.

## 4 Methods

Figure 2 presents a schematic overview of our approach. Starting with the datasets described in Section 3.1, we *pre-processed the data* (Section 3.2) to generate cleaned, filtered, and normalized mobility variables: *Workplaces*, *Stations*, *Recreation*, *Residential*, and *Grocery* (sourced from both Google and Facebook) and *Masks* (from Facebook). Additionally, we *incorporated restriction indices* and *Infection Rates* derived using Sybil [6]. Next, we *calculated three key ratios*, detailed later, based on average Facebook mobility (excluding masks), the stringency index, and infection rates. These ratios allowed us to identify cultural patterns by examining how official restrictions and self-imposed limitations (in the absence of formal regulations) influenced mobility and R_t_ across the studied countries (Section 5.1). Next, we *applied clustering techniques*, specifically K-means [27] and CONNECTOR [35], to analyze differences in social compliance with restrictions between Northern and Southern European countries, also highlighting no disparities in social interactions (Section 5.2). Finally, we *employed XGBoost regression models* [16] to estimate missing mobility data from restriction indicators and infer missing R_t_ values based on mobility patterns (Section 5.3).

**Figure 2:**
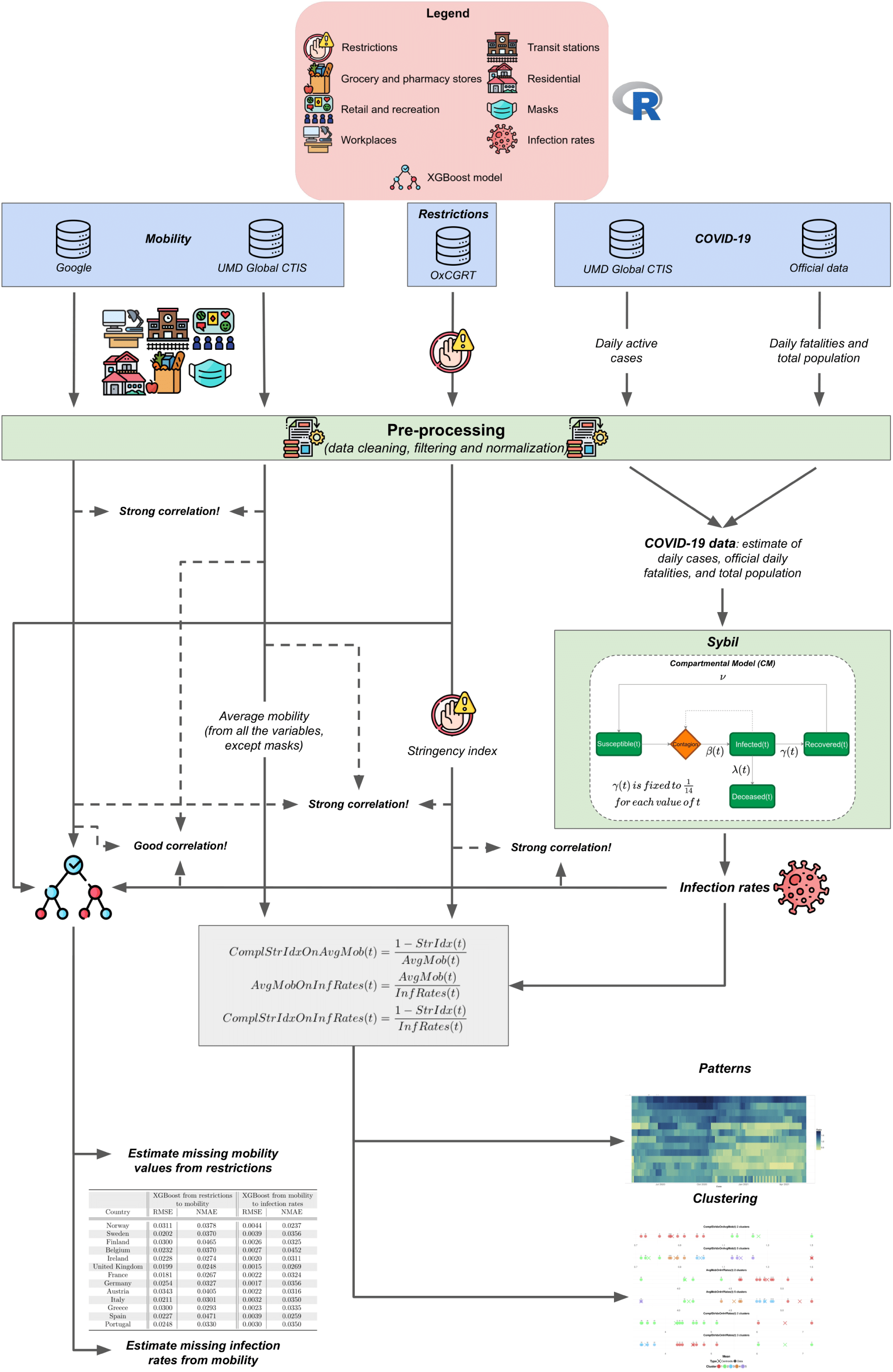
Schematic representation of our study.

XGBoost [16] is a powerful machine learning algorithm known for its high accuracy, particularly in classification and regression tasks. It efficiently handles missing data and uses regularization (L1 and L2) to reduce overfitting, making it robust in complex scenarios. Additionally, it is fast and scalable, leveraging parallel processing to handle large datasets, and provides clear feature importance for interpretability. However, XGBoost can be computationally intensive, requiring significant memory and processing power for large datasets. It also needs careful hyperparameter tuning for optimal performance and may overfit noisy data if not properly regularized. Despite its complexity, it offers strong results when properly configured. XGBoost is an advanced implementation of the gradient boosting algorithm designed for efficiency and performance, which builds an ensemble of decision trees sequentially to optimize predictive accuracy. By focusing on minimizing errors of previous trees through gradient descent and incorporating regularization techniques to prevent over-fitting, XGBoost achieves high accuracy and scalability.

On the other hand, K-means [27] is a popular clustering (unsupervised machine learning) algorithm to cluster similar data points into a predefined number of clusters. It is simple, fast, and efficient, especially for large datasets. The algorithm works by iteratively assigning data points to the nearest cluster center and then recalculating the centers until convergence. However, K-means has some limitations. It assumes clusters are spherical and evenly sized, which may not always be the case in real-world data. It is also sensitive to the initial selection of cluster centers, which can affect the final result. Additionally, K-means struggle with identifying complex or non-convex cluster shapes and can be influenced by outliers. To assess the quality of clustering, the Silhouette score [40] is often used. This score measures how similar a data point is to its own cluster compared to other clusters, with a range from −1 to 1. A higher score indicates better-defined clusters, while values near 0 or negative suggest poor clustering or potential misclassification.

Finally, CONNECTOR [35] is an R package for unsupervised longitudinal data analysis, using a model-based approach to cluster functional data, particularly effective for sparse and irregular curves. It employs spline functions to create smooth curves for accurate modeling and clustering. The package provides multiple visual inspections of the data, specific indexes, and graphics to set the two model’s free parameters (i.e., the dimension of the spline basis vector and the optimal number of clusters). Finally, different visualization plots might be exploited to show the longitudinal clustering result and cluster stability.

Concretely, we used Facebook mobility data in the first period—from April 29^th^, 2020 to May, 19^th^, 2021—to address the second and third points outlined in Section 1.1. We selected this initial period because the pandemic’s onset saw stricter restrictions in place, and individuals were generally more compliant with these measures. Additionally, we computed the following ratios for each country to identify cultural patterns:

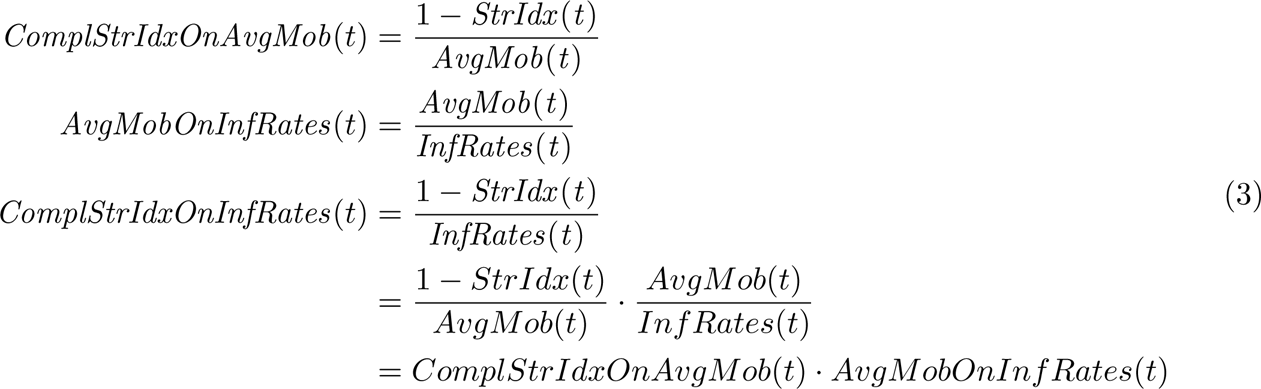

where *AvgMob(t)* is the average mobility computed on the considered variables—*Workplaces*, *Stations*, *Recreation*, *Residential*, and *Grocery* —, without considering *Masks*, at time *t*; *InfRates(t)* is the *Infection Rates* at time *t*; and *StrIdx(t)* is the *Stringency* index at time *t*. *AvgMob(t)* and *InfRates(t)* are always greater than 0. Based on these time series, we identified patterns and applied clustering techniques using K-means [27] and CONNECTOR [35]—the results of the latter are in the ‘Supplementary Material’.

Specifically, the *ComplStrIdxOnAvgMob(t)* ratio emphasizes the relationship between restrictions and mobility, primarily reflecting aspects of social compliance with restrictions. The *AvgMobOnInfRates(t)* ratio illustrates the connection between mobility and infection rates, mainly capturing factors related to social interactions. Finally, the *ComplStrIdxOnInfRates(t)* ratio highlights the relationship between restrictions and infection rates, influenced by both social compliance with restrictions and social interactions.

To address the last point outlined in Section 1.1, we employed XGBoost regression models [16]. Specifically, we used 5-fold cross-validation (80% training and 20% testing) to train XGBoost models for each country. This approach can be viewed as estimating missing data, where the 20% of discarded data represents unobserved values used for model testing, while the models aim to estimate the missing values. These models were designed to estimate mobility variables—namely, *Workplaces*, *Stations*, *Recreation*, *Residential*, and *Grocery* (from Google), as well as *Masks* (from Facebook)—at time *t*, using the whole restriction indices from OxCGRT at the same time point. Additionally, we trained a separate XGBoost model to estimate *Infection Rates* (extracted from Sybil) at time *t*, using the mobility variables from that time.

## 5 Results

Here, we present the key results obtained using the methods outlined in Section 4. Specifically, Sections 5.1 and 5.2 detail the patterns and clusters identified by analyzing the ratios introduced in the previous section, while in Section 5.3, we discuss the results of the XGBoost regression models.

### 5.1 Patterns

Figure 3 illustrates the ratios calculated using Equation 3 for each considered country. Figure 3.*a*, showing the *ComplStrIdxOnAvgMob(t)* ratio, highlights the relationship between restrictions and mobility. Higher values of this ratio indicate that minimal restrictions (low stringency) are sufficient to limit mobility or keep people at home. This often reflects the influence of external factors such as fear of infection or strong social compliance with NPIs. As a result, fewer interventions are needed to achieve the same mobility reduction. Notably, countries like Norway, Sweden, Finland, and Belgium exhibit consistently high values, visible as dark bands in the heatmap.

**Figure 3:**
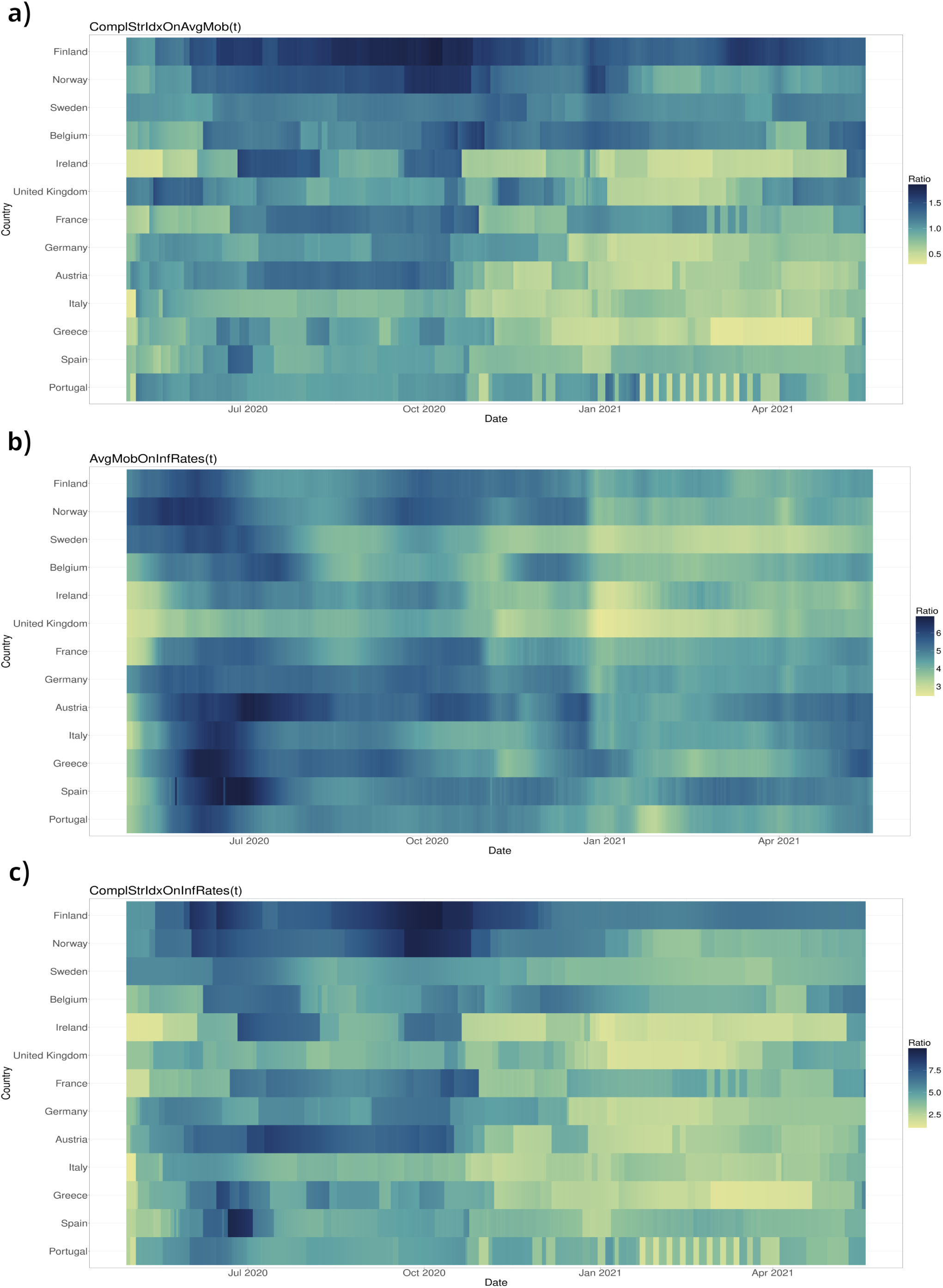
Heatmaps display country ratios: *ComplStrIdxOnAvgMob*, *AvgMobOnInfRates*, and *ComplStrIdxOnInfRates*, with darker colors for higher values.

Figure 3.*b*, which illustrates the *AvgMobOnInfRates(t)* ratio, illustrates the relationship between mobility and infection rates. Higher values indicate that average mobility is increasing faster than the infection rate, or that the infection rate is decreasing relative to mobility. For example, in June and July 2020, most countries showed higher values due to increased mobility and lower infection rates following the first pandemic wave. However, the United Kingdom was an exception, with mobility remaining lower while infection rates began to rise again. Two other notable points occurred around December 2020 (Christmas) and April 2021 (Easter), when seasonality played a crucial role. In December 2020, a sharp decline in mobility caused the ratio to drop—except in Greece, where the infection peak occurred later and the decline in mobility was slower. After this decline, the ratio increased as mobility gradually rose, though more slowly in the United Kingdom. In April 2021, a similar pattern emerged, but with a faster rise in mobility. In the ‘Supplementary Material,’ additional details on country-specific data are provided. In general, Figure 3.*b* reveals no significant distinctions among countries in the impact of mobility on infection rates, with few exceptions such as the United Kingdom, Ireland, and Sweden where lighter bands are noticeable, particularly after Christmas 2020. This suggests a broadly similar relationship across populations. The lack of clear differentiation indicates that, despite regional and cultural variations, mobility patterns and their influence on infection rates were largely consistent during the observed period, driven primarily by social interactions.

Figure 3.*c* shows the *ComplStrIdxOnInfRates(t)* ratio, which highlights the relationship between restrictions and infection rates. Higher values of this ratio signify that the complement of the stringency index is high (i.e., restrictions are minimal) while infection rates remain relatively low. This suggests scenarios where either limited restrictions are sufficient to manage the infection rate or where infection rates are inherently low, resulting in a higher ratio. In this case, multiple factors come into play, such as effective natural mitigation mechanisms, high societal compliance, the nature of social interactions, and other external influences that collectively help control the spread of infections despite minimal interventions. Additionally, Finland exhibits different patterns starting from the end of 2020, largely because restrictions remained unchanged for several months.

### 5.2 Clustering

In this section, we apply K-means clustering [27] to group countries based on the three ratios described in Section 4—in the ‘Supplementary Material,’ we also use CONNECTOR [35] for comparison. Specifically, we perform clustering using the average values of these ratios computed over the entire period for each country. Additionally, we explore clustering by incorporating both the average values and their corresponding standard deviations to assess the impact of variability.

Figure 4 shows the average Silhouette score [40], computed using K-means and evaluated with both the average alone and the combination of the average and standard deviation, for the three considered ratios. Specifically, the optimal number of clusters appears to be two in all cases. However, when considering the average, three clusters for the *ComplStrIdxOnInf Rates* ratio also seem to be a reasonable choice, while five clusters are appropriate for the *AvgMobOnInf Rates* and *ComplStrIdxOnAvgMob* ratios. Additionally, when considering both the average and standard deviation, three clusters also appear to be a reasonable choice for the *ComplStrIdxOnAvgMob* ratio and six for the *AvgMobOnInf Rates* and *ComplStrIdxOnAvgMob* ratios.

**Figure 4:**
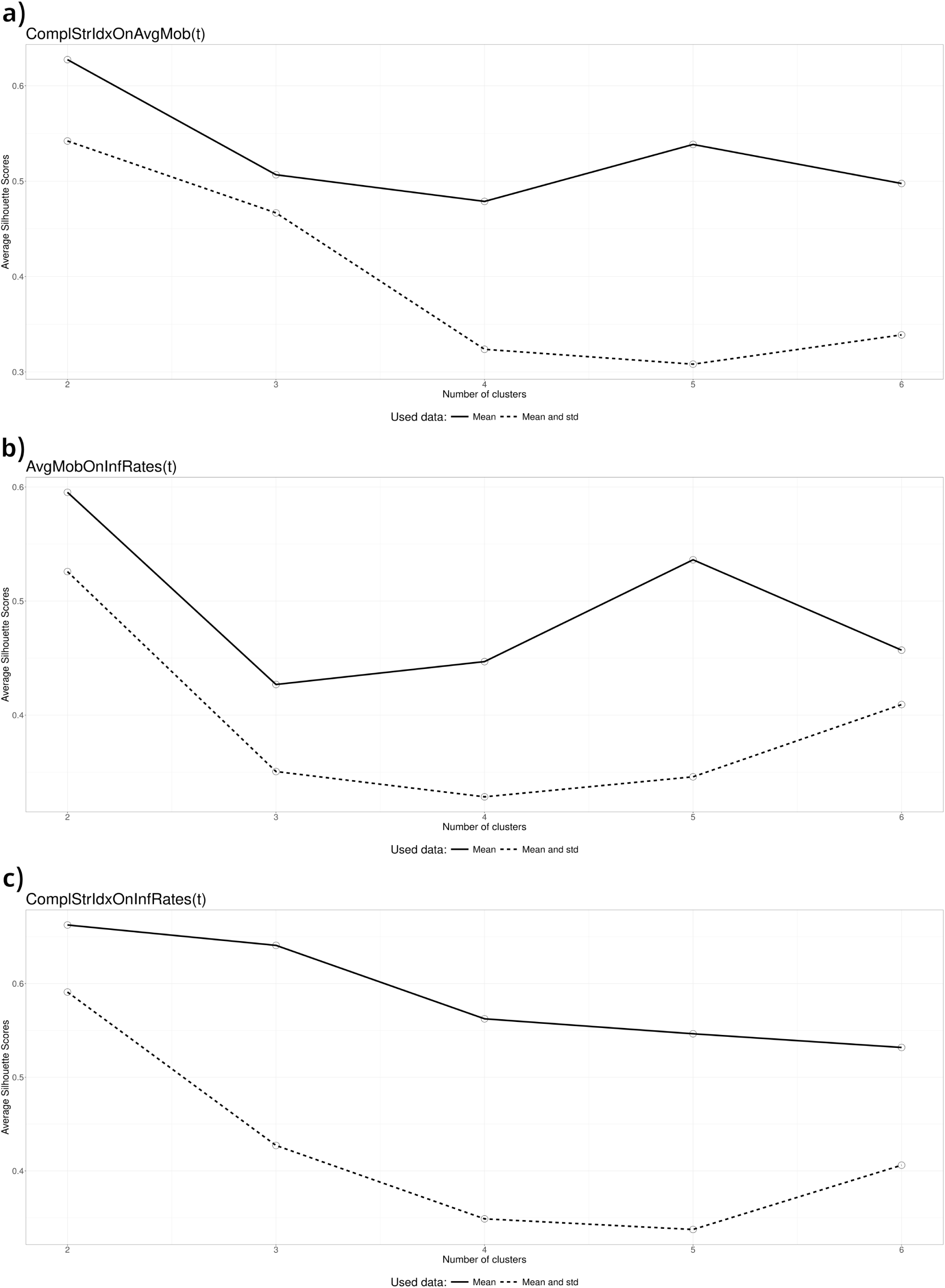
Silhouette score [40] computed using K-means—evaluated both with the average alone and with the combination of the average and standard deviation—for the three considered ratios: *AvgMobOnInf Rates*, *ComplStrIdxOnInf Rates*, and *ComplStrIdxOnAvgMob*.

Figure 5 illustrates the clusters derived using only the average values. Focusing on the *ComplStrIdxOnAvgMob* ratio, the first two plots show the results for two and five clusters, respectively. These clusters reveal a clear separation between Northern and Southern European countries. Notably, in both cases, the three analyzed Northern European countries—Finland, Norway, and Sweden—form a distinct cluster, separate from the other nations. When considering five clusters, Finland is placed in its own separate cluster. Additionally, in both cluster configurations, Belgium is grouped with the Northern countries, likely due to the fact that fewer restrictions were implemented there as well.

As noted in Section 5.1, in these countries, less stringent measures were effective in controlling infections, largely due to high levels of social compliance. In Northern European countries, cultural norms, behavioral tendencies, and strong trust in public health guidance reduce the need for coercive measures, as people are more likely to voluntarily adhere to recommendations. Moreover, we observe that countries like Italy, Greece, Spain, and Portugal—typically considered Southern European countries—cluster more to the left, alongside several intermediate European countries like Ireland, Germany, France, Austria, and the United Kingdom. This suggests that these countries needed more stringent restrictions to encourage people to stay at home. The three clusters on the left in the second plot reflect progressively increasing levels of restrictions, from right to left.

Considering the *AvgMobOnInf Rates* ratio—the fourth and fifth plots in Figure 5, which show two and five clusters, respectively—we observe no clear distinction between Northern and Southern countries. Instead, countries appear mixed, as noted in Section 5.1. This can be attributed to the fact that, when examining the effect of mobility on infection rates, we are primarily considering the role of social interactions, instead of social compliance. Unlike measures that target mobility alone, the influence of social interactions encompasses a wide range of factors, including cultural norms and community behaviors. In countries with higher social interactions, mobility may have less of an impact on infection rates, while in others, even small changes in mobility can lead to significant fluctuations in infections. This mix of factors contributes to the lack of a clear distinction and highlights the complexity of how mobility and social behaviors interact in different contexts.

**Figure 5:**
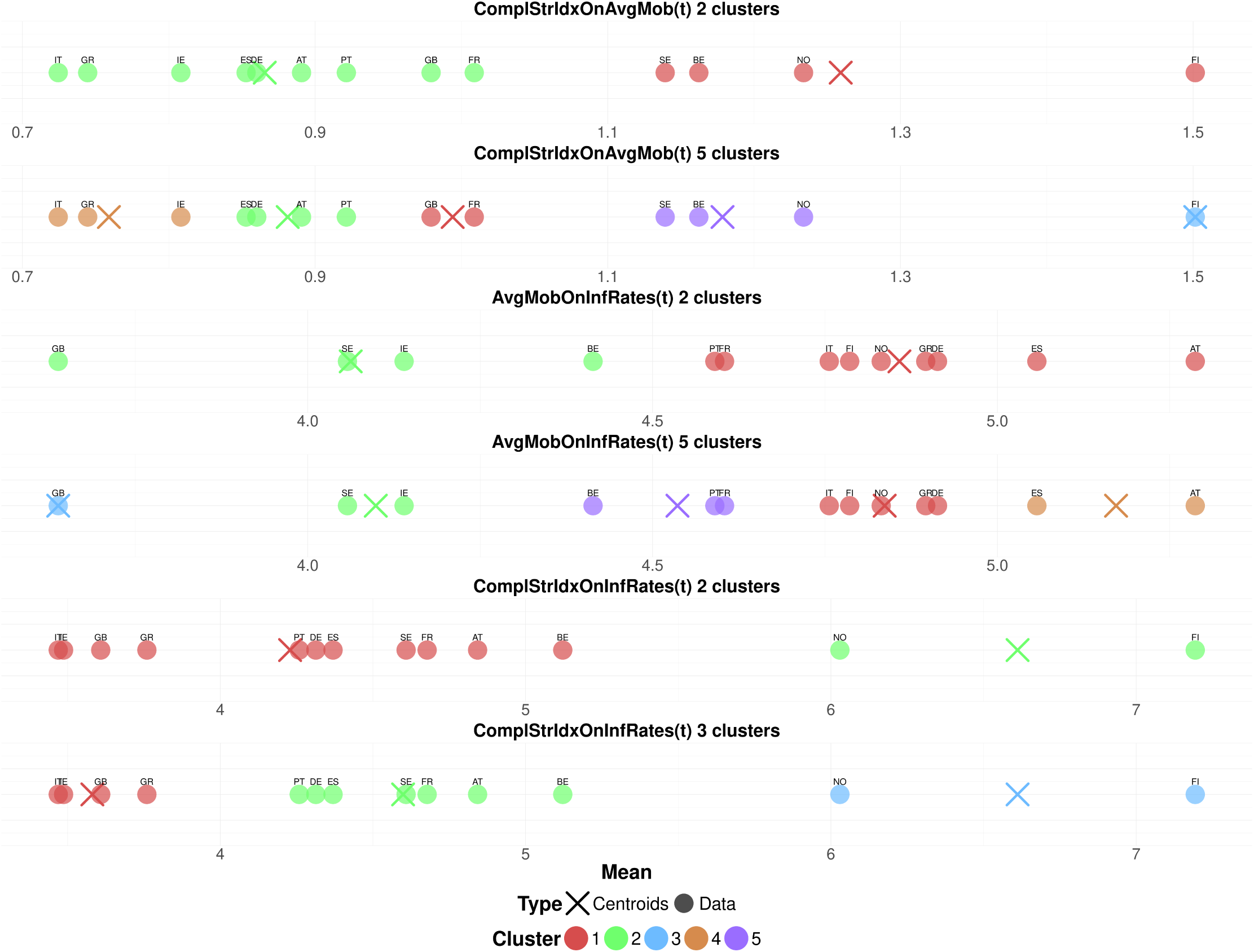
Clusters derived using the average values for the *ComplStrIdxOnAvgMob*, *AvgMobOnInf Rates*, and *ComplStrIdxOnInf Rates* ratios with K-means [27].

Finally, when examining the *ComplStrIdxOnInf Rates* ratio—the last two plots in Figure 5, which show two and three clusters, respectively—we observe that Finland and Norway remain distinctly separated, while Sweden does not. In this case, both social compliance and social interactions are considered, as we are directly linking restrictions to infection rates. This suggests that the clusters observed here are more similar to those seen in the first and second plots than to those in the third and fourth plots, as is indeed the case.

Figure 6 illustrates the clusters derived using both the average values and the standard devia-tion. These clusters align closely with those shown in Figure 5. For the *ComplStrIdxOnAvgMob* ratio with three clusters, a clear separation is observed among Finland and Norway, Belgium, and Sweden, consistent with the pattern noted in the second plot of Figure 5. For the *AvgMobOnInf Rates* ratio with six clusters, we observed the same general separation as with five clusters when considering only the average; however, the cluster containing Norway, Greece, Germany, Italy, and Finland is further split into two distinct clusters. Similarly, for the *ComplStrIdxOnAvgMob* ratio with six clusters, Finland, Norway, and Austria form three separate singleton clusters. Meanwhile, Ireland and Greece are grouped into a fourth cluster, Italy and the United Kingdom constitute a fifth cluster, and all remaining countries are included in the final cluster.

**Figure 6:**
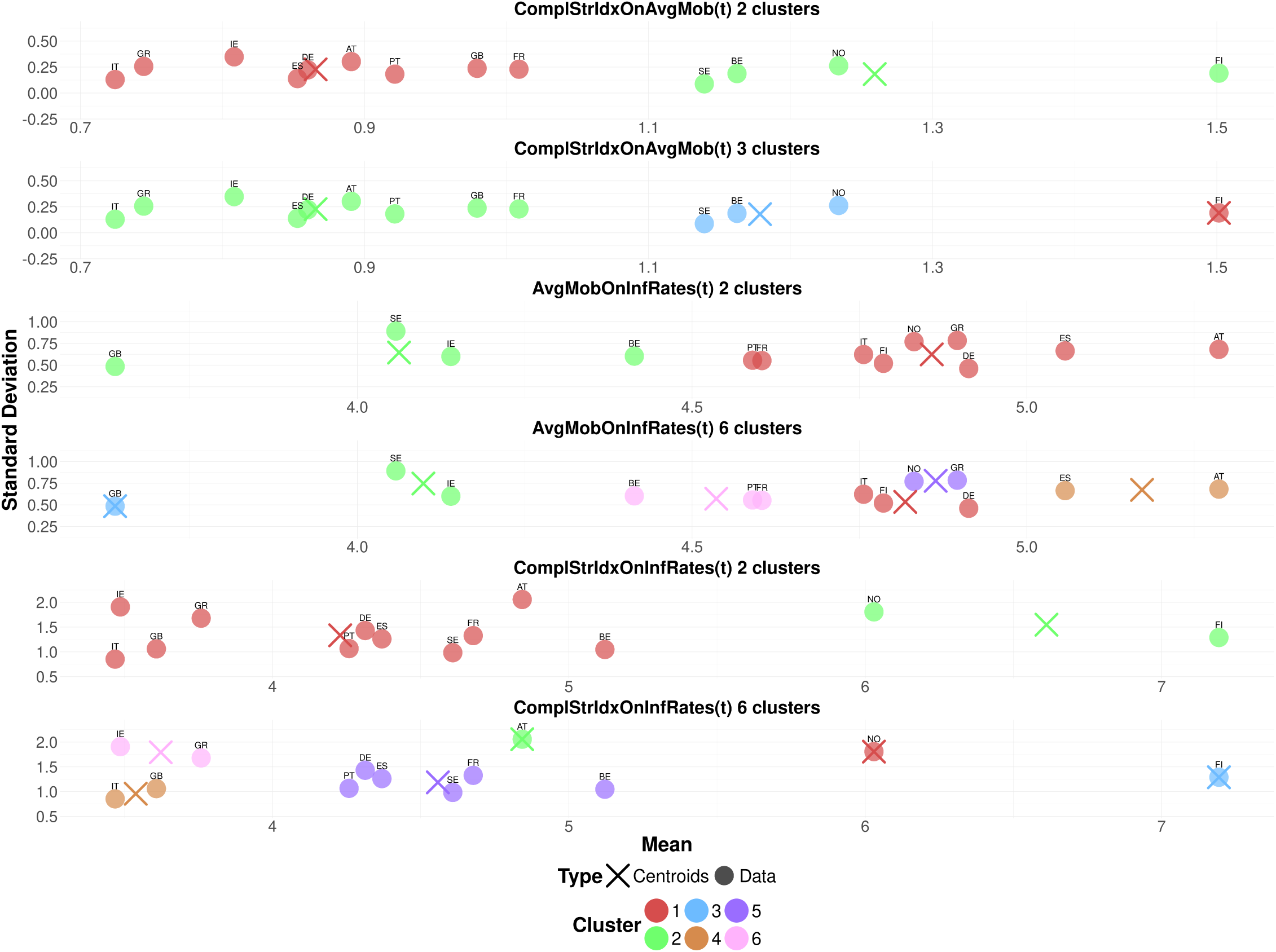
Clusters derived using both average values and standard deviations for the *ComplStrIdxOnAvgMob*, *AvgMobOnInf Rates*, and *ComplStrIdxOnInf Rates* ratios with K-means [27]. Colors may differ with respect to Figure 5.

To summarize, in Section 5.1 we identified patterns analyzing the interplay among restrictions, mobility, and infection rates, while in Section 5.2 we analyzed country clustering based on three key ratios using K-means [27], considering both average values and their standard deviations. Northern European countries, like Finland, Norway, and Sweden formed distinct clusters when social compliance plays a crucial role in the ratios, reflecting high compliance and effective infection control with fewer restrictions. Southern and intermediate European countries required stricter measures, highlighting differences in compliance levels. The *AvgMobOnInf Rates* ratio showed mixed clustering, driven by social interactions rather than compliance. The *ComplStrIdxOnInf Rates* ratio, linking restrictions to infection rates, revealed clusters similar to those based on mobility ratios, emphasizing the combined role of compliance and interactions. These findings underscore the importance of tailored strategies in managing public health measures, potentially by grouping countries into clusters, as we have identified.

### 5.3 Regression Models

Table 2 presents the average root mean squared error (RMSE) and normalized mean absolute error (NMAE) between the test set (ground truth, 20% of the data) and the corresponding estimates, averaged across the five considered folds, for the XGBoost [16] regression models. These models were used to estimate missing mobility data based on restrictions—we averaged the errors over the six mobility variables—and missing R_t_ data based on mobility patterns for each country. The reported errors highlight the models’ strong performance in accurately estimating missing values. In the ‘Supplementary Material’, we provide additional details.

**Table 2:**
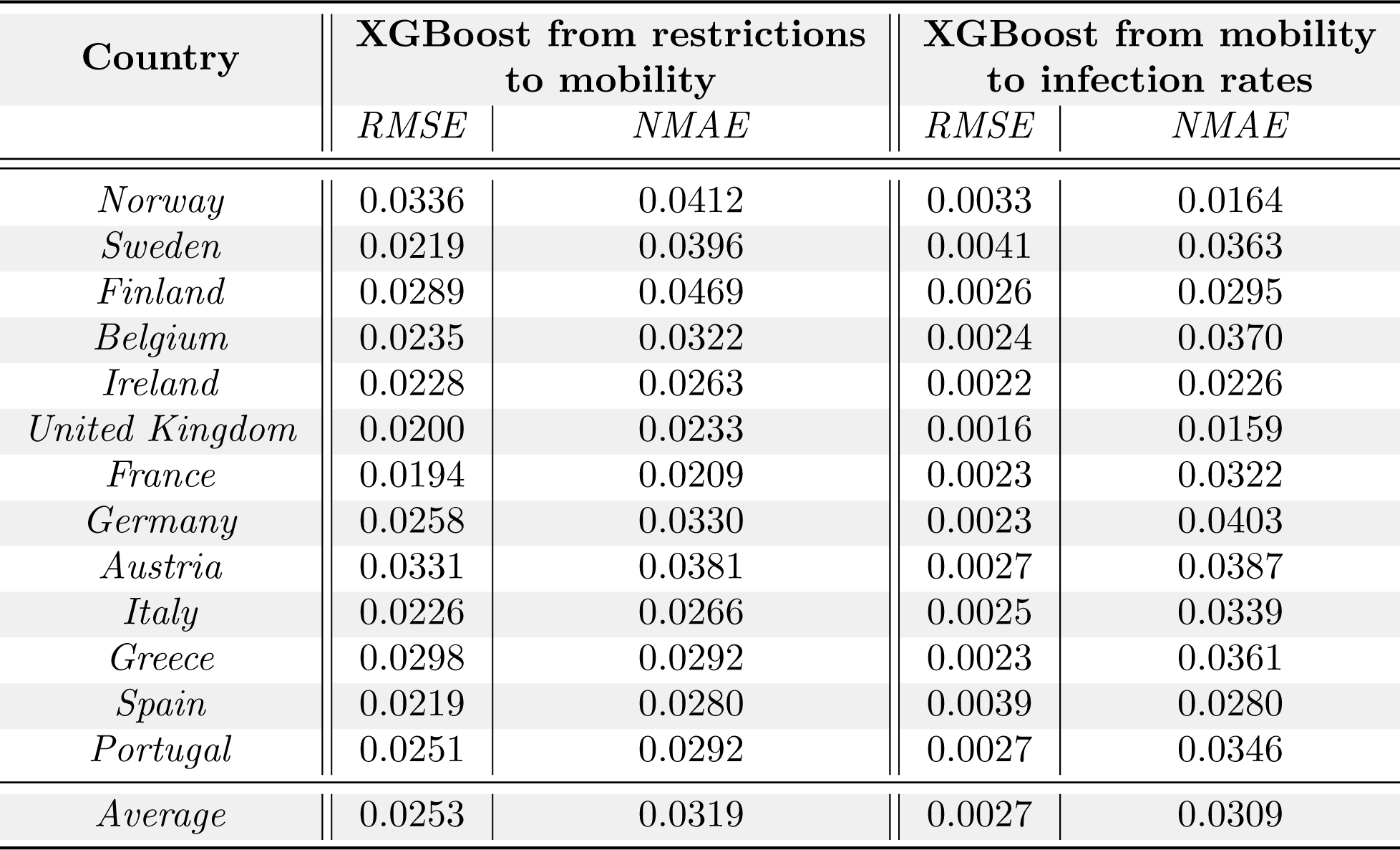
Average RMSE and NMAE across five folds for XGBoost models estimating missing mobility data from restrictions—the errors are averaged over the six variables—and missing R_t_ data from mobility patterns for each country.

The estimated values can be used to fill in gaps in time series data, allowing us to uncover patterns, simulate various scenarios, or test hypotheses regarding the relationships between variables. Furthermore, the completed time series can be utilized with other models, such as neural networks for forecasting, decision trees for classification, or reinforcement learning models for optimizing policy interventions.

## 6 Discussion

Table 3 shows a summary of key findings. Our analysis highlights a strong *correlation* between government stringency measures and mobility reductions, as evidenced by the high correlation values observed in Figure 1 and Table 1. This indicates that restrictions are highly effective in modifying public behavior, with an almost immediate response in mobility patterns, largely due to social compliance with public health measures. However, the relationship between mobility and R_t_ is weaker, suggesting a delayed and more complex interaction between changes in movement and its influence on infection rates. This lag can be attributed to additional factors, such as social interactions, incubation periods, pre-existing transmission dynamics, and the delay between an increase in mobility and the subsequent rise in R_t_.

**Table 3:** Summary of key findings.

|  | Findings |
| --- | --- |
| **Correlation** | A strong correlation is observed between the analyzed restrictions and mobility variables, in contrast to the weaker correlation identified between mobility and infection rates. |
| **Patterns** | Clear cultural patterns emerge when examining the impact of the stringency index on mobility and infection rates and the effect of mobility on infection rates. |
| **Clustering** | There is a clear separation between Northern and Southern European countries when social compliance is the key factor, but this distinction disappears when social interactions are the main focus. |
| **Estimations** | XGBoost models perform very well in estimating missing mobility data from restrictions and missing infection rates from mobility. |

Analyzing the ratios defined in Section 4, distinct *cultural patterns* emerge in the interplay between restrictions, mobility, and infection rates. These patterns enable us to cluster countries accordingly. The *clustering analysis* in Section 5.2 reveals a clear separation between Northern and Southern European countries when *social compliance* plays a major role. Northern European countries, such as Finland, Norway, and Sweden, demonstrated higher compliance with restrictions, leading to significant mobility reductions with less stringent measures. In contrast, Southern European countries required stricter measures to achieve similar levels of compliance. This distinction reflects cultural and societal differences, including public trust in authorities, social norms, and economic conditions. When *social interactions* become the dominant factor, the clustering distinction between Northern and Southern countries disappears. This finding underscores the universal role of interpersonal behavior in driving disease transmission, irrespective of regional or cultural differences. It highlights the importance of addressing social interaction patterns alongside mobility restrictions in designing effective public health strategies.

Finally, regression models enable us to accurately *estimate* missing mobility data from restrictions and missing infection rates from mobility. For instance, these estimated values can be used to complete time series data, which can then be employed to identify patterns in the data, simulate different scenarios, or test hypotheses about the relationships between variables. Additionally, the completed time series can be applied to other models, such as neural networks for forecasting, decision trees for classification tasks, or reinforcement learning models for optimizing policy interventions.

The findings have significant implications for policy-making in managing infectious diseases. First, the rapid and robust response of mobility to restrictions underscores the importance of timely and decisive government interventions. Policymakers should prioritize measures that effectively reduce high-risk mobility patterns, such as public gatherings and inter-regional travel. Second, the observed cultural and regional variations suggest the need for context-specific strategies. Northern European countries may benefit from less coercive measures, harnessing their higher levels of public trust and compliance. In contrast, Southern European countries may require more stringent interventions to achieve similar outcomes. Such tailored approaches can optimize the balance between restriction effectiveness and societal acceptance. Finally, the weaker correlation between mobility and R_t_ emphasizes the importance of complementing mobility-focused measures with other interventions, such as improved testing, contact tracing, and vaccination campaigns. Addressing factors beyond mobility can accelerate the reduction in transmission rates.

## 7 Conclusions

In this study, we utilized diverse data sources to investigate the relationship between government-imposed restrictions, mobility patterns, and the effective reproduction number R_t_ of COVID-19. Due to its unprecedented global impact and the extensive data collected during the pandemic, COVID-19 served as an ideal case study, offering valuable insights into the dynamics of infectious disease control in a highly interconnected world.

We focused on several European countries—specifically Finland, Norway, Sweden, Belgium, Ireland, the United Kingdom, France, Germany, Austria, Italy, Greece, Spain, and Portugal— to identify cultural patterns and cluster these nations using two state-of-the-art approaches: K-means [27] and CONNECTOR [35]. Moreover, we employed XGBoost [16] to estimate missing mobility data from the whole restriction dataset and to infer missing R_t_ values from mobility data—an innovative approach that represents a novel contribution to the existing literature.

Our findings emphasize the importance of tailoring public health strategies to account for cultural and regional differences, particularly in compliance with NPIs. Moreover, the methodological approach adopted in this study—combining machine learning techniques with epidemiological insights—provides a robust framework for analyzing mobility and infection dynamics in future pandemics.

Despite the valuable insights provided by this study, several limitations should be noted. First, while the focus on European countries offers important cross-cultural comparisons, the findings may not be fully generalizable to regions with different socio-cultural or economic contexts. Second, we trained separate XGBoost models for each country, which limits the generalization of the models. Additionally, due to issues with changing survey questions on platforms like Facebook, the study primarily focuses on the initial phase of the COVID-19 pandemic and may not fully account for evolving pandemic dynamics, such as the emergence of new variants, which could impact the relationship between mobility patterns and infection rates. Finally, we attempted to predict future mobility trends based on restrictions and future R_t_ values based on mobility, achieving relatively good results. However, other time-series techniques, such as Prophet [44], could give more accurate predictions.

Future work may include: *i)* expanding the analysis to include additional countries and regions, particularly those with differing socio-cultural and economic contexts, would enhance the generalization of the results, *ii)* training general XGBoost models to estimate mobility from restrictions and infection rates from mobility data, and *iii)* extending the current approach to other infectious diseases, public health scenarios, or more broadly, to different contexts. This would provide a deeper understanding of how easily observable variables can influence more challenging-to-observe variables.

## Data Availability

All datasets used in this work are open-source and publicly available. In particular, we used official surveillance data from the COVID-19 Data Hub, mobility data from UMD Global CTIS's survey, estimates of active cases computed from the UMD Global CTIS's survey, information on restrictions from the Oxford Covid-19, and Google mobility data. The UMD Global CTIS's survey data (in aggregated form) are openly accessible at \url{https://github.com/GCGImdea/coronasurveys/}. The microdata of the CTIS survey from which the aggregated data was obtained cannot be shared, as per the Data Use Agreements signed with Facebook, Carnegie Mellon University (CMU), and the University of Maryland (UMD). Thus, the authors do not have permission to share the microdata of the CTIS survey. The code is available at \url{https://github.com/daniele-baccega/mobility-study}.

https://github.com/GCGImdea/coronasurveys/

https://github.com/OxCGRT/covid-policy-dataset

https://covid19datahub.io/

https://ourworldindata.org/covid-mobility-trends

## 8 Ethical declaration

The Ethics Board (IRB) of IMDEA Networks Institute gave ethical approval for this work on 2021/07/05. IMDEA Networks has signed Data Use Agreements with Facebook, Carnegie Mellon University (CMU) and the University of Maryland (UMD) to access their data, specifically UMD project 1587016-3 entitled C-SPEC: Symptom Survey: COVID-19 and CMU project STUDY2020 00000162 entitled ILI Community-Surveillance Study. The data used in this study was collected by the University of Maryland through The University of Maryland Social Data Science Center Global COVID-19 Trends and Impact Survey in partnership with Facebook. Informed consent has been obtained from all participants in this survey by this institution. All the methods in this study have been carried out in accordance with relevant of ethics and privacy guidelines and regulations.

## 9 Availability of materials and data

All datasets used in this work are open-source and publicly available. In particular, we used official surveillance data from the COVID-19 Data Hub [25, 24], mobility data from UMD Global CTIS’s survey [5, 21, 30], estimates of active cases computed from the UMD Global CTIS’s survey [5, 38, 3, 39], information on restrictions from the Oxford Covid-19, and Google mobility data [32]. The UMD Global CTIS’s survey data (in aggregated form) are openly accessible at https://github.com/GCGImdea/coronasurveys/. The microdata of the CTIS survey from which the aggregated data was obtained cannot be shared, as per the Data Use Agreements signed with Facebook, Carnegie Mellon University (CMU), and the University of Maryland (UMD). Thus, the authors do not have permission to share the microdata of the CTIS survey. The code is available at https://github.com/daniele-baccega/mobility-study.

## 12 Acknowledgement

D.B. is a Ph.D. student enrolled in the National Ph.D. in Artificial Intelligence, XXXVII cycle, health and life sciences course organized by Università Campus Bio-Medico di Roma.

## Footnotes

1 The possible answers are: *All the time, Most of the time, Some of the time, A little of the time, and None of the time*. For example, if a person answered *Some of the time*, we assigned the following vector to that response: [0, 0, 1, 0, 0]. By applying this approach to all responses and summing the vectors, we can determine the number of people who selected each specific answer.

## References

[1] Michael O Adeniyi et al. “Assessing the impact of public compliance on the use of non-pharmaceutical intervention with cost-effectiveness analysis on the transmission dynamics of COVID-19: Insight from mathematical modeling”. In: Modeling, control and drug development for COVID-19 outbreak prevention (2022), pp. 579–618. doi: 10.1007/978-3-030-72834-2_17.

[2] Laura Alessandretti. “What human mobility data tell us about COVID-19 spread”. In: Nature Reviews Physics 4.1 (2022), pp. 12–13. doi: 10.1038/s42254-021-00407-1.

[3] Javier Álvarez, et al. “Estimating active cases of COVID-19”. In: MedRxiv (2021), pp. 2021–12. doi: 10.48550/arXiv.2108.03284.

[4] Nikolaos Askitas, Konstantinos Tatsiramos, and Bertrand Verheyden. “Estimating worldwide effects of non-pharmaceutical interventions on COVID-19 incidence and population mobility patterns using a multiple-event study”. In: Scientific reports 11.1 (2021), p. 1972. doi: 10.1038/s41598-021-81442-x.

[5] Christina M Astley et al. “Global monitoring of the impact of the COVID-19 pandemic through online surveys sampled from the Facebook user base”. In: Proceedings of the National Academy of Sciences 118.51 (2021), e2111455118. doi: 10.1073/pnas.2111455118.

[6] Daniele Baccega et al. “Enhancing COVID-19 forecasting precision through the integration of compartmental models, machine learning and variants”. In: Scientific Reports 14.1 (2024), p. 19220. doi: 10.1038/s41598-024-69660-5.

[7] Hamada S Badr et al. “Association between mobility patterns and COVID-19 transmission in the USA: a mathematical modelling study”. In: The Lancet Infectious Diseases 20.11 (2020), pp. 1247–1254. doi: 10.1016/S1473-3099(20)30553-3.

[8] D Banerjee, A Campbell, J Gruger, et al. “What has the pandemic revealed about the US health care system—and what needs to change”. In: MIT News (2021). url: https://news.mit.edu/2021/what-has-pandemic-revealed-about-us-health-care-what-needs-change-0405#:∼:text=The%20spread%20of%20the%20virus,the%20labor%20of%20underpaid%20health (visited on 12/23/2024).

[9] Nicolas Banholzer et al. “Estimating the effects of non-pharmaceutical interventions on the number of new infections with COVID-19 during the first epidemic wave”. In: PLOS ONE 16.6 (June 2021), pp. 1–16. doi: 10.1371/journal.pone.0252827. url: https://doi.org/10.1371/journal.pone.0252827.

[10] Vesna Barros et al. “A causal inference approach for estimating effects of non-pharmaceutical interventions during Covid-19 pandemic”. In: Plos one 17.9 (2022), e0265289. doi: 10.1371/journal.pone.0265289.

[11] Francisco Benita. “Human mobility behavior in COVID-19: A systematic literature review and bibliometric analysis”. In: Sustainable Cities and Society 70 (2021), p. 102916. doi: 10.1016/j.scs.2021.102916.

[12] Patrick Bryant and Arne Elofsson. “Estimating the impact of mobility patterns on COVID-19 infection rates in 11 European countries”. In: PeerJ 8 (2020), e9879. doi: 10.7717/peerj.9879.

[13] Andrew William Byrne et al. “Inferred duration of infectious period of SARS-CoV-2: rapid scoping review and analysis of available evidence for asymptomatic and symptomatic COVID-19 cases”. In: BMJ open 10.8 (2020), e039856. doi: 10.1136/bmjopen-2020-039856.

[14] Armando Cartenì, Luigi Di Francesco, and Maria Martino. “How mobility habits influenced the spread of the COVID-19 pandemic: Results from the Italian case study”. In: Science of the Total Environment 741 (2020), p. 140489. doi: 10.1016/j.scitotenv.2020.140489.

[15] Ho Fai Chan et al. “How confidence in health care systems affects mobility and compliance during the COVID-19 pandemic”. In: PloS one 15.10 (2020), e0240644. doi: 10.1371/journal.pone.0240644.

[16] Tianqi Chen and Carlos Guestrin. “Xgboost: A scalable tree boosting system”. In: Proceedings of the 22nd acm sigkdd international conference on knowledge discovery and data mining. 2016, pp. 785–794. doi: 10.1145/2939672.2939785.

[17] Jin Young Chung, Choong-Ki Lee, and Yae-Na Park. “Trust in social non-pharmaceutical interventions and travel intention during a pandemic”. In: Journal of Vacation Marketing 27.4 (2021), pp. 437–448. doi: 10.1177/13567667211009584.

[18] Marco Ciotti et al. “The COVID-19 pandemic”. In: Critical reviews in clinical laboratory sciences 57.6 (2020), pp. 365–388. doi: 10.1080/10408363.2020.1783198.

[19] C De Boor. “A practical guide to splines”. In: Springer-Verlag google schola 2 (1978), pp. 4135–4195. doi: 10.2307/2006241.

[20] Facebook Data for Good. COVID-19 symptom survey – request for data access. 2020. url: https://dataforgood.fb.com/docs/covid-19-symptom-survey-request-for-data-access/ (visited on 05/27/2024).

[21] J Fan, et al. “The University of Maryland Social Data Science Center Global COVID-19 Trends and Impact Survey”. In: Partnership with Facebook (2020).

[22] Santi Garćıa-Cremades, et al. “Improving prediction of COVID-19 evolution by fusing epidemiological and mobility data”. In: Scientific Reports 11.1 (2021), p. 15173. doi: 10.1038/s41598-021-94696-2.

[23] Grace Guan et al. “Early detection of COVID-19 outbreaks using human mobility data”. In: PloS one 16.7 (2021), e0253865. doi: 10.1371/journal.pone.0253865.

[24] Emanuele Guidotti. “A worldwide epidemiological database for COVID-19 at fine-grained spatial resolution”. In: Scientific Data 9.1 (2022), p. 112. doi: 10.1038/s41597-022-01245-1.

[25] Emanuele Guidotti and David Ardia. “COVID-19 Data Hub”. In: Journal of Open Source Software 5.51 (2020), p. 2376. doi: 10.21105/joss.02376.

[26] Thomas Hale et al. “A global panel database of pandemic policies (Oxford COVID-19 Government Response Tracker)”. In: Nature human behaviour 5.4 (2021), pp. 529–538. doi: 10.1038/s41562-021-01079-8.

[27] John A Hartigan, Manchek A Wong, et al. “A k-means clustering algorithm”. In: Applied statistics 28.1 (1979), pp. 100–108. doi: 10.2307/2346830.

[28] Tao Hu et al. “Human mobility data in the COVID-19 pandemic: characteristics, applications, and challenges”. In: International Journal of Digital Earth 14.9 (2021), pp. 1126–1147. doi: 10.1080/17538947.2021.1952324.

[29] Cornelia Ilin et al. “Public mobility data enables COVID-19 forecasting and management at local and global scales”. In: Scientific reports 11.1 (2021), p. 13531. doi: 10.1038/s41598-021-92892-8.

[30] Frauke Kreuter et al. “Partnering with a global platform to inform research and public policy making: What needs to be in place to make a global COVID-19 survey work?” In: Survey Research Methods. Vol. 14. 2. 2020, pp. 159–163. doi: 10.18148/srm/2020.v14i2.7761.

[31] Yu Liu, Richard B Saltman, and Ming-Jui Yeh. “From bureaucratic administration to effective intervention: Comparing early governmental responses to the COVID-19 virus across East Asian and western health systems”. In: Health Services Management Research 36.3 (2023), pp. 193–204. doi: 10.1177/09514848221139680.

[32] Edouard Mathieu, et al. “Coronavirus Pandemic (COVID-19)”. In: Our World in Data (2020). https://ourworldindata.org/coronavirus.

[33] Zahra Mohammadi, Monica Gabriela Cojocaru, and Edward Wolfgang Thommes. “Human behaviour, NPI and mobility reduction effects on COVID-19 transmission in different countries of the world”. In: BMC Public Health 22.1 (2022), p. 1594. doi: 10.1186/s12889-022-13921-3.

[34] Marcus Painter and Tian Qiu. “Political beliefs affect compliance with covid-19 social distancing orders”. In: Covid Economics 4.April (2020), pp. 103–23. doi: 10.1016/j.jebo.2021.03.019.

[35] Simone Pernice et al. “CONNECTOR, fitting and clustering of longitudinal data to reveal a new risk stratification system”. In: Bioinformatics 39.5 (2023), btad201. doi: 10.1093/bioinformatics/btad201.

[36] Nicola Perra. “Non-pharmaceutical interventions during the COVID-19 pandemic: A review”. In: Physics Reports 913 (2021), pp. 1–52. doi: 10.1016/j.physrep.2021.02.001.

[37] Wenjing Pian, Jianxing Chi, and Feicheng Ma. “The causes, impacts and countermeasures of COVID-19 “Infodemic”: A systematic review using narrative synthesis”. In: Information processing & management 58.6 (2021), p. 102713. doi: 10.1016/j.ipm.2021.102713.

[38] Jesús Rufino, et al. “Using survey data to estimate the impact of the omicron variant on vaccine efficacy against COVID-19 infection”. In: Scientific Reports 13.1 (2023), p. 900. doi: 10.1038/s41598-023-27951-3.

[39] Joshua A Salomon et al. “The US COVID-19 Trends and Impact Survey: Continuous real-time measurement of COVID-19 symptoms, risks, protective behaviors, testing, and vaccination”. In: Proceedings of the National Academy of Sciences 118.51 (2021), e2111454118. doi: 10.1073/pnas.2111454118.

[40] Ketan Rajshekhar Shahapure and Charles Nicholas. “Cluster quality analysis using silhouette score”. In: 2020 IEEE 7th international conference on data science and advanced analytics (DSAA). IEEE. 2020, pp. 747–748. doi: 10.1109/DSAA49011.2020.000 96.

[41] Wenjing Shao, Jingui Xie, and Yongjian Zhu. “Mediation by human mobility of the association between temperature and COVID-19 transmission rate”. In: Environmental research 194 (2021), p. 110608. doi: 10.1016/j.envres.2020.110608.

[42] Berber T Snoeijer et al. “Measuring the effect of Non-Pharmaceutical Interventions (NPIs) on mobility during the COVID-19 pandemic using global mobility data”. In: NPJ digital medicine 4.1 (2021), p. 81. doi: 10.1038/s41746-021-00451-2.

[43] Charles Spearman. “The proof and measurement of association between two things.” In: (1961). doi: https://psycnet.apa.org/doi/10.2307/1412159.

[44] Sean J Taylor and Benjamin Letham. “Forecasting at scale”. In: The American Statistician 72.1 (2018), pp. 37–45. doi: https://peerj.com/preprints/3190v2/.

[45] The University of Maryland Social Data Science Center. COVID19 symptom survey intl V11 noneu. Accessed 2 July 2024. 2021. url: https://covidmap.umd.edu/document/COVID19_symptom_survey_intl_V11_0723.pdf.

[46] Ahmad Ilderim Tokey. “Spatial association of mobility and COVID-19 infection rate in the USA: A county-level study using mobile phone location data”. In: Journal of Transport & Health 22 (2021), p. 101135. doi: 10.1016/j.jth.2021.101135.

[47] Benjamin Van Rooij et al. “Compliance with COVID-19 mitigation measures in the United States”. In: Amsterdam law school research paper 2020-21 (2020). doi: 10.2139/ssrn.3582626.

[48] Jack West, Serenydd Everden, and Nikitas Nikitas. “A case of COVID-19 reinfection in the UK”. In: Clinical medicine 21.1 (2021), e52. doi: 10.7861/clinmed.2020-0912.

[49] WHO COVID-19 dashboard. 2024. url: https://data.who.int/dashboards/covid19/deaths?n=o (visited on 11/24/2024).

[50] Mengxi Zhang et al. “Human mobility and COVID-19 transmission: a systematic review and future directions”. In: Annals of GIS 28.4 (2022), pp. 501–514. doi: 10.1080/19475683.2022.2041725.

