## Supplementary Material for "Social Compliance with NPIs, Mobility Patterns, and Reproduction Number: Lessons from COVID-19 in Europe"

#### Contents

**1 Introduction** **1**

**2 Extended data analysis** **1**

**3 Extended clustering** **10**

**4 Extended results on regression models** **16**

**5 How to reproduce the results** **30**

### 1 Introduction

In this document, we provide additional details on the preliminary data analysis conducted for each country (Section 2). We also present clustering results using CONNECTOR [13] (Section 3), which is applied directly to time series data, in contrast to K-means [7], which relies on average values and, eventually, standard deviations. Next, we show more details on the XGBoost [2] regression models (Section 4). Finally, we provide instructions on how to reproduce the results (Section 5).

### 2 Extended data analysis

In this section, we show detailed plots for each of the 13 countries considered. Specifically, Figures SM1 and SM2 show the time evolution of each of the six considered Facebook mobility variables—*grocery*, *residential*, *stations*, *recreation*, *workplaces*, and *masks*—, the *stringency* index, and the *infection rates* from April 2020 to May 2021. Figures SM5 and SM6 show the same information considering Google mobility variables instead of Facebook ones—except for the Facebook *masks* variable—from April 2020 to March 2022. Figures SM3 and SM4 show the average Facebook mobility—computed on *grocery*, *residential*, *stations*, *recreation*, and *workplaces*—, along with the complement of the *stringency* index and *infection rates*. Figures SM7 and SM8 show the same information considering Google mobility variables instead of Facebook ones from April 2020 to March 2022.

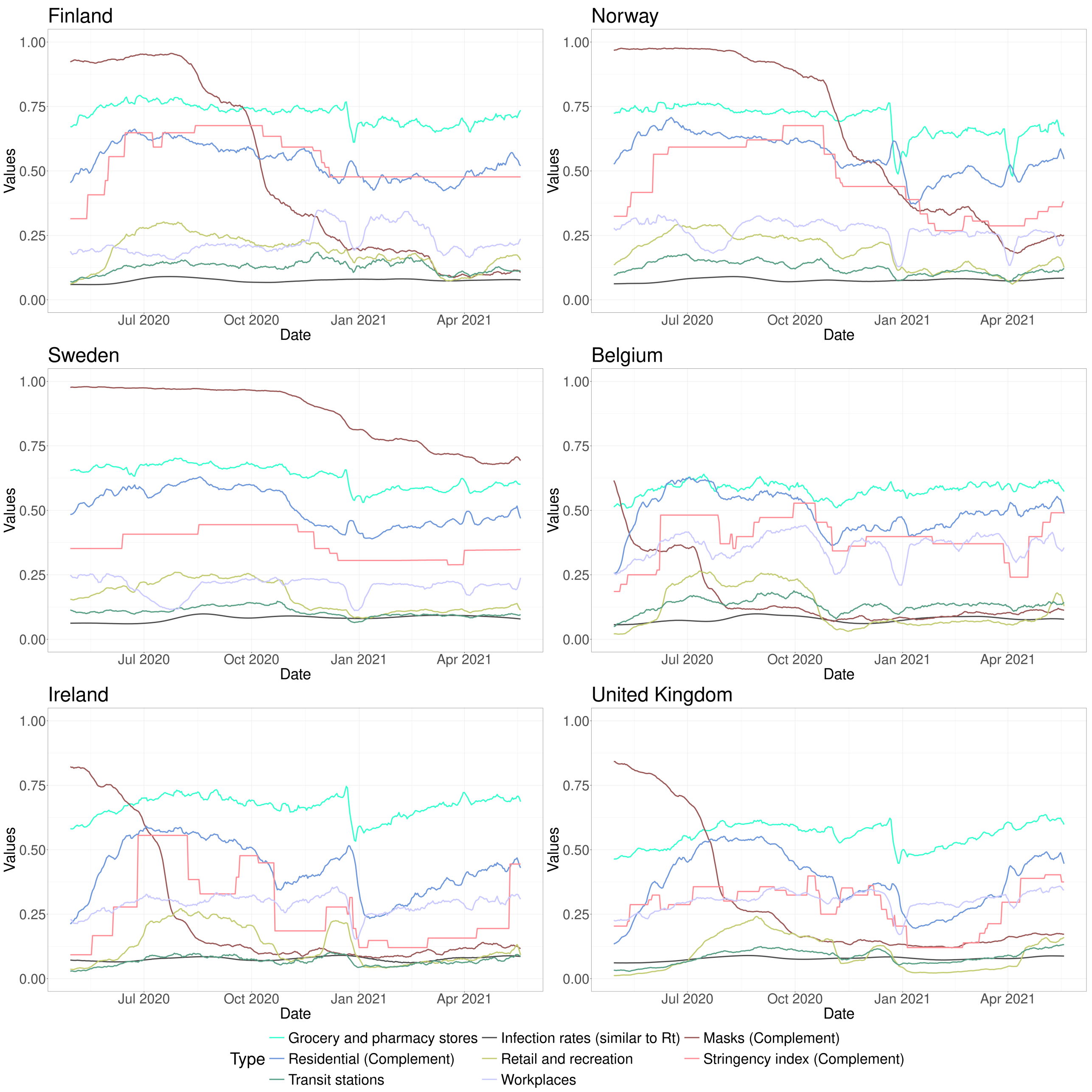

Figure SM1: Evolution of the considered Facebook mobility variables—*grocery*, *residential*, *stations*, *recreation*, *workplaces*, and *masks*—, alongside the complement of the *stringency* index and *infection rates* for Finland, Norway, Sweden, Belgium, Ireland, and the United Kingdom from April 2020 to May 2021.

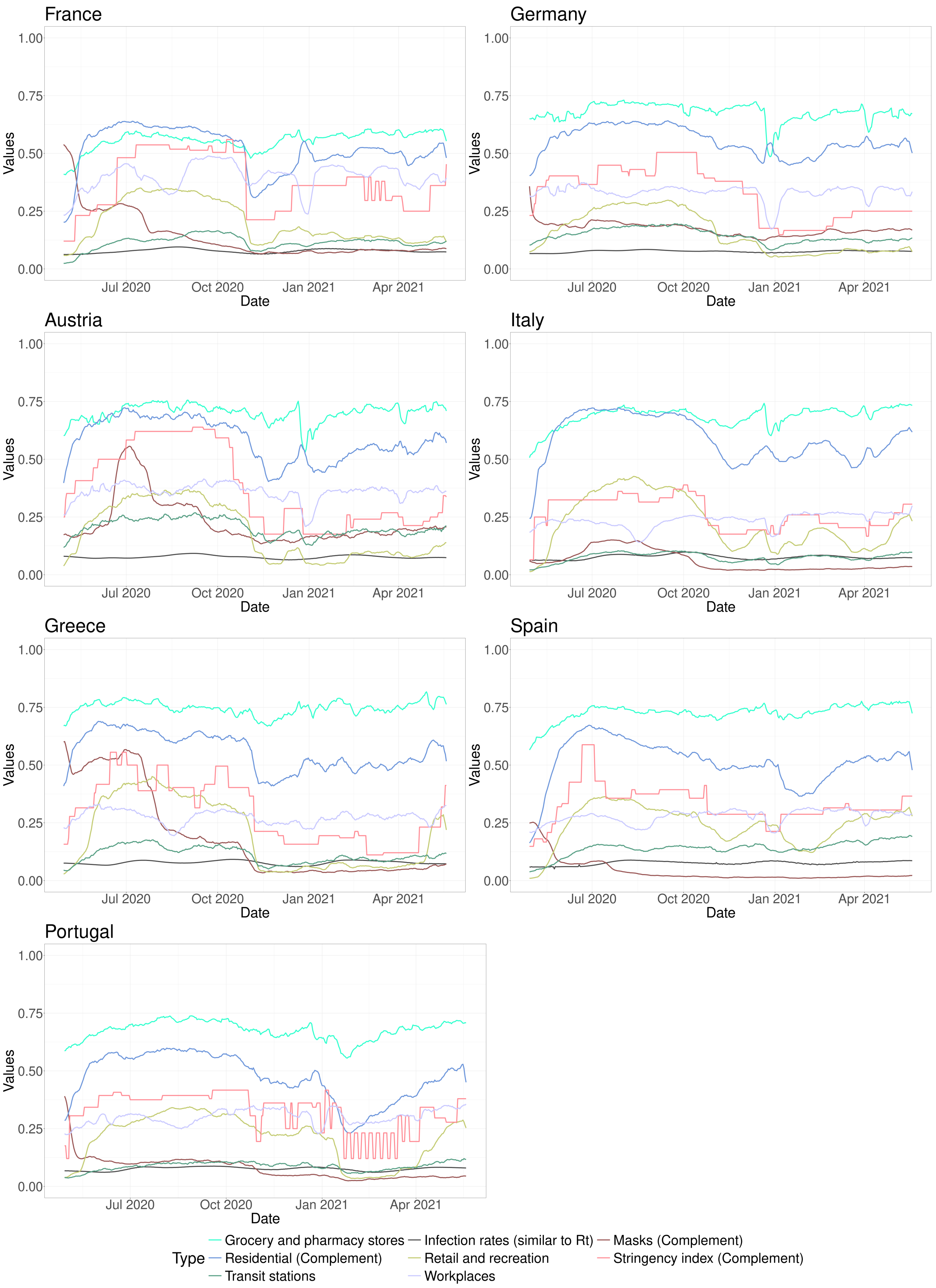

Figure SM2: Evolution of the considered Facebook mobility variables—*grocery*, *residential*, *stations*, *recreation*, *workplaces*, and *masks*—, alongside the complement of the *stringency* index and *infection rates* for France, Germany, Austria, Italy, Greece, Spain, and Portugal from April 2020 to May 2021.

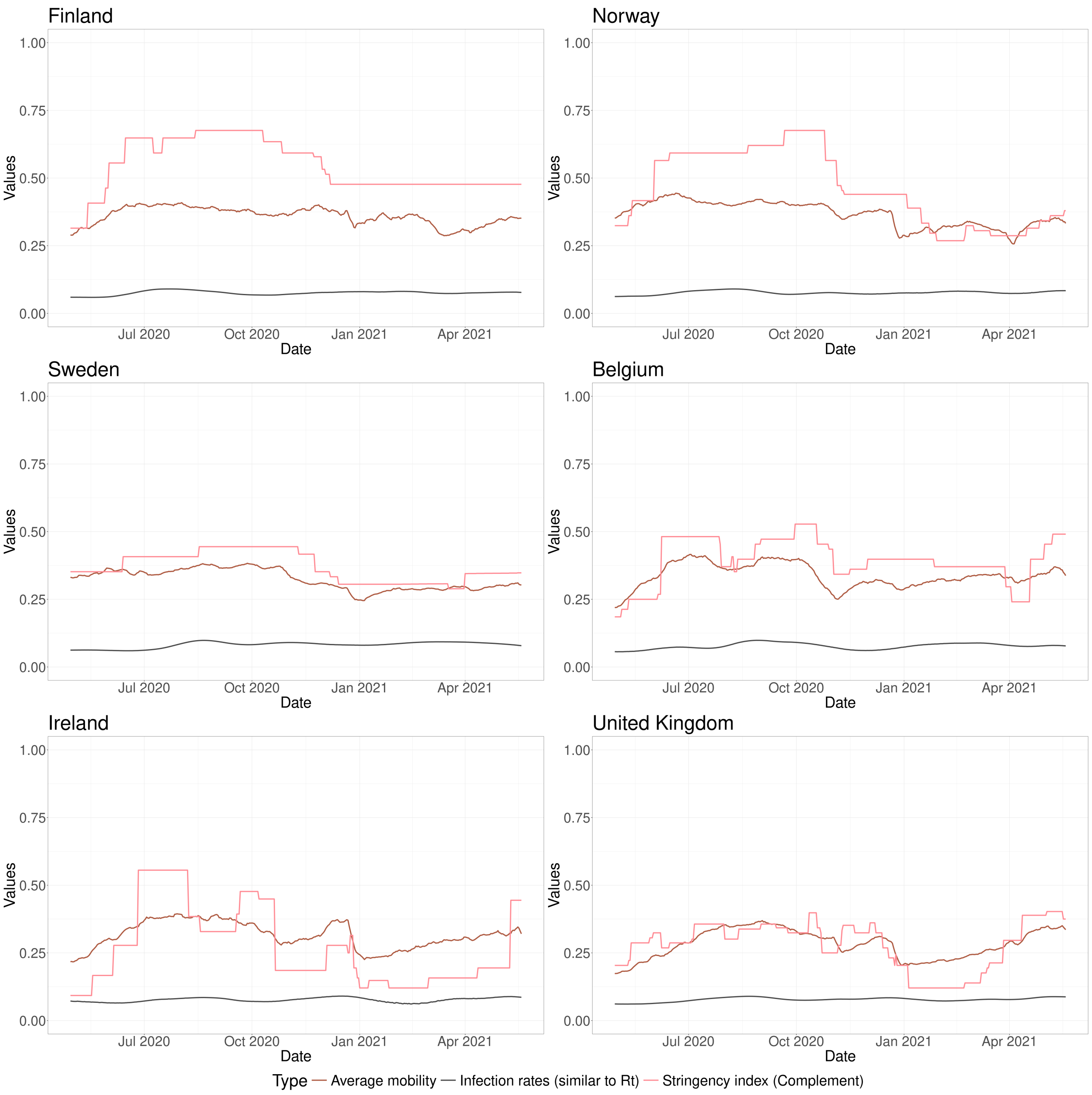

Figure SM3: Evolution of the average Facebook mobility—computed on *grocery*, *residential*, *stations*, *recreation*, and *workplaces*—, alongside the complement of the *stringency* index and *infection rates* for Finland, Norway, Sweden, Belgium, Ireland, and the United Kingdom from April 2020 to May 2021.

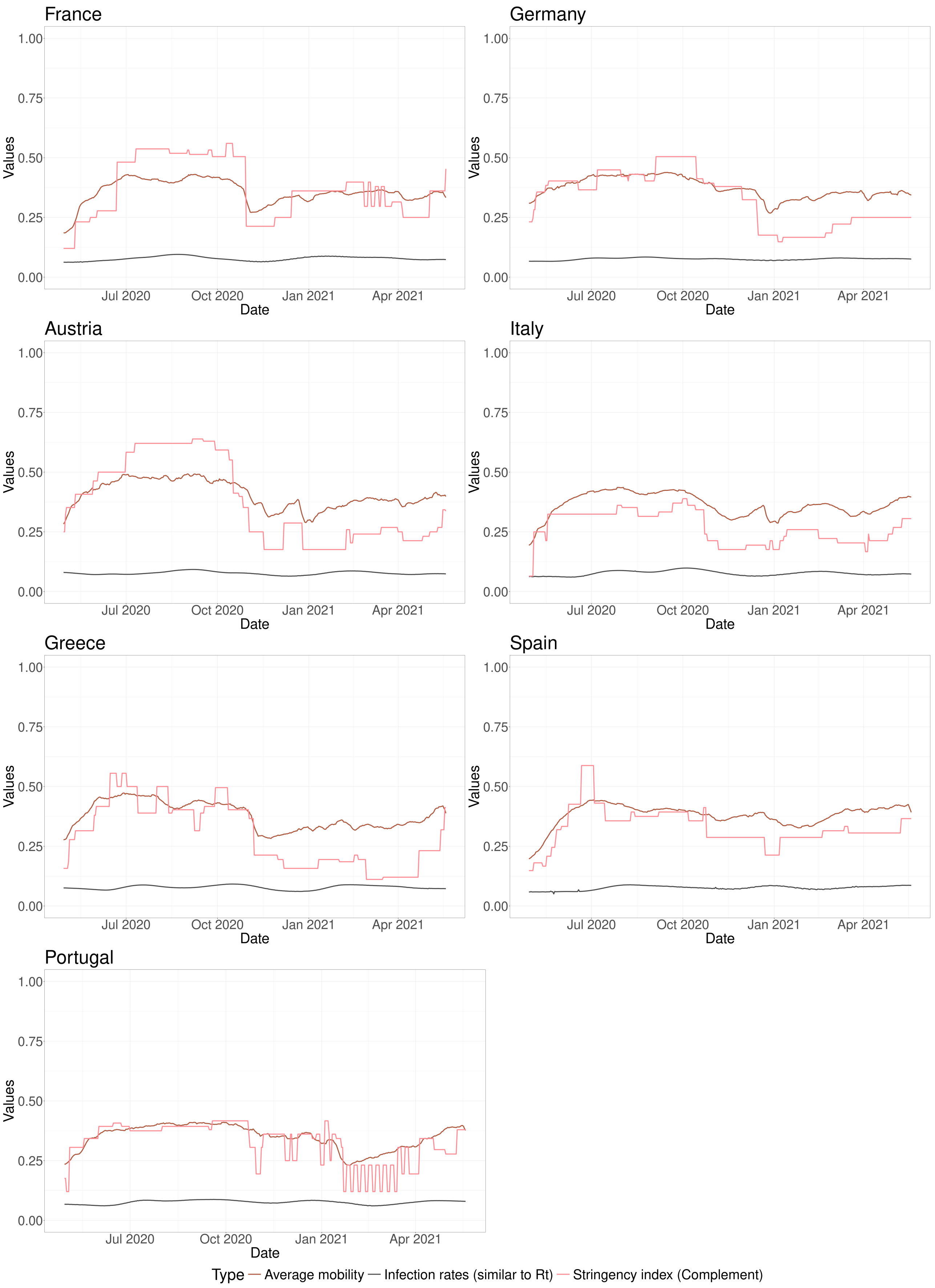

Figure SM4: Evolution of the average Facebook mobility—computed on *grocery*, *residential*, *stations*, *recreation*, and *workplaces*—, alongside the complement of the *stringency* index and *infection rates* for France, Germany, Austria, Italy, Greece, Spain, and Portugal from April 2020 to May 2021.

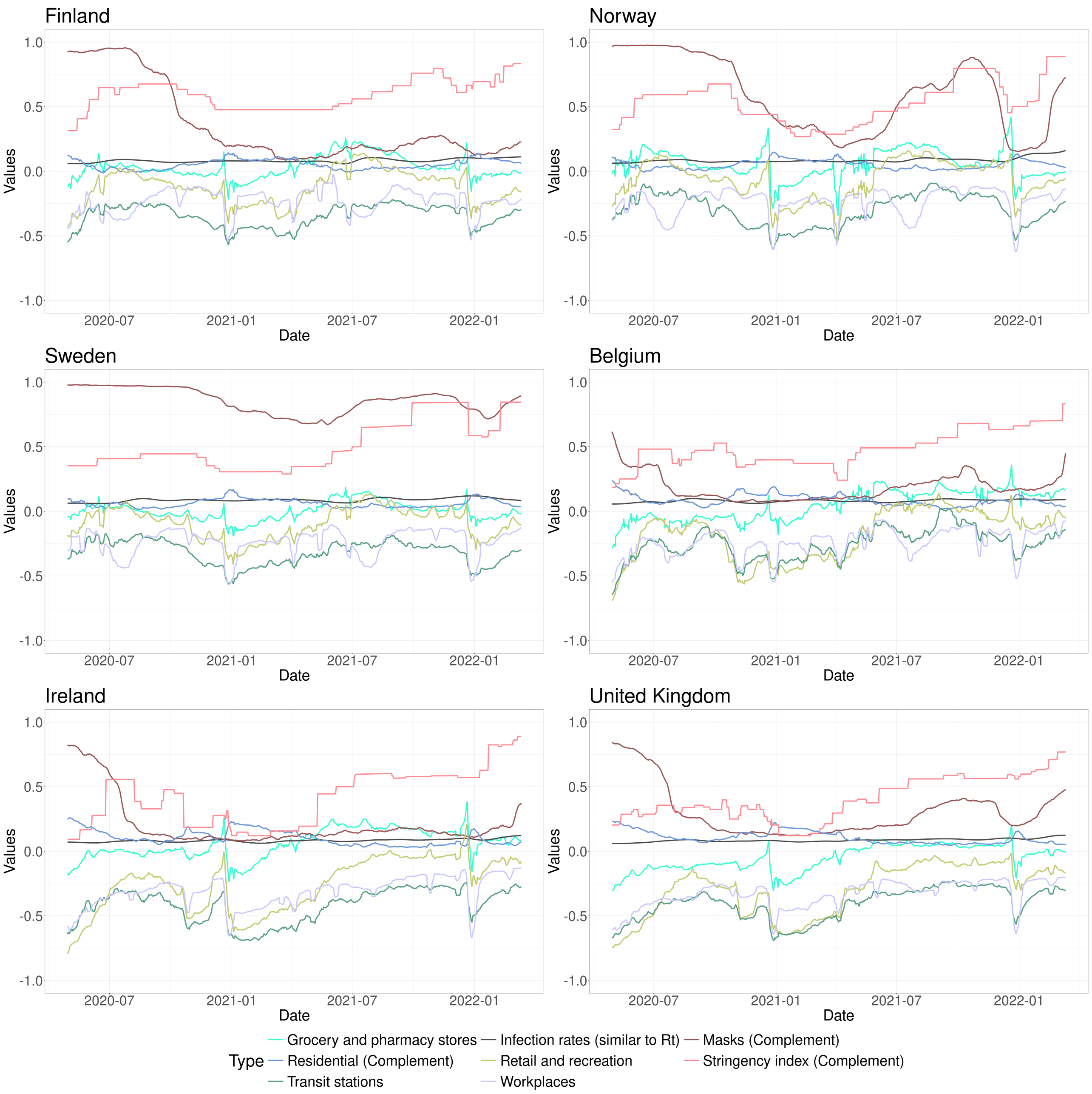

Figure SM5: Evolution of the considered Google mobility variables—*grocery*, *residential*, *stations*, *recreation*, *workplaces*, and the Facebook *masks*—, alongside the complement of the *stringency* index and *infection rates* for Finland, Norway, Sweden, Belgium, Ireland, and the United Kingdom from April 2020 to March 2022.

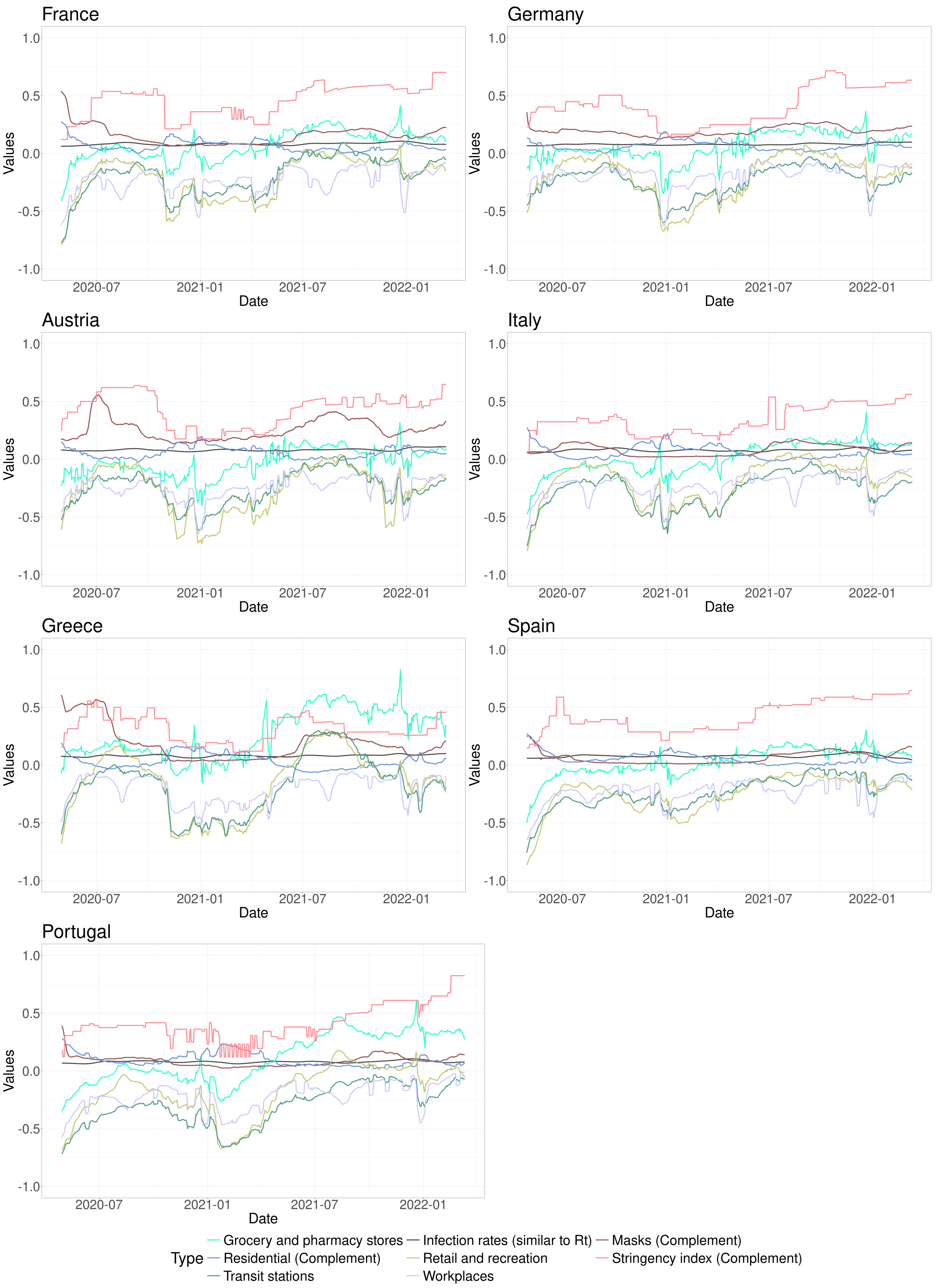

Figure SM6: Evolution of the considered Google mobility variables—*grocery*, *residential*, *stations*, *recreation*, *workplaces*, and the Facebook *masks*—, alongside the complement of the *stringency* index and *infection rates* for France, Germany, Austria, Italy, Greece, Spain, and Portugal from April 2020 to March 2022.

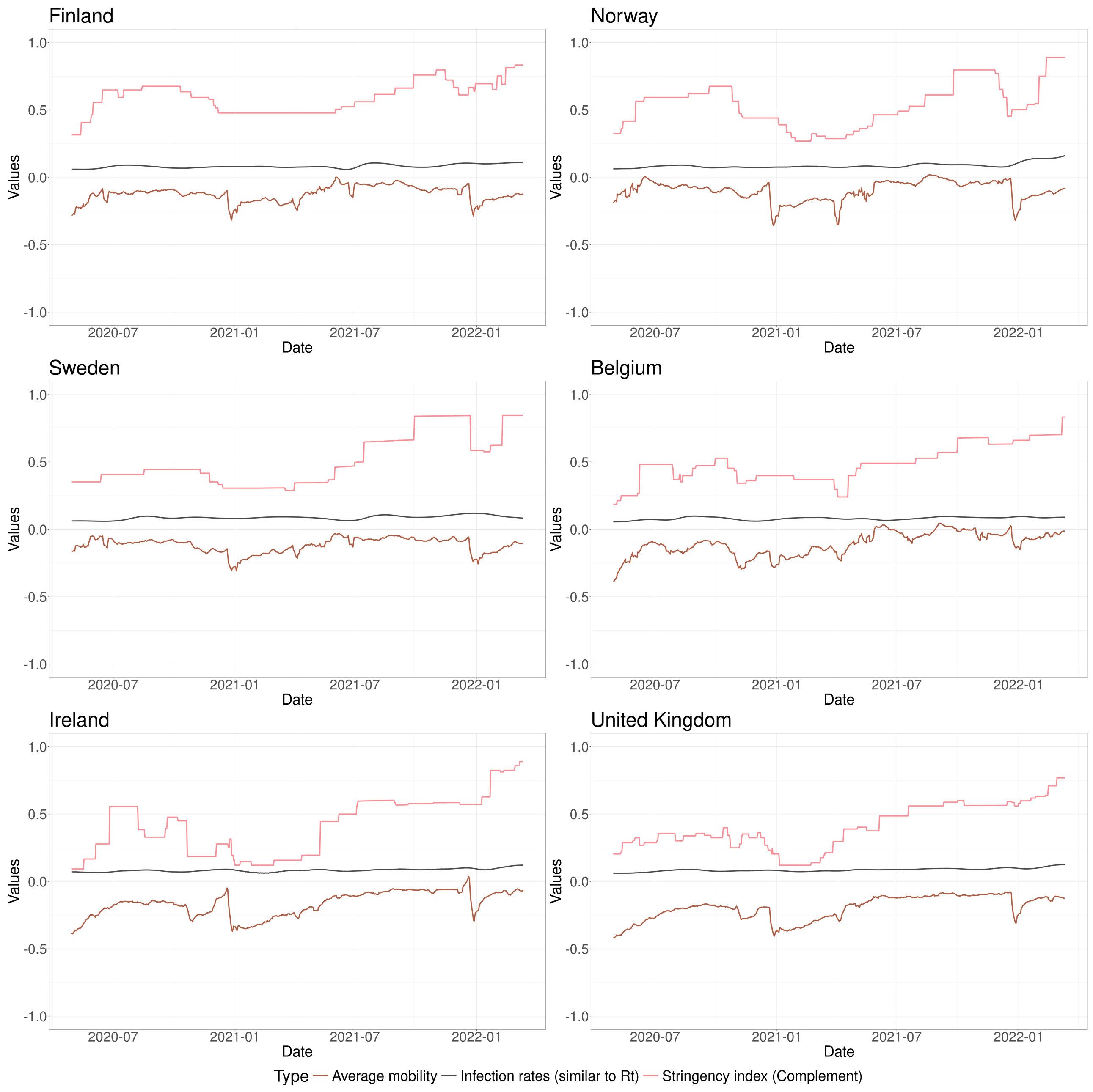

Figure SM7: Evolution of the average Google mobility—computed on *grocery*, *residential*, *stations*, *recreation*, and *workplaces*—, alongside the complement of the *stringency* index and *infection rates* for Finland, Norway, Sweden, Belgium, Ireland, and the United Kingdom from April 2020 to March 2022.

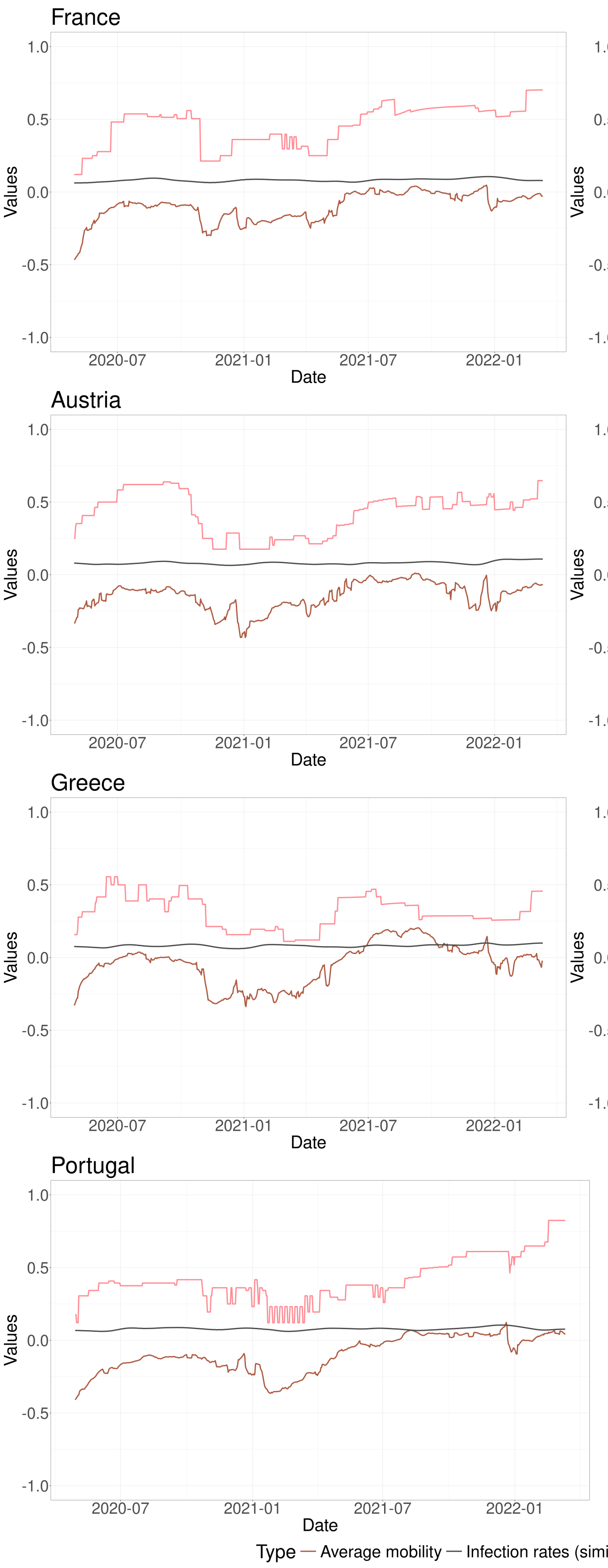

Figure SM8: Evolution of the average Google mobility—computed on *grocery*, *residential*, *stations*, *recreation*, and *workplaces*—, alongside the complement of the *stringency* index and *infection rates* for France, Germany, Austria, Italy, Greece, Spain, and Portugal from April 2020 to March 2022.

### 3 Extended clustering

In this section, we cluster countries based on the ratios defined in the main document using an alternative clustering technique, CONNECTOR [13]. As detailed in the *Methods* section of the main document, CONNECTOR is an R package designed for the unsupervised analysis of longitudinal data. It employs a model-based approach to cluster functional data, making it particularly effective for handling sparse and irregular curves.

Figure SM9 shows the dimension of the spline basis vector, determined by the *cross-log-likelihood*—see [13] for more details. This dimension can be chosen as the one corresponding to the largest cross-validated likelihood, as proposed in [9]. In particular, we chose 8 for the $ComplStrIdxOnAvgMob(t)$ ratio and 11 for the $AvgMobOnInfRates(t)$ and $ComplStrIdxOnInfRates(t)$ ratios.

Figure SM10 illustrates the cluster separation measure, known as fDB, as defined in [13]. This measure is an adaptation of the well-known Davies–Bouldin (DB) index [3] for functional data. Additionally, the figure presents the total tightness, a metric used to quantify dispersion within clusters. As the number of clusters increases, total tightness decreases, approaching zero when the number of fitted clusters matches the number of sampled curves. For this analysis, we select two, five, and six clusters for each ratio.

Figure SM11 illustrates the clustering results for the $ComplStrIdxOnAvgMob(t)$ ratio, with configurations of two, five, and six clusters. For the two-cluster configuration, the results replicate the separation between Northern and Southern countries achieved using K-means in the main document. Specifically, Finland, Norway, Sweden, and Belgium form one cluster, while the remaining countries are grouped into the other cluster. In the five-cluster configuration, Finland forms its own cluster, while the United Kingdom constitutes another separate cluster. Italy, Spain, and Portugal are grouped into a third cluster. Norway, Sweden, Belgium, and France are assigned to a fourth cluster, while Ireland, Germany, Austria, and Greece comprise the fifth cluster. In this case, while the clusters differ slightly from those obtained using K-means, the separation between Northern and Southern countries remains evident. Finally, in the six-cluster configuration, the fourth cluster is divided into two subgroups: Germany and Austria form one cluster, while Ireland and Greece constitute the other.

Figure SM12 illustrates the clustering results for the $AvgMobOnInfRates(t)$ ratio with two, five, and six cluster configurations. In the two-cluster configuration, Sweden, the United Kingdom, and Ireland are grouped in the first cluster, while the remaining countries form the second cluster. This clustering yields similar results to those obtained using K-means in the main document, where Belgium was also included in the first cluster. In the five-cluster configuration, Sweden is placed in the first cluster, Finland, Norway, and Germany in the second, Ireland and the United Kingdom in the third, Austria and Spain in the fourth, and Italy, Greece, Belgium, France, and Portugal in the fifth. In this case, these clusters differ slightly from the ones obtained using K-means in the main document. Finally, for the six-cluster configuration, the fifth cluster is split into two subgroups: Italy and Greece form one cluster, while Belgium, France, and Portugal constitute the other. These six clusters are slightly different with respect to the ones obtained using K-means considering the average and the standard deviation in the previous section.

Figure SM13 illustrates the clustering results for the $ComplStrIdxOnInfRates(t)$ ratio, with configurations of two, five, and six clusters. In the two-cluster configuration, Finland and Norway form one cluster, while all other countries belong to the second cluster. As with this ratio, we observe the same separation as obtained using K-means in the main document. In the five-cluster configuration, Finland and Norway are grouped in one cluster, Ireland forms the second cluster, Austria is in the third, Sweden, Belgium, France, and Spain are in the fourth, and the United Kingdom, Germany, Italy, Greece, and Portugal are placed in the fifth cluster. Finally, for the six-cluster configuration, Finland and Norway are grouped in one cluster, Ireland forms the second cluster, Austria is in the third, Belgium and France are in the fourth, the United Kingdom, Italy, and Greece are in the fifth, and Sweden, Germany, Spain, and Portugal are in the sixth. In this case, as well, these six clusters are slightly different with respect to the ones obtained using K-means.

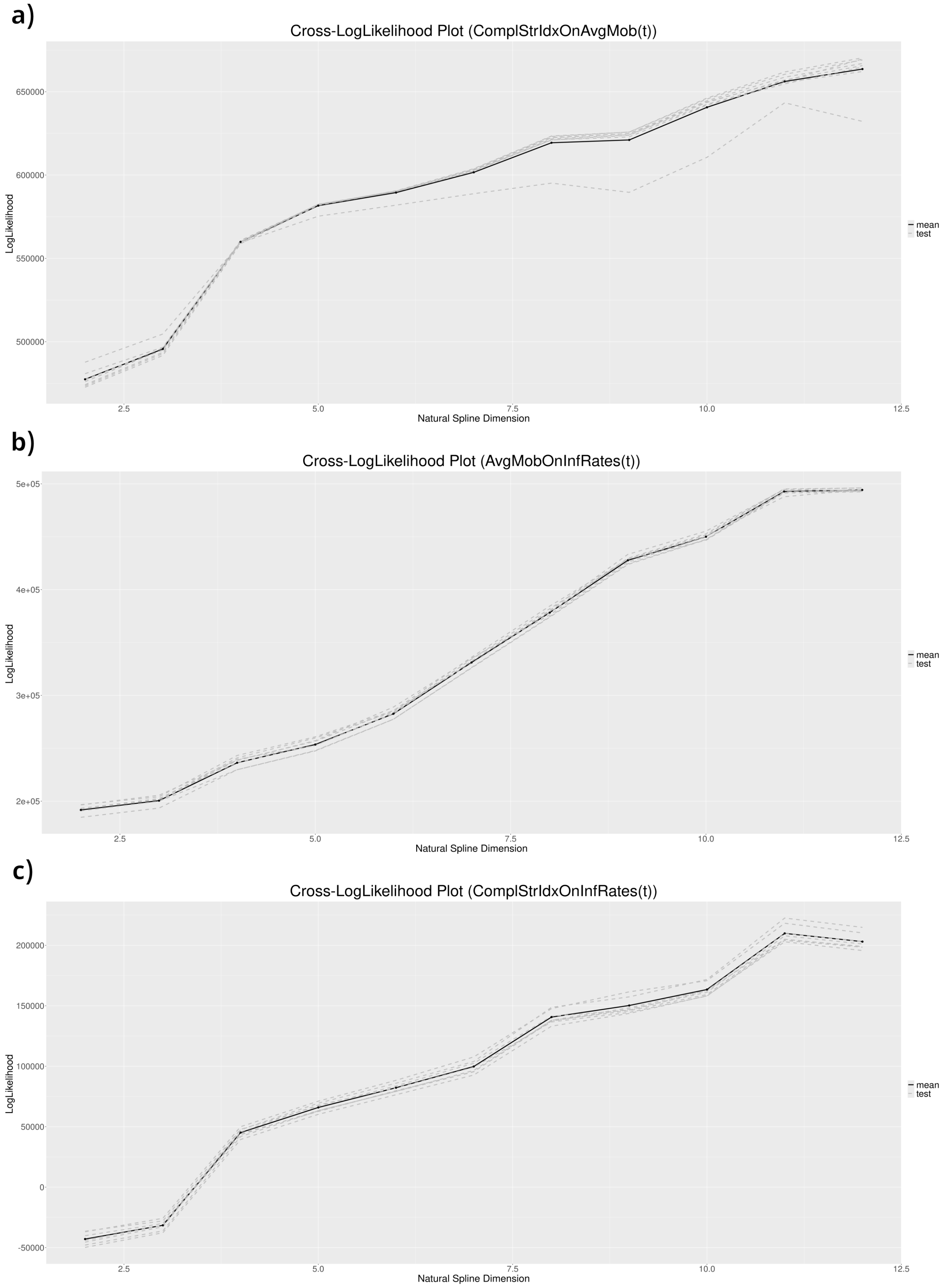

Figure SM9: The dimension of the spline basis vector, determined by the *cross-log-likelihood* [13], is shown for each ratio. The optimal dimension is selected by maximizing this value.

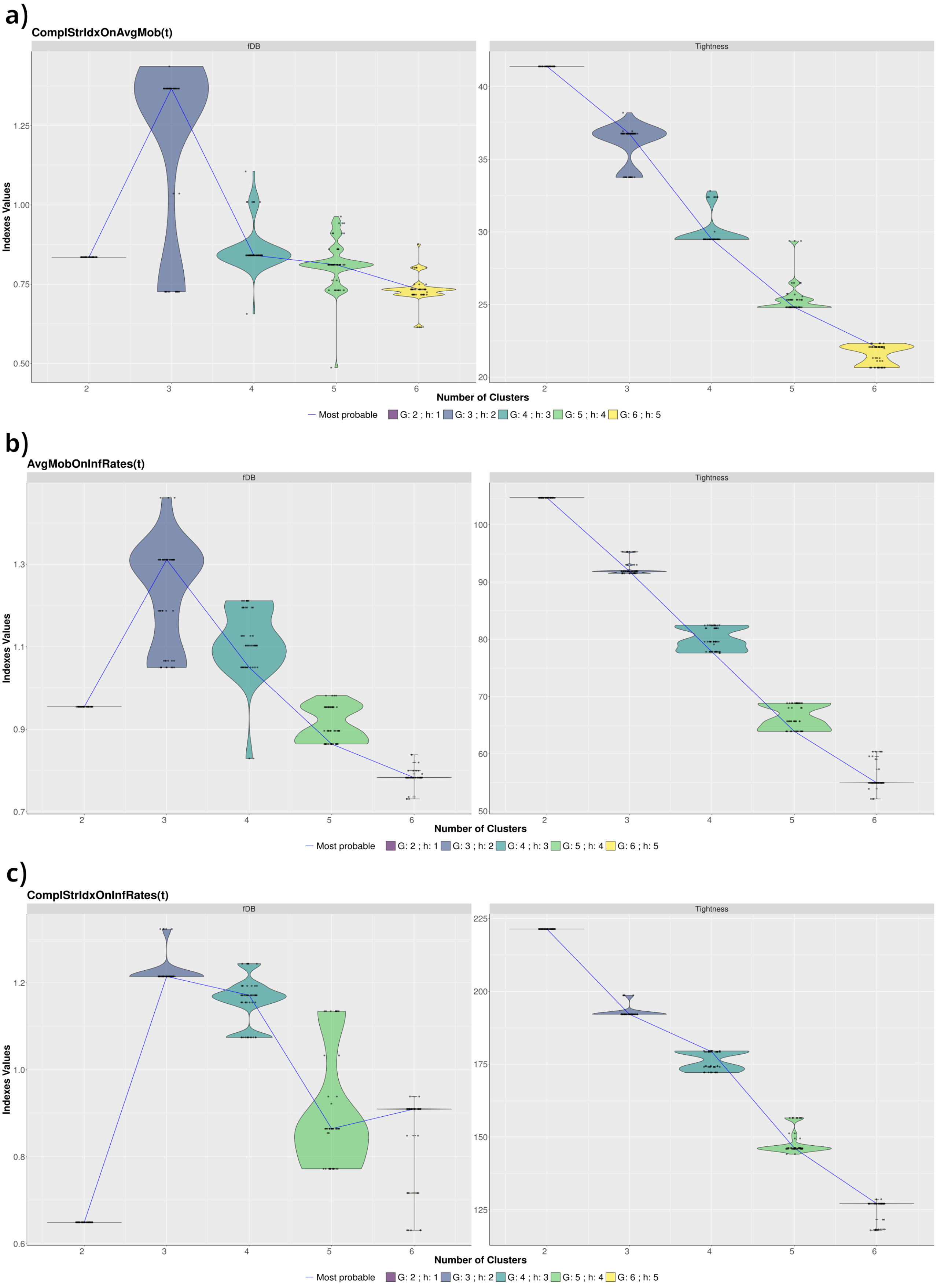

Figure SM10: Cluster separation measure (fDB) and total tightness [13] are shown for each ratio. The optimal number of clusters is chosen to minimize the fDB while balancing total tightness.

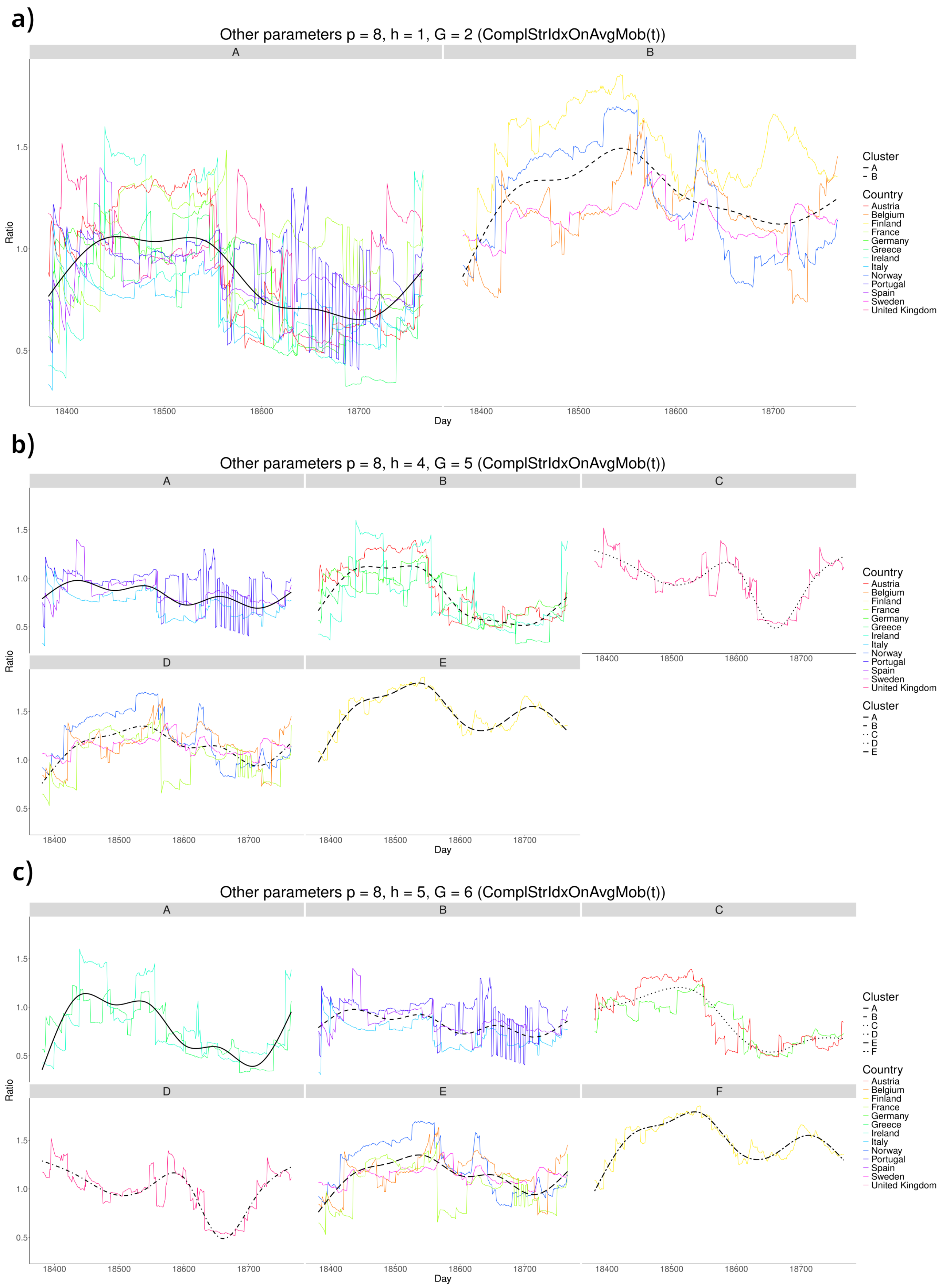

Figure SM11: Two, five, and six clusters for the $ComplStrIdxOnAvgMob(t)$ ratio.

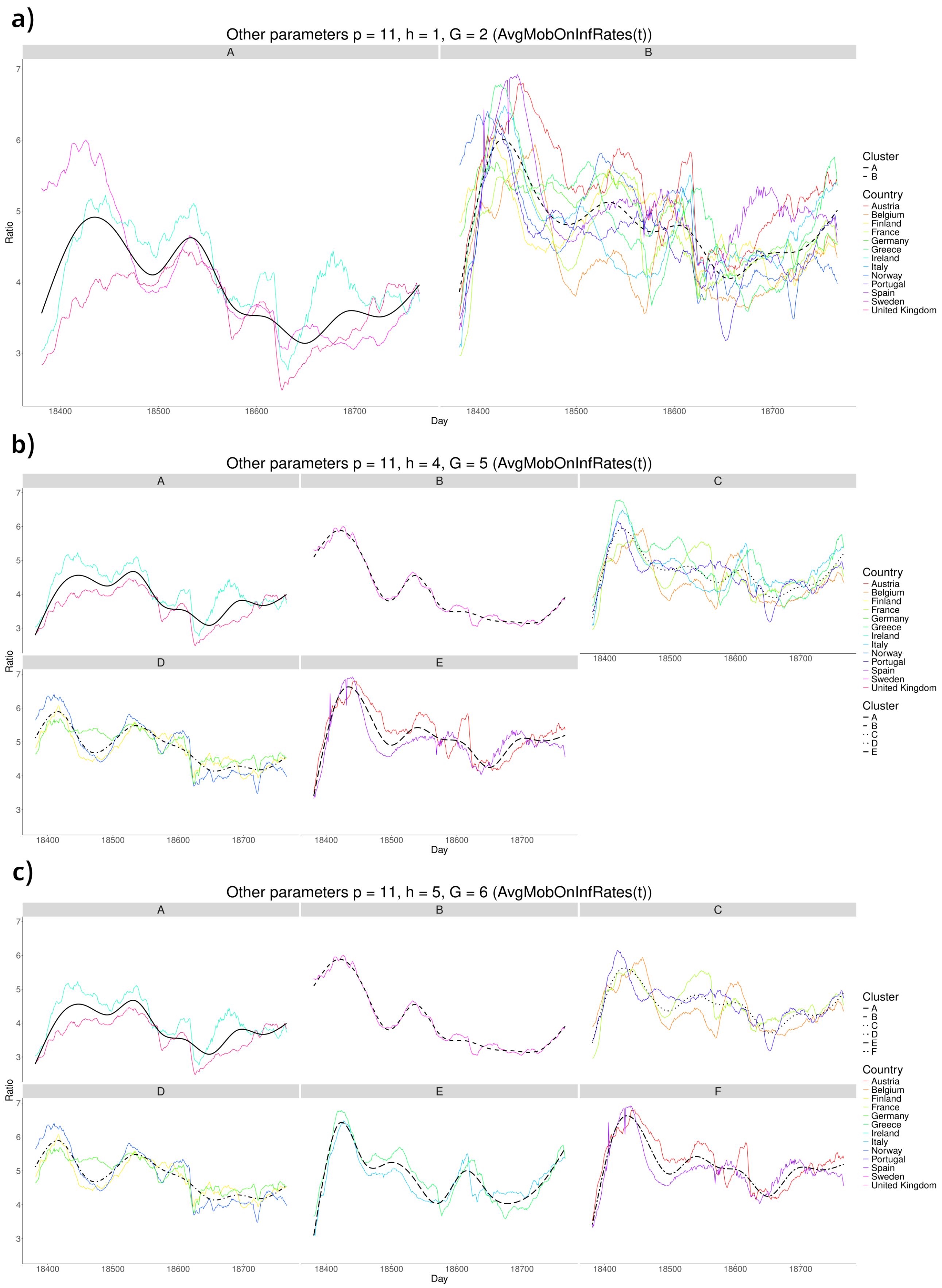

Figure SM12: Two, five, and six clusters for the $AvgMobOnInfRates(t)$ ratio.

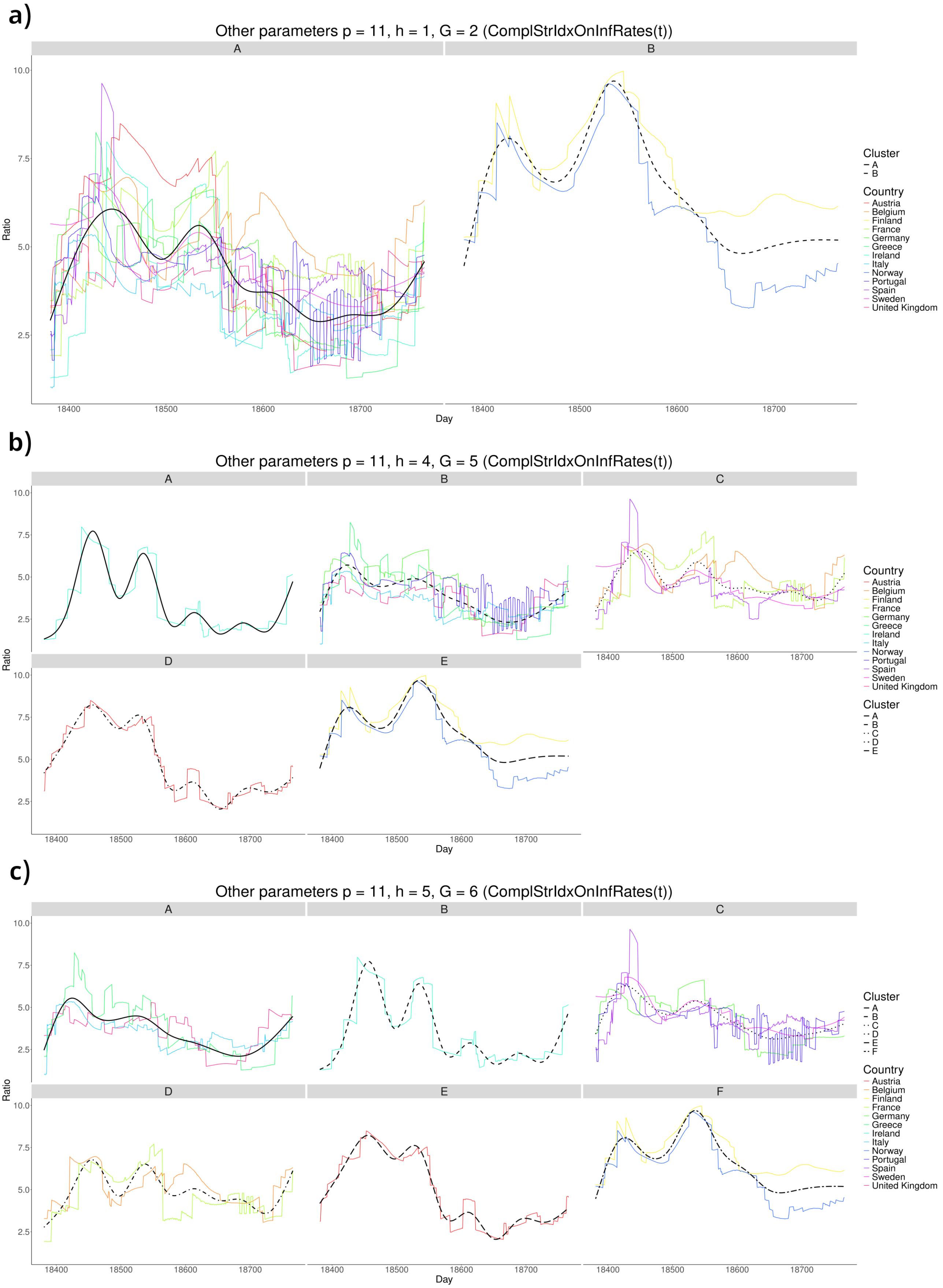

Figure SM13: Two, five, and six clusters for the $ComplStrIdxOnInfRates(t)$ ratio.

### 4 Extended results on regression models

In this section, we show more details on the XGBoost [2] regression models used in the main document to estimate missing mobility data based on NPIs and missing $R_t$ data based on mobility patterns. In particular, we show some plots that illustrate a comparison between the ground truth and the predicted values for the test set (20% of the data), for each country. For brevity, we present results for the first fold only, noting that similar observations apply consistently across the remaining four folds.

Figures SM14 to SM26 show the predictions obtained using XGBoost regression models to estimate missing mobility data based on NPIs, for each mobility variable considered. Figures SM27 and SM28 show the predictions obtained using XGBoost regression models to estimate missing $R_t$ values based on mobility patterns. These figures show that XGBoost models work very well in estimating missing data.

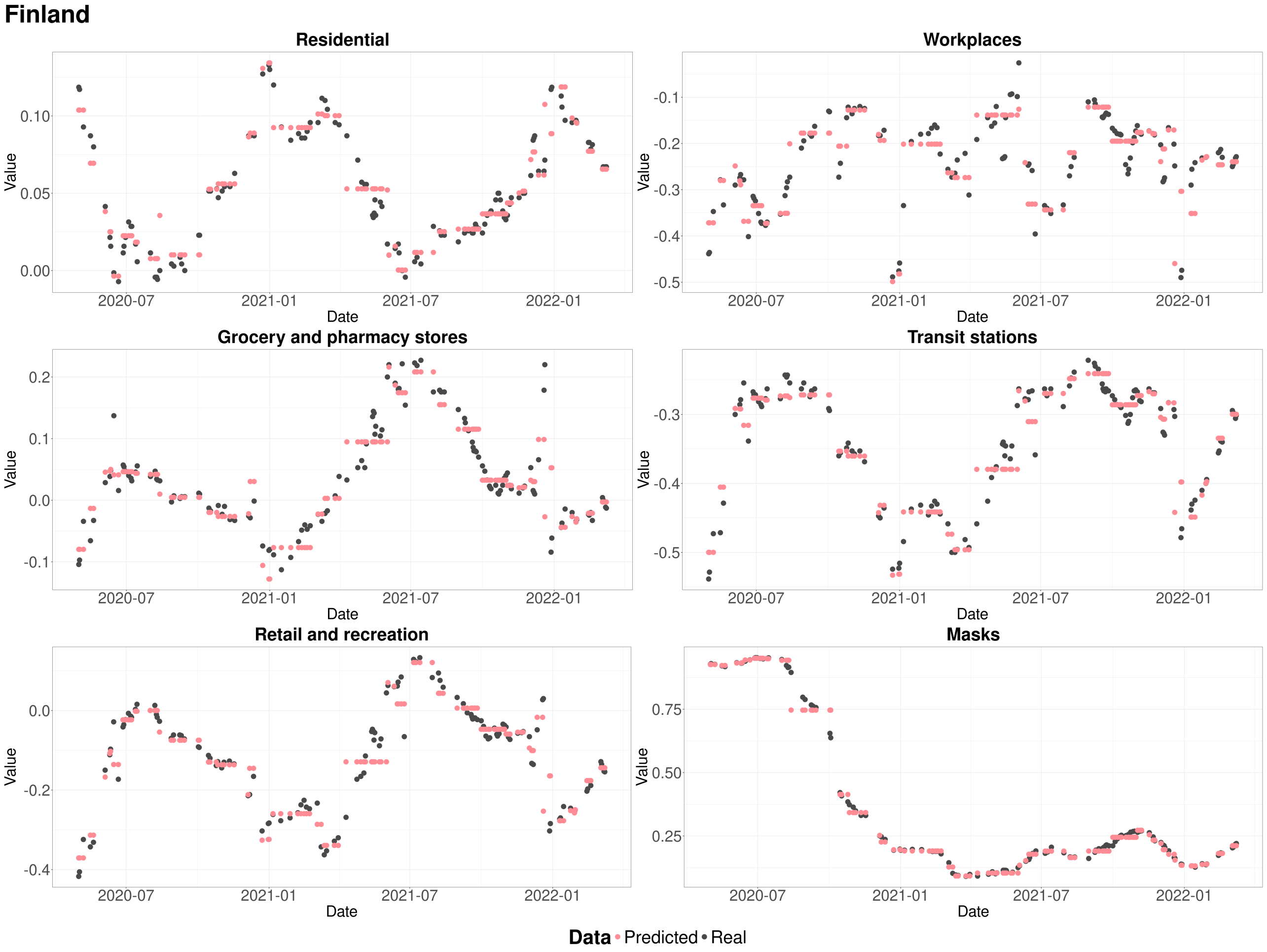

Figure SM14: Comparison between the ground truth and the predicted values for the test set (20% of the data, first fold) for the XGBoost model from NPIs to mobility in Finland.

#### Norway

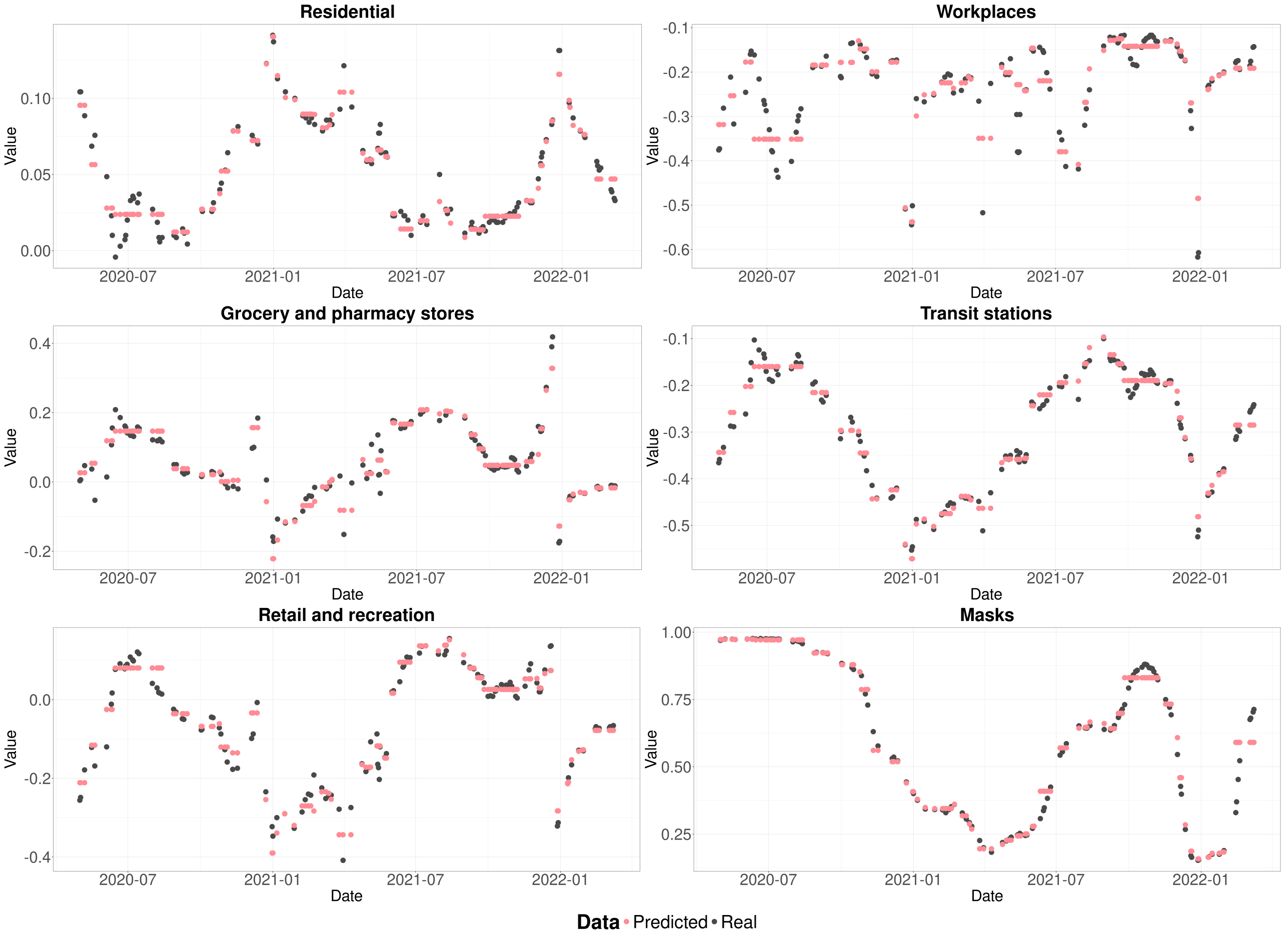

Figure SM15: Comparison between the ground truth and the predicted values for the test set (20% of the data, first fold) for the XGBoost model from NPIs to mobility in Norway.

#### Sweden

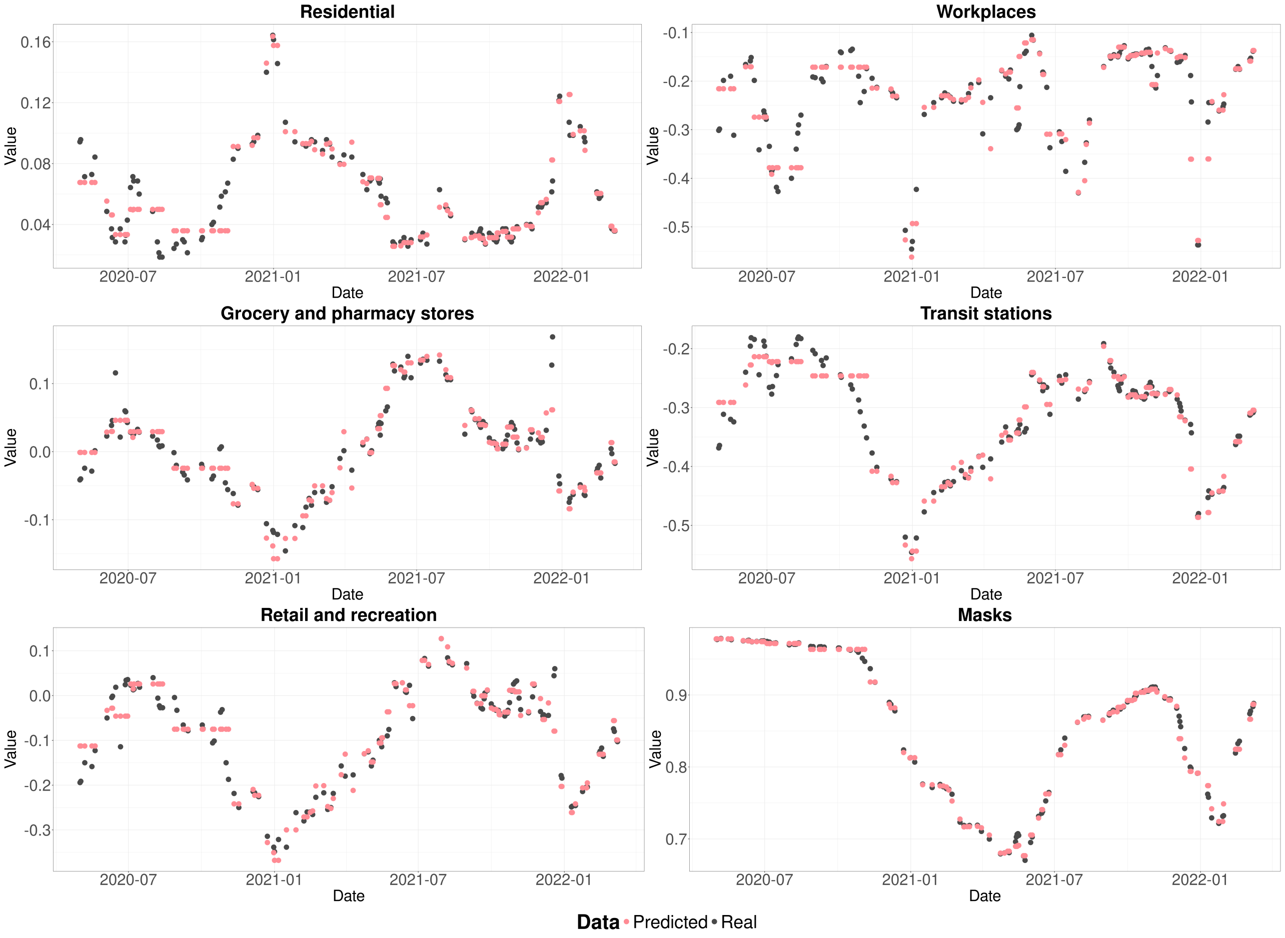

Figure SM16: Comparison between the ground truth and the predicted values for the test set (20% of the data, first fold) for the XGBoost model from NPIs to mobility in Sweden.

#### Belgium

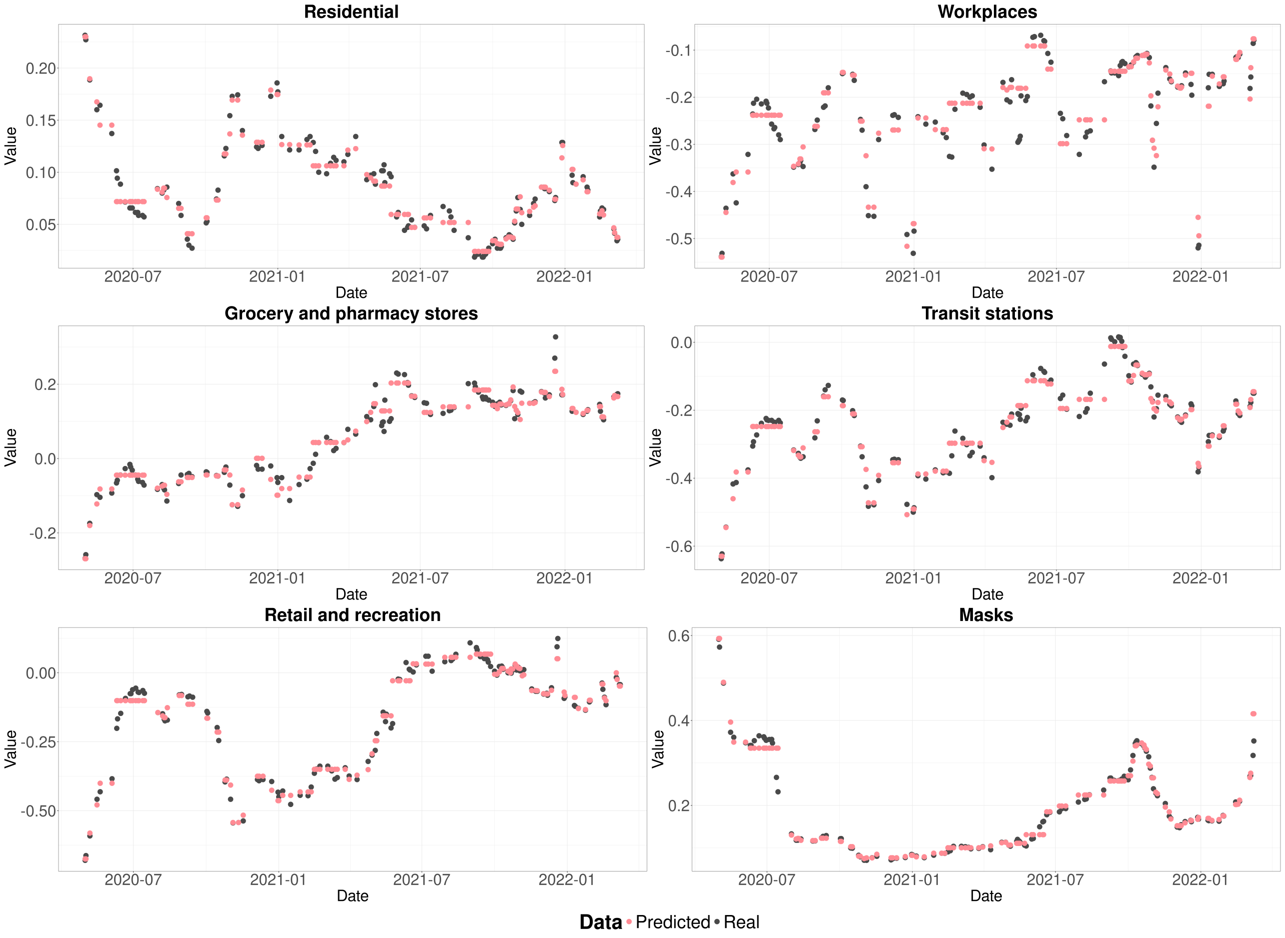

Figure SM17: Comparison between the ground truth and the predicted values for the test set (20% of the data, first fold) for the XGBoost model from NPIs to mobility in Belgium.

#### Ireland

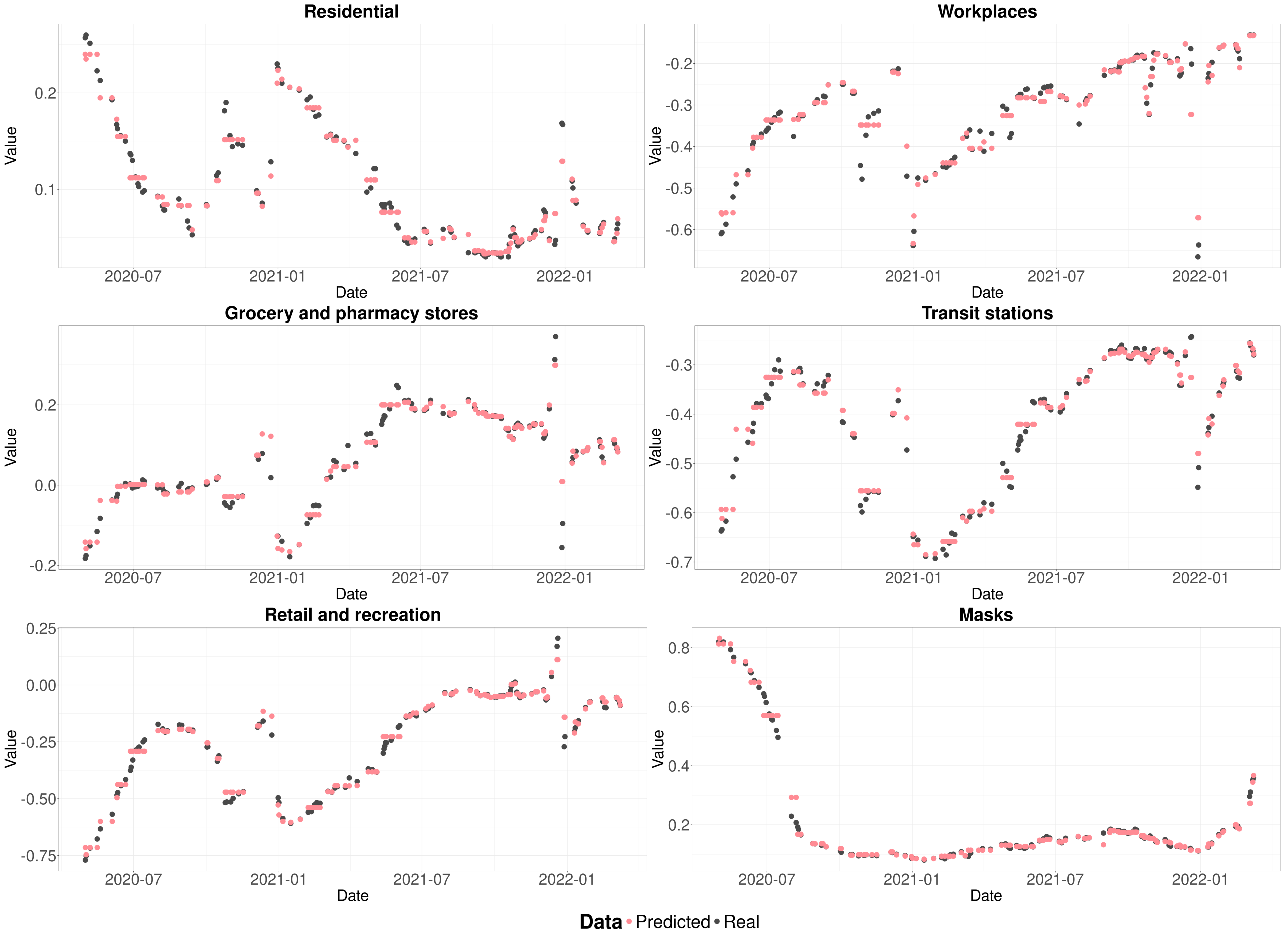

Figure SM18: Comparison between the ground truth and the predicted values for the test set (20% of the data, first fold) for the XGBoost model from NPIs to mobility in Ireland.

#### United Kingdom

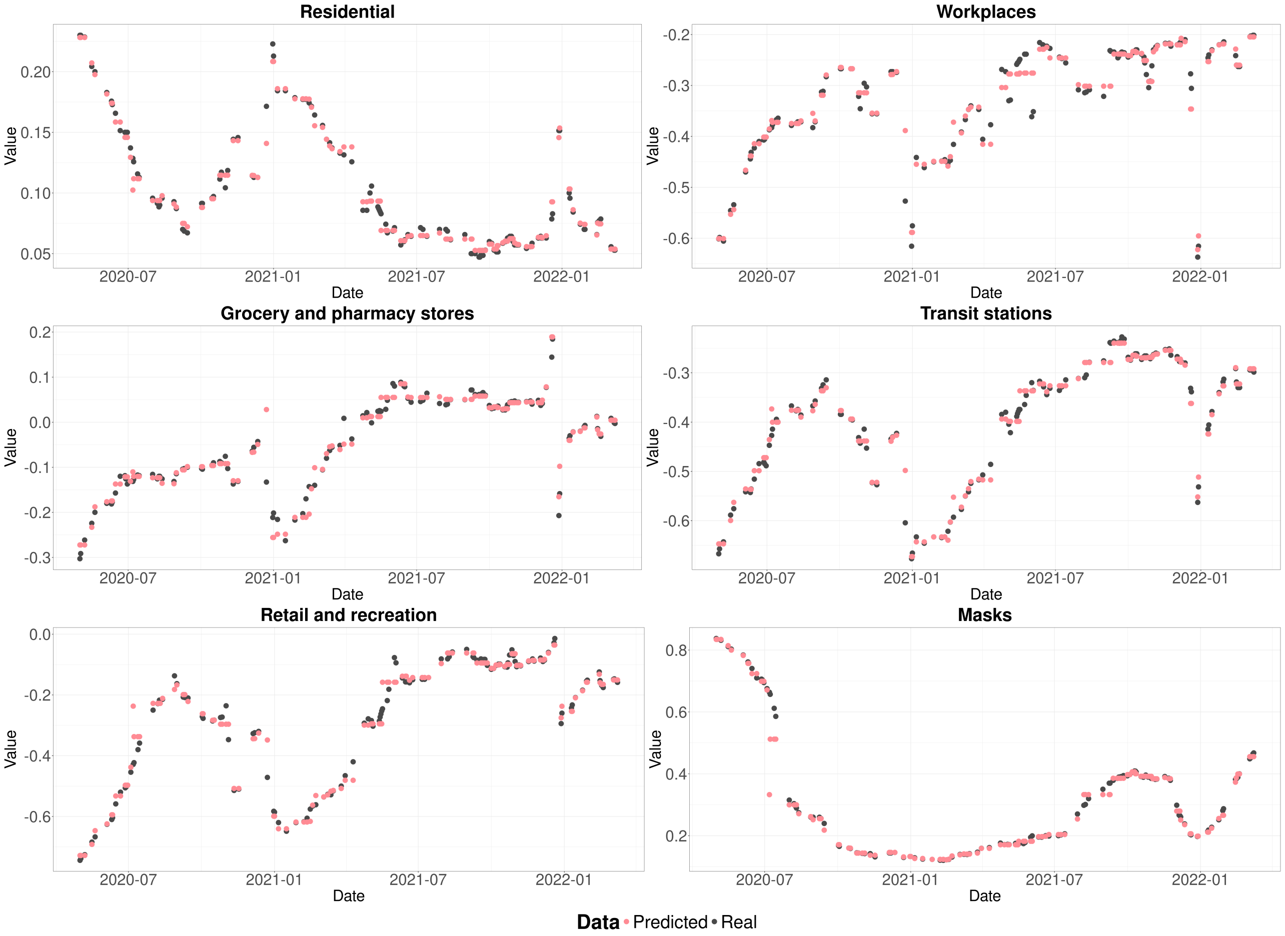

Figure SM19: Comparison between the ground truth and the predicted values for the test set (20% of the data, first fold) for the XGBoost model from NPIs to mobility in the United Kingdom.

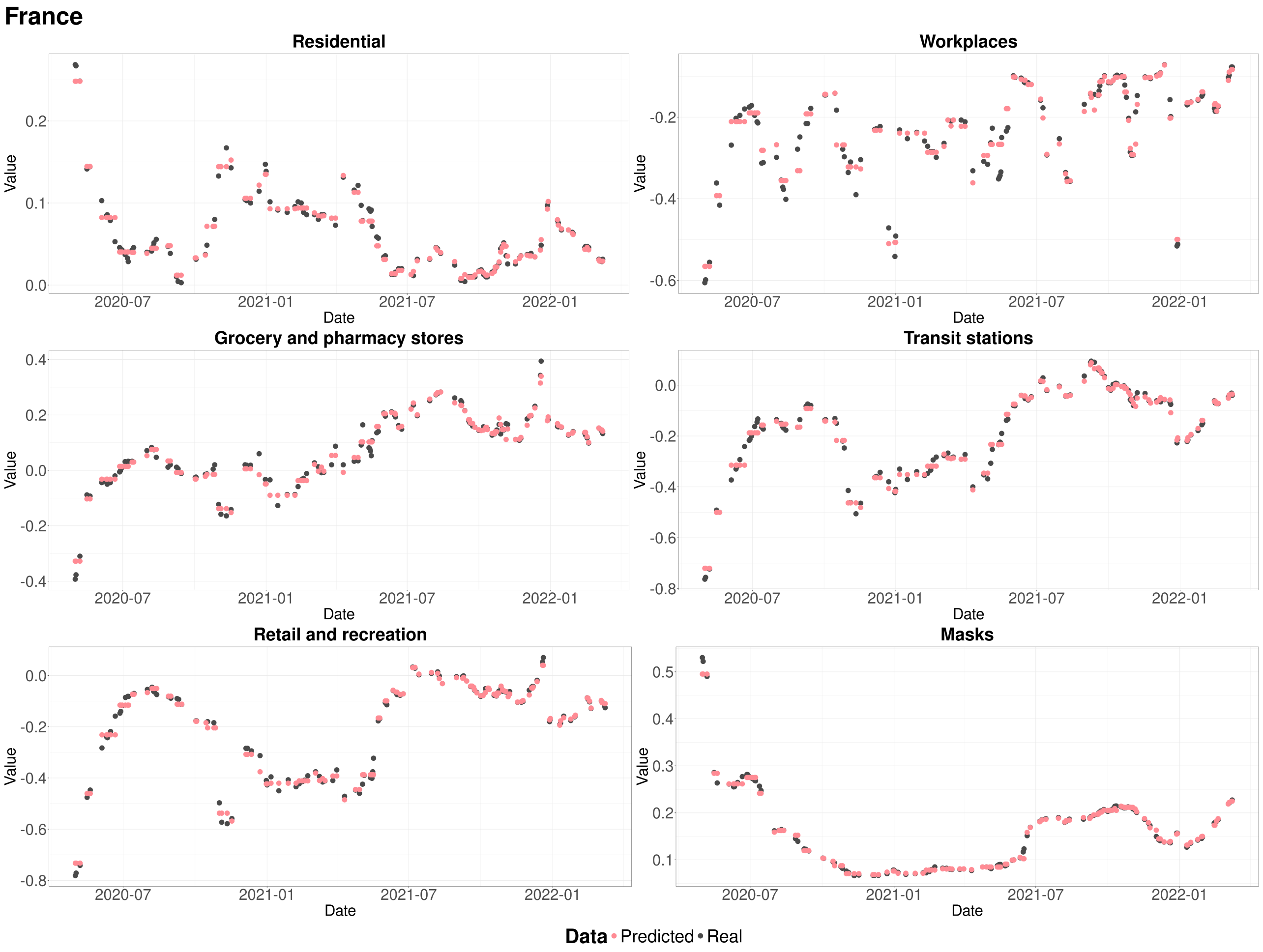

Figure SM20: Comparison between the ground truth and the predicted values for the test set (20% of the data, first fold) for the XGBoost model from NPIs to mobility in France.

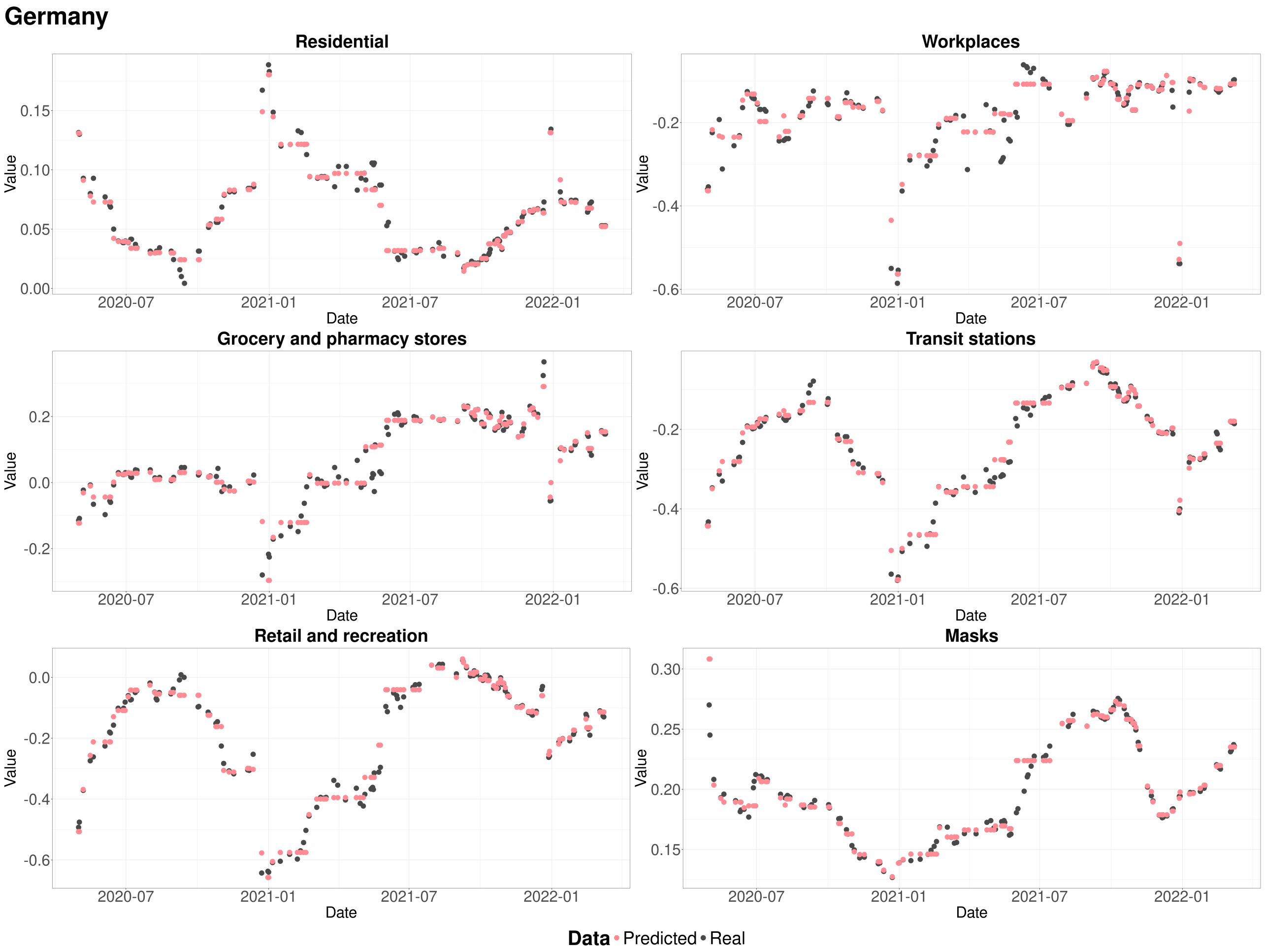

Figure SM21: Comparison between the ground truth and the predicted values for the test set (20% of the data, first fold) for the XGBoost model from NPIs to mobility in Germany.

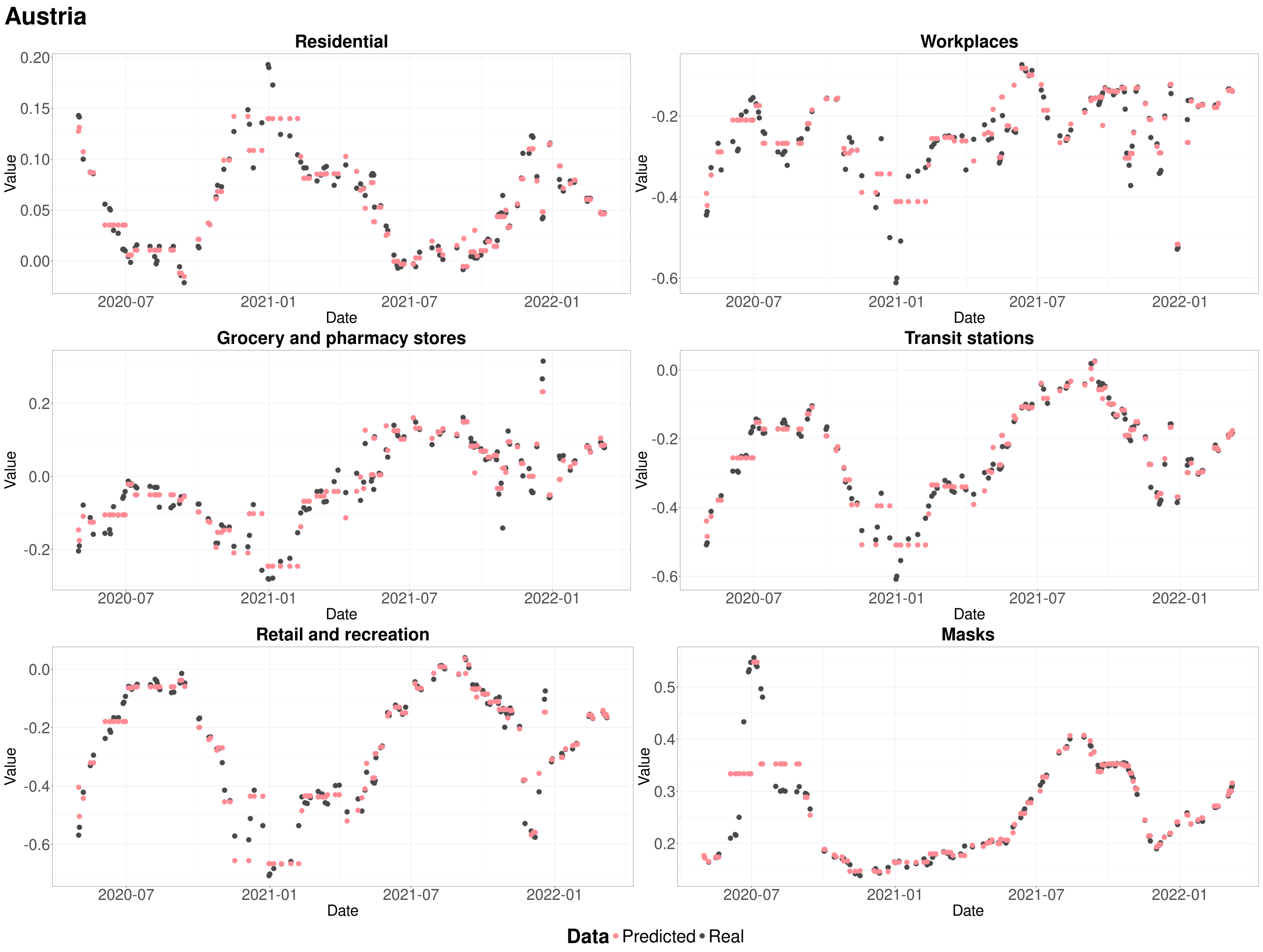

Figure SM22: Comparison between the ground truth and the predicted values for the test set (20% of the data, first fold) for the XGBoost model from NPIs to mobility in Austria.

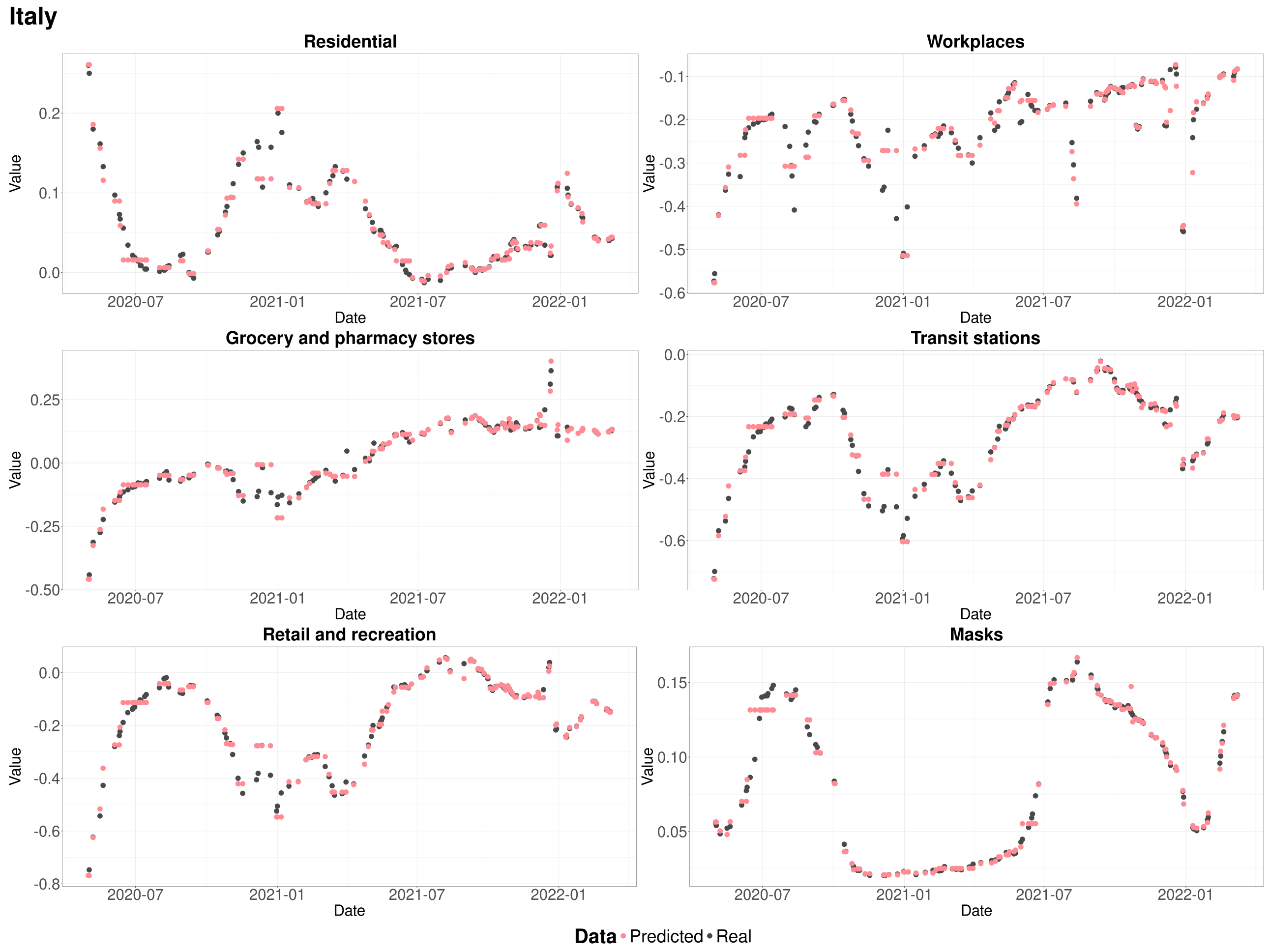

Figure SM23: Comparison between the ground truth and the predicted values for the test set (20% of the data, first fold) for the XGBoost model from NPIs to mobility in Italy.

#### Greece

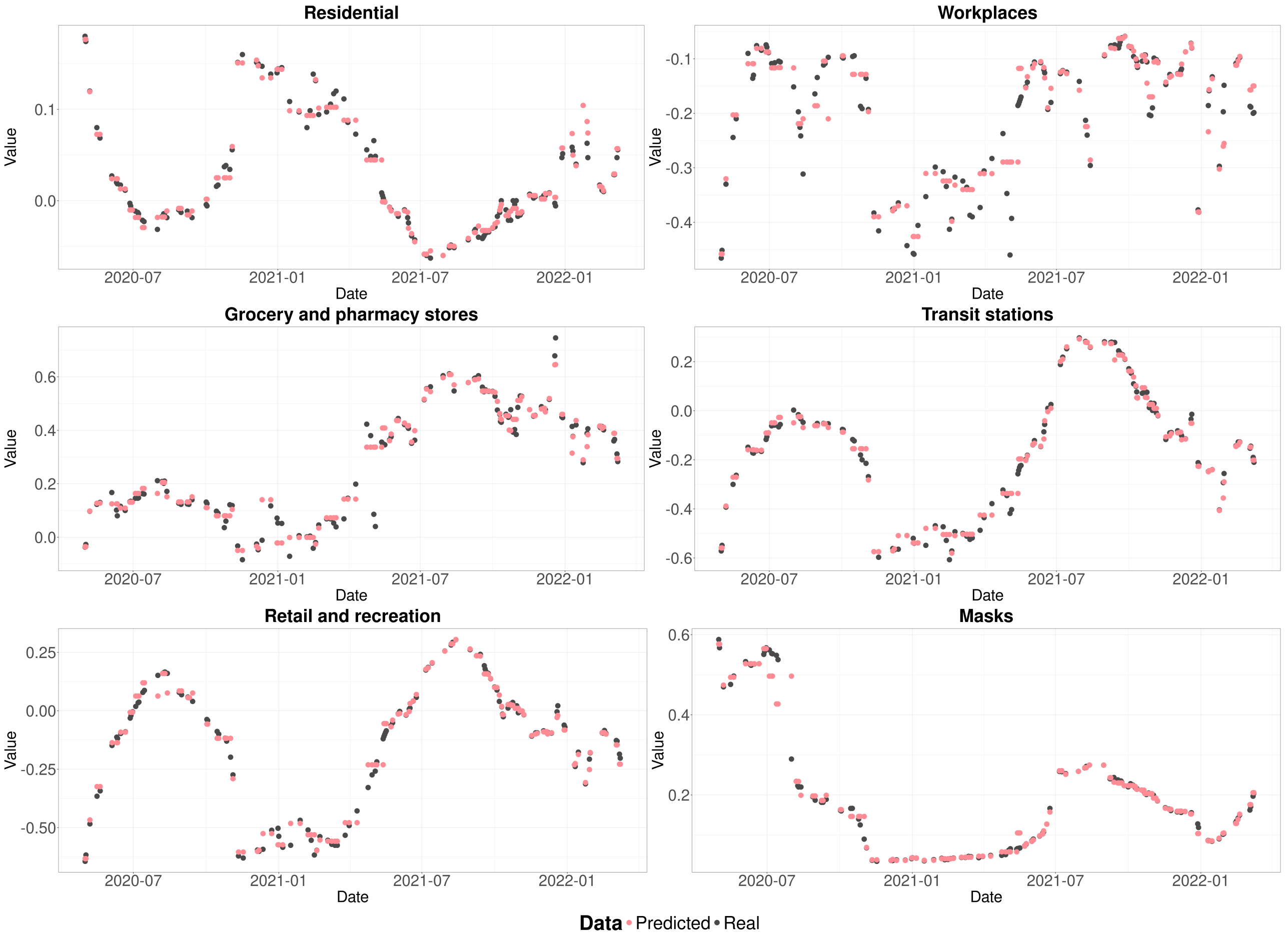

Figure SM24: Comparison between the ground truth and the predicted values for the test set (20% of the data, first fold) for the XGBoost model from NPIs to mobility in Greece.

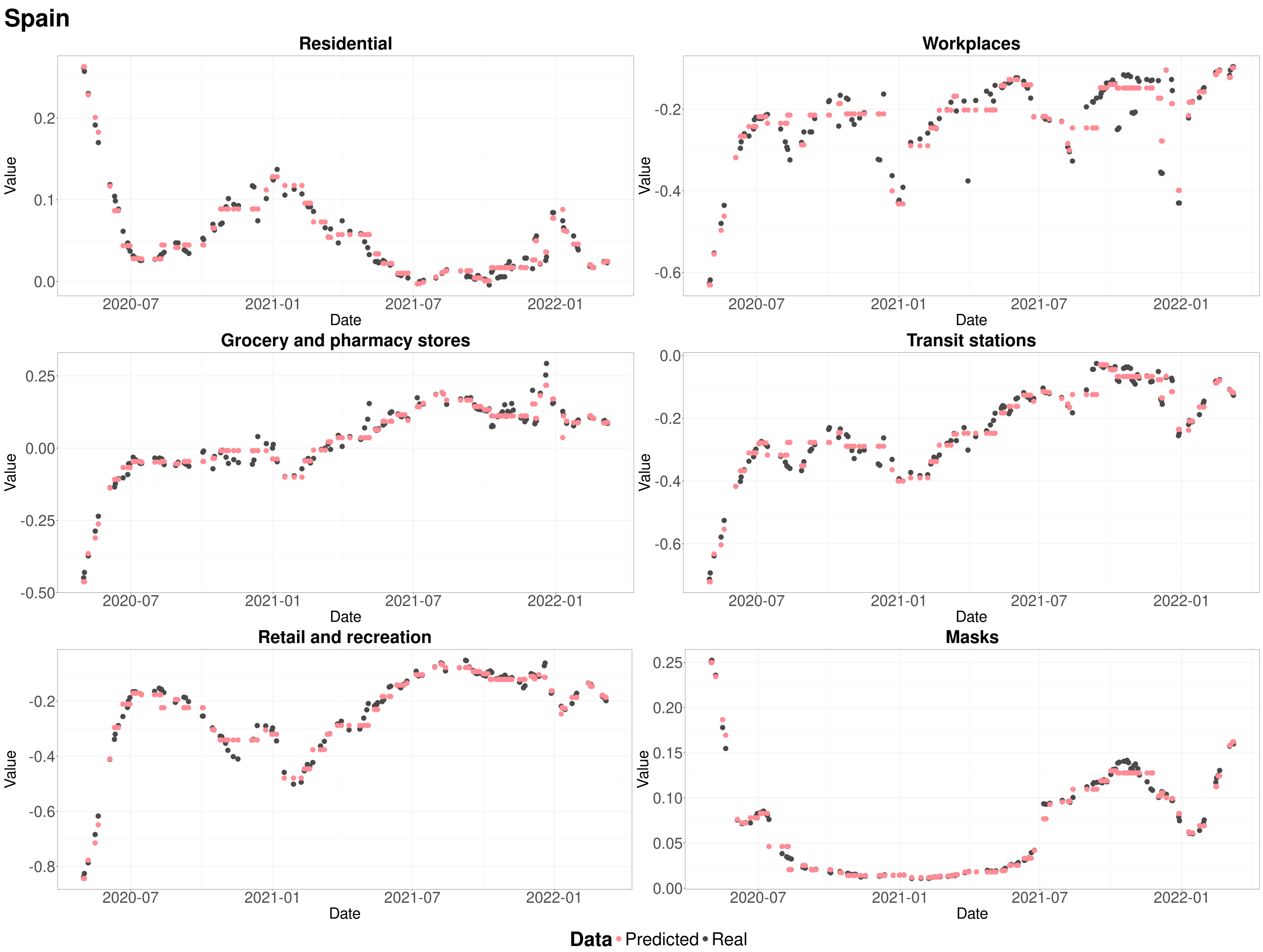

Figure SM25: Comparison between the ground truth and the predicted values for the test set (20% of the data, first fold) for the XGBoost model from NPIs to mobility in Spain.

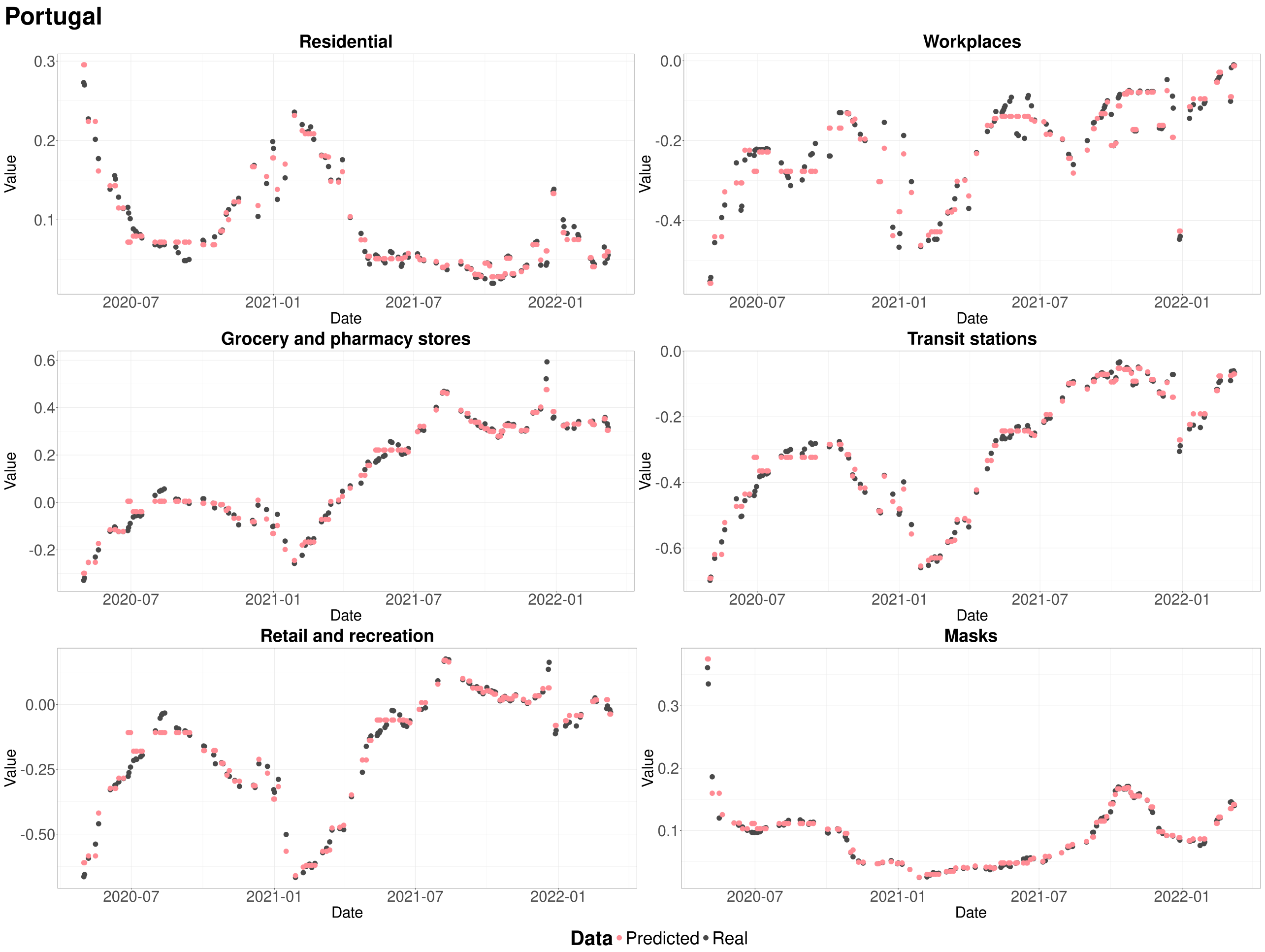

Figure SM26: Comparison between the ground truth and the predicted values for the test set (20% of the data, first fold) for the XGBoost model from NPIs to mobility in Portugal.

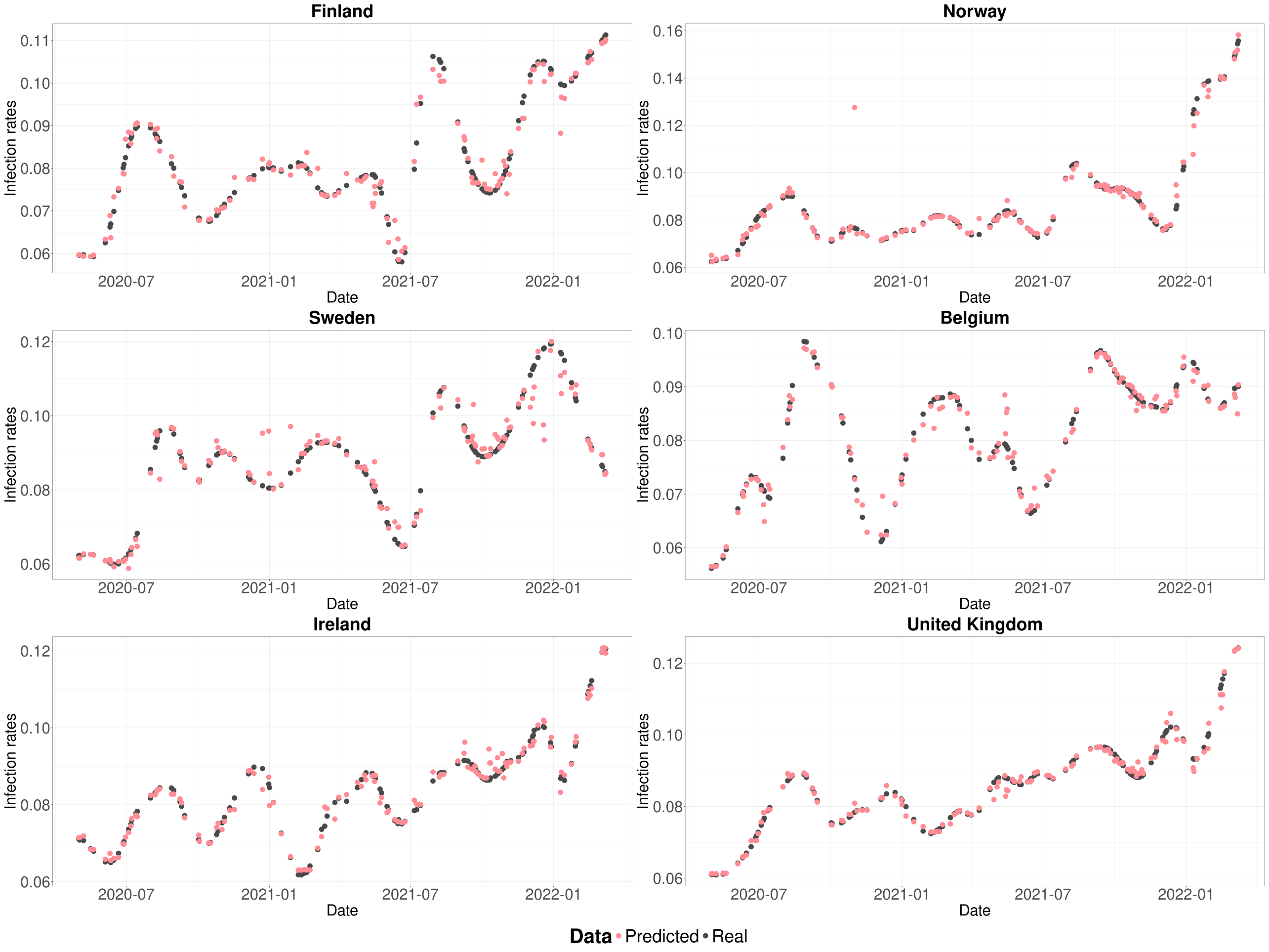

Figure SM27: Comparison between the ground truth and the predicted values for the test set (20% of the data, first fold) for the XGBoost model from mobility to infection rates in Finland, Norway, Sweden, Belgium, Ireland, and the United Kingdom.

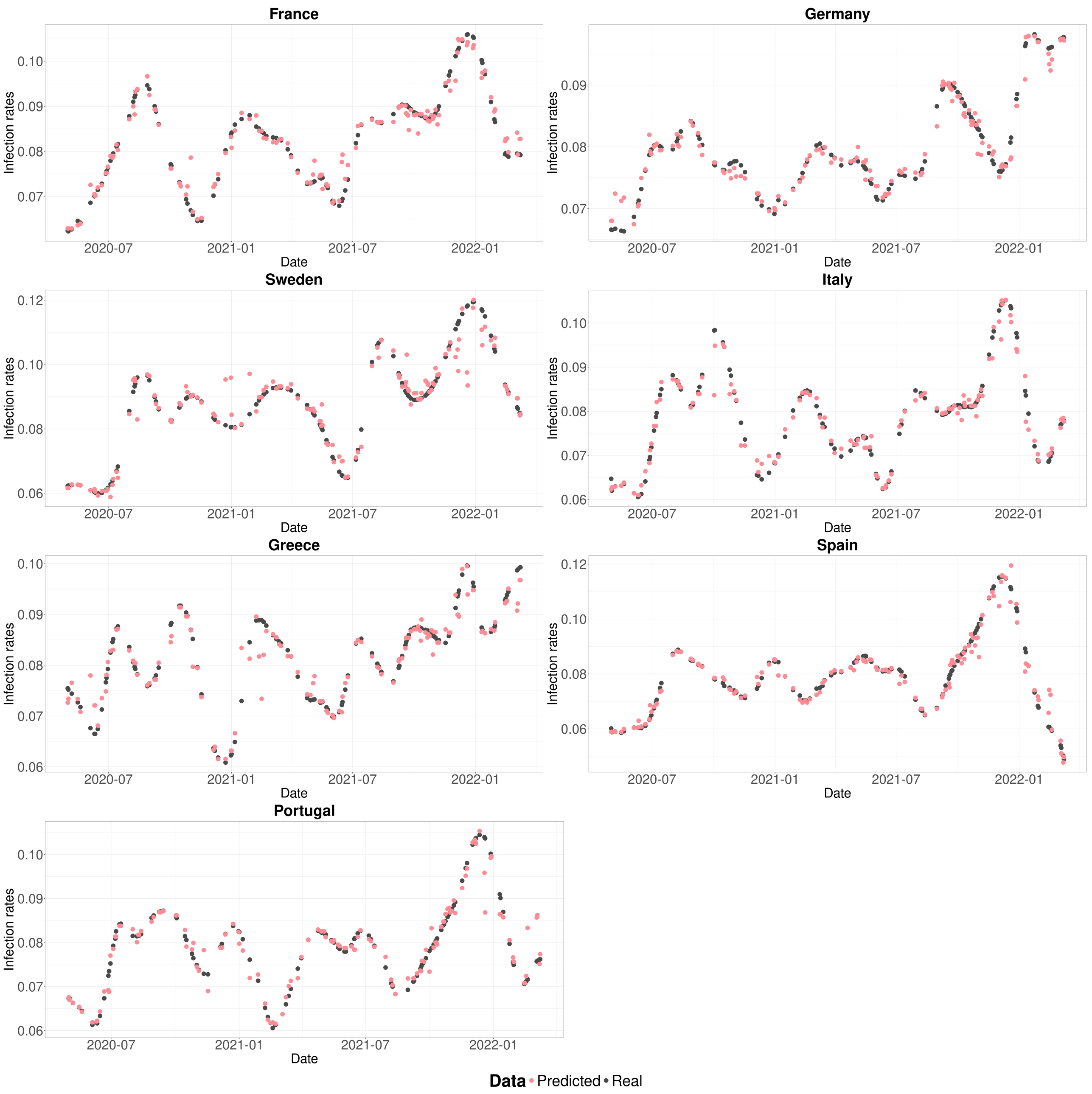

Figure SM28: Comparison between the ground truth and the predicted values for the test set (20% of the data, first fold) for the XGBoost model from mobility to infection rates in France, Germany, Austria, Italy, Greece, Spain, and Portugal.

### 5 How to reproduce the results

#### 5.1 Packages

In this study we used the R software [14] together with the following libraries: *dplyr* [24], *ggplot2* [17], *zoo* [25], *forecast* [8], *COVID19* [6, 5], *prophet* [16], *scales* [22], *patchwork* [12], *countrycode* [1], *plotfunctions* [15], *reshape* [20], *MOMAColors* [11], *xgboost* [2], *cluster* [10], *stringr* [21], *DTWBI* [18], *data.table* [4], *tidyr* [19], *scales* [23], and *connector* [13].

#### 5.2 Requirements

You need to have docker installed on your computer, for more info see this document: `https://docs.docker.com/engine/installation/`. Ensure your user has the right to run docker (without using sudo). To create the docker group and add your user in a Unix system:

- Create the docker group:

```
$ sudo groupadd docker
```

- Add your user to the docker group:

```
$ sudo usermod -aG docker $USER
```

- Log out and log back in so that your group membership is re-evaluated.

##### 5.3 Reproduce

To reproduce the results clone the repository (`https://github.com/daniele-baccega/mobility-study`) and run:

```
$ cd mobility-study
$ ./reproduce.sh
```

#### References

[1] Vincent Arel-Bundock, Nils Enevoldsen, and CJ Yetman. "countrycode: An R package to convert country names and country codes". In: *Journal of Open Source Software* 3.28 (2018), p. 848. DOI: `http://dx.doi.org/10.21105/joss.00848`.

[2] T Chen. "Xgboost: extreme gradient boosting". In: *R package version 0.4-2* 1.4 (2015). URL: `https://cran.r-project.org/web/packages/xgboost/`.

[3] David L Davies and Donald W Bouldin. "A cluster separation measure". In: *IEEE transactions on pattern analysis and machine intelligence* 2 (1979), pp. 224–227. DOI: `https://doi.org/10.1109/TPAMI.1979.4766909`.

[4] Matt Dowle et al. "Package 'data.table'". In: *Extension of 'data. frame* 596 (2019), p. 952. URL: `https://cran.r-project.org/web/packages/data.table/`.

[5] Emanuele Guidotti. "A worldwide epidemiological database for COVID-19 at fine-grained spatial resolution". In: *Scientific Data* 9.1 (2022), p. 112. DOI: `https://doi.org/10.1038/s41597-022-01245-1`.

[6] Emanuele Guidotti and David Ardia. "COVID-19 Data Hub". In: *Journal of Open Source Software* 5.51 (2020), p. 2376. DOI: `https://doi.org/10.21105/joss.02376`.

[7] John A Hartigan, Manchek A Wong, et al. "A k-means clustering algorithm". In: *Applied statistics* 28.1 (1979), pp. 100–108. DOI: `https://doi.org/10.2307/2346830`.

[8] Rob J Hyndman and Yeasmin Khandakar. "Automatic time series forecasting: the forecast package for R". In: *Journal of Statistical Software* 27.3 (2008), pp. 1–22. DOI: `https://doi.org/10.18637/jss.v027.i03`.

[9] GM James, TJ Hastie, and CA Sugar. "Principal component models for sparse functional data". In: *Biometrika* 87.3 (Sept. 2000), pp. 587–602. ISSN: 0006-3444. DOI: `10.1093/biomet/87.3.587`. eprint: `https://academic.oup.com/biomet/article-pdf/87/3/587/830673/870587.pdf`. URL: `https://doi.org/10.1093/biomet/87.3.587`.

[10] Martin Maechler et al. *cluster: Cluster Analysis Basics and Extensions*. R package version 2.1.8 — For new features, see the 'NEWS' and the 'Changelog' file in the package source). 2024. URL: `https://CRAN.R-project.org/package=cluster`.

[11] BR Mills. "MoMAColors: Color Palettes Inspired by Artwork at the Museum of Modern Art in New York City". In: *R package* (2024).

[12] Thomas Lin Pedersen. *patchwork: The Composer of Plots*. R package version 1.2.0. 2024. URL: `https://CRAN.R-project.org/package=patchwork`.

[13] Simone Pernice et al. "CONNECTOR, fitting and clustering of longitudinal data to reveal a new risk stratification system". In: *Bioinformatics* 39.5 (2023), btad201. DOI: `https://doi.org/10.1093/bioinformatics/btad201`.

[14] R Core Team. *R: A Language and Environment for Statistical Computing*. R Foundation for Statistical Computing. Vienna, Austria, 2021. URL: `https://www.R-project.org/`.

[15] Jacolien van Rij. "Package 'plotfunctions'". In: (2020). URL: `https://cran.r-project.org/web/packages/plotfunctions/`.

[16] Sean J Taylor and Benjamin Letham. “Forecasting at scale”. In: *The American Statistician* 72.1 (2018), pp. 37–45. DOI: http://dx.doi.org/10.7287/peerj.preprints.3190v2.

[17] Hadley Wickham. *ggplot2: Elegant Graphics for Data Analysis.* Springer-Verlag New York, 2016. ISBN: 978-3-319-24277-4. URL: https://ggplot2.tidyverse.org.

[18] Hadley Wickham. “Package ‘DBI’”. In: (2015). URL: https://cran.r-project.org/web/packages/DBI/.

[19] Hadley Wickham. “Package ‘tidyr’”. In: *Easily Tidy Data with’spread’and’gather ()’Functions* (2017). URL: https://cran.r-project.org/web/packages/tidyr/.

[20] Hadley Wickham. “Reshaping data with the reshape package”. In: *Journal of statistical software* 21 (2007), pp. 1–20. DOI: https://doi.org/10.18637/jss.v021.i12.

[21] Hadley Wickham. “stringr: modern, consistent string processing”. In: *The R Journal* 2.2 (2010), pp. 38–40. DOI: 10.32614/RJ-2010-012. URL: https://doi.org/10.32614/RJ-2010-012.

[22] Hadley Wickham, Thomas Lin Pedersen, and Dana Seidel. *scales: Scale Functions for Visualization.* R package version 1.3.0. 2023. URL: https://CRAN.R-project.org/package=scales.

[23] Hadley Wickham, Maintainer Hadley Wickham, and Imports RColorBrewer. “Package ‘scales’”. In: *R package version* 1.0 (2016). URL: https://cran.r-project.org/web/packages/scales/.

[24] Hadley Wickham et al. *dplyr: A Grammar of Data Manipulation.* R package version 1.1.4. 2023. URL: https://CRAN.R-project.org/package=dplyr.

[25] Achim Zeileis and Gabor Grothendieck. “zoo: S3 Infrastructure for Regular and Irregular Time Series”. In: *Journal of Statistical Software* 14.6 (2005), pp. 1–27. DOI: https://doi.org/10.18637/jss.v014.i06.
